# Physical, cognitive and mental health impacts of COVID-19 following hospitalisation – a multi-centre prospective cohort study

**DOI:** 10.1101/2021.03.22.21254057

**Authors:** PHOSP-COVID Collaborative Group, Rachael Andrea Evans, Hamish McAuley, Ewen M Harrison, Aarti Shikotra, Amisha Singapuri, Marco Sereno, Omer Elneima, Annemarie B Docherty, Nazir I Lone, Olivia C Leavy, Luke Daines, J Kenneth Baillie, Jeremy S Brown, Trudie Chalder, Anthony De Soyza, Nawar Diar Bakerly, Nicholas Easom, John R Geddes, Neil J Greening, Nick Hart, Liam G Heaney, Simon Heller, Luke Howard, Joseph Jacob, R Gisli Jenkins, Caroline Jolley, Steven Kerr, Onn M Kon, Keir Lewis, Janet M Lord, Gerry P McCann, Stefan Neubauer, Peter JM Openshaw, Paul Pfeffer, Matthew Rowland, Malcolm G Semple, Sally J Singh, Aziz Sheikh, David Thomas, Mark Toshner, James D Chalmers, Ling-Pei Ho, Alex Horsley, Michael Marks, Krisnah Poinasamy, Louise V Wain, Christopher E Brightling

## Abstract

**Background:** The impact of COVID-19 on physical and mental health, and employment following hospitalisation is poorly understood.

**Methods:** PHOSP-COVID is a multi-centre, UK, observational study of adults discharged from hospital with a clinical diagnosis of COVID-19 involving an assessment between two- and seven-months later including detailed symptom, physiological and biochemical testing. Multivariable logistic regression was performed for patient-perceived recovery with age, sex, ethnicity, body mass index (BMI), co-morbidities, and severity of acute illness as co-variates. Cluster analysis was performed using outcomes for breathlessness, fatigue, mental health, cognition and physical function.

**Findings:** We report findings of 1077 patients discharged in 2020, from the assessment undertaken a median 5 [IQR4 to 6] months later: 36% female, mean age 58 [SD 13] years, 69% white ethnicity, 27% mechanical ventilation, and 50% had at least two co-morbidities. At follow-up only 29% felt fully recovered, 20% had a new disability, and 19% experienced a health-related change in occupation. Factors associated with failure to recover were female, middle-age, white ethnicity, two or more co-morbidities, and more severe acute illness. The magnitude of the persistent health burden was substantial and weakly related to acute severity. Four clusters were identified with different severities of mental and physical health impairment: 1) Very severe (17%), 2) Severe (21%), 3) Moderate with cognitive impairment (17%), 4) Mild (46%), with 3%, 7%, 36% and 43% feeling fully recovered, respectively. Persistent systemic inflammation determined by C-reactive protein was related to cluster severity, but not acute illness severity.

**Interpretation:** We identified factors related to recovery from a hospital admission with COVID-19 and four different phenotypes relating to the severity of physical, mental, and cognitive health five months later. The implications for clinical care include the potential to stratify care and the need for a pro-active approach with wide-access to COVID-19 holistic clinical services.

<u>Funding:</u> UKRI and NIHR

## Introduction

As of March 2021, the number of reported cases of severe acute respiratory syndrome coronavirus 2 (SARS-CoV-2)) disease (COVID-19) exceeds 120 million worldwide with >2·5 million deaths. In the UK of the 4·2 million cases 450,000 have been admitted to hospital.^1^ The in-hospital mortality has reduced from initially >30% to <20%.^2^ Thus there are >300,000 post-hospitalisation survivors of COVID-19 in the UK. It is well-established that in survivorship cohorts of hospitalised patients following critical illness, prolonged morbidity with reduced functional status and impaired mental health persists for many subsequent years.^3^ In the case of COVID-19, people with lived experience of prolonged symptoms after recovery from the acute phase of infection have termed their condition ‘Long COVID’.^4^ Recently the UK National Institute for Health and Care Excellence (NICE) suggested three phases: acute, subacute, and post-covid syndrome where symptoms persist beyond 12 weeks of the initial symptoms.^5^

To date, single centre cohort studies have reported that at least 50% of those hospitalised for COVID-19 infection have ongoing symptoms at three months^6, 7^ compared to up to 30% of those who remained in the community during their acute illness.^8, 9^ The largest post-hospitalisation cohort study published to date (from Wuhan, China) reported ongoing symptoms at six months with a positive association with severity of illness.^10^ However, even in the milder group not requiring continuous supplemental oxygen during the admission, over 80% had persistent symptoms at six months.^10^

In the UK, the post-hospitalisation COVID-19 study (PHOSP-COVID) was established as a national consortium to understand and improve long-term health outcomes following COVID-19. In this first analysis, we report the outcomes at first review for patients hospitalised with COVID-19 (discharged March-November 2020). The aim was to determine the impact on health and employment, to identify factors associated with recovery and to describe recovery phenotypes.

## Methods

### Study design and participants

This prospective longitudinal cohort study recruited patients aged over 18 years old who were discharged from one of 53 National Health Service (NHS) hospitals across England, Northern Ireland, Scotland and Wales following admission to a medical assessment or ward for confirmed or clinician-diagnosed COVID-19. We excluded patients who i) had a confirmed diagnosis of a pathogen unrelated to the objectives of this study, ii) attended an accident and emergency department but were not admitted, iii) had another life-limiting illness with life expectancy less than six months such as disseminated malignancy. This analysis is restricted to participants who consented to attend two follow-up research visits within one-year post-discharge in addition to routine clinical care. Written informed consent was obtained from all study participants. The study was approved by the Leeds West Research Ethics Committee (20/YH/0225) and is registered on the ISRCTN Registry (ISRCTN10980107).

### Procedures

Participants were invited to attend a research visit between two and seven months (+/- two weeks) post-hospital discharge. Where possible this was scheduled alongside clinical follow-up and data from clinically collected assessments within four weeks of a research visit were used. A core set of data variables, including demographics, anthropometric measures, tests of physical performance, questionnaires, pulmonary physiology and biochemical tests, were obtained from the clinical records where part of clinical care or at the research visit (when not performed clinically) (Table SM1).

Patient demographics and characteristics of their acute COVID-19 admission, including results of a polymerase chain reaction (PCR) test for SARS-CoV-2, treatments and organ support received, were obtained from hospital notes by the study team at each site. Indices of Multiple Deprivation were obtained using postcode.^11^ Duration of admission was calculated using the hospital discharge date and the earliest admission date to the same or different hospital for the participant’s COVID-19 episode. Severity of acute illness was determined by the highest level of organ support received whilst admitted categorised using the WHO clinical progression scale: i) 3-4 = no continuous supplemental oxygen needed, ii) 5 = continuous supplemental oxygen only, iii) 6 = Continuous Positive Airway Pressure ventilation (CPAP), Bi-Level Positive Airway Pressure (BIPAP) or High Flow Nasal Oxygen, iv) 7-9 = Invasive Mechanical Ventilation (IMV), Extra-Corporeal Membrane Oxygenation (ECMO).^12^ For the purpose of this analysis, those receiving Renal Replacement Therapy (RRT) acutely were assigned to category 7-9. Only participants with complete data for date of admission, organ support during admission, sex at birth and attendance at a research visit 2-7 months post-discharge were included in this analysis (Figure SR1). A pre-existing comorbidity was considered absent if not indicated by a ‘yes’ on the case report form.

Patient reported outcomes were collected using the following validated questionnaires: EuroQol five-dimension five-level (EQ-5D-5L) questionnaire including the EuroQol Visual Analogue Scale (EQ-VAS)^13^, the General Anxiety Disorder 7 Questionnaire (GAD-7)^14^, the Patient Health Questionnaire-9 (PHQ-9)^15^, Post Traumatic Stress Disorder Checklist (PCL-5)^16^, Dyspnoea-12^17^, FACIT Fatigue Scale (FACIT)^18^, the Brief Pain Inventory (BPI)^19^ and the Washington Group Short Set on Functioning (WG-SS) (Table SM1 and SM2).^20^ In addition, participants completed a study specific clinical questionnaire that asked about their general recovery, symptoms and working status since their COVID-19 admission. Patients were asked if they had experienced a change in their occupational status after COVID-19 and if so, whether it was due to health, employer or other reasons. The Montreal Cognitive Assessment (MoCA)^21^ and Rockwood Clinical Frailty Scale^22^ assessments were administered by a researcher. The incremental shuttle walk test (ISWT) ^23^ and short physical performance battery (SPPB)^24^ were conducted as per recommended standards^25^ (Table SM2). Due to COVID-19 related restrictions on aerosol-generating procedures during the study period, access to spirometry and lung function was limited. Forced Expired Volume in 1 second (FEV_1_) and Forced Vital Capacity (FVC) were obtained according to ATS/ERS criteria^26^ and used to calculate the ratio FEV_1_/FVC. Transfer Capacity of the Lung for the uptake of carbon monoxide (TLCO) and carbon monoxide transfer coefficient (KCO) were obtained from the best of two repeat readings. Percent predicted values for FEV_1_, FVC TLCO and KCO were calculated^27^.

A panel of blood tests were undertaken according to the specifications of the local recruiting site service laboratory including N-terminal pro B-type natriuretic peptide (NT-BNP) or BNP, HbA1c, D-Dimer, C-reactive protein (CRP) and estimated Glomerular Filtration Rate (eGFR) (Table SM2).

### Statistical Analysis

Continuous variables were presented as median and interquartile range (IQR) or mean and standard deviation (SD). Binary and categorical variables were presented as counts and percentages (by row or by column as indicated in table legends). Participants were stratified by the severity of their acute COVID-19 illness (based on four independent categories defined by WHO), by number of pre-existing comorbidities, or by cluster (see methods below). Missing data were reported within each variable and per category. A Chi-squared test was used to identify differences in proportions across multiple categories. For normally distributed and non-normally distributed continuous data, analysis of variance (ANOVA F-test) and Kruskal Wallis tests respectively, were used to test differences across categories.

We reported univariable and multivariable logistic regression with and without imputed data where applicable. Missing values in variables were handled using multiple imputation by chained equations, under the missing at random assumption. Ten sets, each with 10 iterations, were imputed using the following variables: age, sex, ethnicity, IMD, BMI (measured at the 2-7 month follow-up visit used as a surrogate for BMI at admission), severity (WHO clinical progression scale), comorbidity categories, admission duration, treatment with steroid, treatment with antibiotics, treatment with therapeutic anticoagulation, and the outcome variable (recovered from COVID-19). Model derivation and validation was performed in imputed datasets, with Rubin’s rules^28^ used to combine results. Recovery from COVID-19 was modelled using hierarchical multivariable logistic regression, with admission hospital incorporated as a random effect. Criterion-based model building used the following principles: relevant explanatory variables were identified a priori for exploration; interactions were checked at first order level and incorporated if significant; final model selection was informed by the Akaike Information Criterion (AIC) and c-statistic, with appropriate assumptions checked including the distribution of residuals. Sensitivity analyses were performed using complete case data. The final model was presented as a forest plot with odds ratios and 95% confidence intervals.

Unsupervised clustering of patient symptom questionnaire, physical performance and cognitive assessment data (Dyspnoea-12, FACIT, GAD-7, PHQ-9, PCL-5, SPPB and MoCA as continuous variables) was performed using the clustering large applications (CLARA) k-medoids approach.^29^ Scores were centred, normalised and transformed so higher burden of disease represented higher values. A Euclidean distance metric was used and the optimal number of clusters chosen using a silhouette plot. Cluster membership was determined for each individual and characteristics presented as stratified tables.

All tests were two-tailed and p values <0·05 were considered statistically significant. We used R (version 3.6.3) with the *finalfit*, *tidyverse*, *mice*, *cluster*, and *recipes* packages for all statistical analysis.

## Results

### Cohort description

1,170 patients, discharged from hospital following treatment for COVID-19 between 5^th^ March 2020 to 30^th^ November 2020, were assessed between 14^th^ August 2020 to 5^th^ March 2021 and 1,077 were included in the analysis (Figure SR1). Overall, 35.7% were female, mean [SD] age 58 [13] years, 68·6% white ethnicity, and 29.3%, 20.6%, 50.1% of the cohort had none, one, or at least two co-morbidities, respectively (Table 1). The most common co-comorbidities were cardiovascular (42.2%), respiratory (26.4%), and type II diabetes (19.8%) (a complete list of recorded comorbidities is shown in supplementary Table SR1). The cohort demographics and pre-existing co-morbidities were similar across the severity of acute illness categories, except for a higher proportion of males (74%) in those receiving mechanical ventilation (WHO category 7-9, Table 1). Before their hospital admission, 67.5% (641/950) of participants were working either full (n=547) or part time (n=94) (Table SR2a). The median length of stay was 9 [IQR 4-21] days and 89·5% had a PCR positive test for COVID-19 at the time of admission (Table 1).

**Table 1.** Comparison of participant demographics, pre-existing co-morbidities, and admission characteristics stratified by acute illness severity by WHO clinical progression scale

|  | **WHO –**<br>**class 3-4** | **WHO –**<br>**class 5** | **WHO –**<br>**class 6** | **WHO –**<br>**class 7-9** | **Total** |
| --- | --- | --- | --- | --- | --- |
| Total N (%) | 226 (21·0) | 378 (35·1) | 185 (17·2) | 288 (26·7) | 1077 |
| Age at admission (y)†† | 54·8 (15·0) | 60·7 (12·5) | 59·1 (12·8) | 55·7 (10·9) | 57·9 (13·0) |
| Female sex at birth ¶ | 115 (50·9) | 134 (35·4) | 59 (31·9) | 76 (26·4) | 384 (35·7) |
| **Ethnicity¶** | .. | .. | .. | .. | .. |
| White | 148 (67·3) | 251 (70·3) | 130 (73·0) | 181 (64·6) | 710 (68·6) |
| South Asian | 48 (21·8) | 49 (13·7) | 28 (15·7) | 41 (14·6) | 166 (16·0) |
| Black | 18 (8·2) | 27 (7·6) | 13 (7·3) | 31 (11·1) | 89 (8·6) |
| Mixed | 5 (2·3) | 12 (3·4) | 2 (1·1) | 4 (1·4) | 23 (2·2) |
| Other | 1 (0·5) | 18 (5·0) | 5 (2·8) | 23 (8·2) | 47 (4·5) |
| *Missing Ethnicity n* | 6 | 21 | 7 | 8 | 42 |
| **Occupation¶** | .. | .. | .. | .. | .. |
| Working Full-time | 110 (55) | 172 (53) | 90 (54) | 175 (69) | 547 (58) |
| Working Part-time | 23 (11) | 31 (10) | 19 (11) | 21 (8) | 94 (10) |
| Other occupation status | 69 (34) | 124 (38 ) | 58 (35 ) | 58 (23) | 309 (33) |
| *Missing Occupation status n* | 24 | 51 | 18 | 34 | 127 |
| Healthcare worker ¶ | 52 (25) | 58 (18) | 30 (19) | 57 (21) | 197 (21) |
| *Missing Healthcare worker n* | 15 | 59 | 26 | 21 | 121 |
| **Indices of Multiple Deprivation (IMD) ¶** | .. | .. | .. | .. | .. |
| 1 | 44 (19·9) | 84 (22·6) | 30 (17·0) | 57 (20·0) | 215 (20·4) |
| 2 | 43 (19·5) | 84 (22·6) | 51 (29·0) | 64 (22·5) | 242 (23·0) |
| 3 | 45 (20·4) | 63 (17·0) | 34 (19·3) | 60 (21·1) | 202 (19·2) |
| 4 | 47 (21·3) | 71 (19·1) | 30 (17·0) | 48 (16·8) | 196 (18·6) |
| 5 | 42 (19·0) | 69 (18·6) | 31 (17·6) | 56 (19·6) | 198 (18·8) |
| *Missing IMD n* | 5 | 7 | 9 | 3 | 24 |
| Body Mass Index† | 29·1 (25·1 - 33·5) | 29·8 (26·8 - 34·0) | 32·1 (28·2 - 35·9) | 30·3 (27·7 - 34·8) | 30·1 (26·8 - 34·5) |
| BMI ≥30 ¶ | 80 (43·7) | 157 (48·6) | 97 (61·0) | 130 (53·5) | 464 (51·1) |
| *Missing BMI n* | 43 | 55 | 26 | 45 | 169 |
| Never smoker¶ | 111 (59·4) | 166 (54·2) | 80 (53·3) | 148 (59·7) | 505 (56·7) |
| Ex-smoker¶ | 69 (36·9) | 136 (44·4) | 68 (45·3) | 97 (39·1) | 370 (41·5) |
| Current smoker¶ | 7 (3·7) | 4 (1·3) | 2 (1·3) | 3 (1·2) | 16 (1·8) |
| *Missing Smoking Status n* | 39 | 72 | 35 | 40 | 186 |
| No· of comorbidities† | 1 (0 to 3) | 2 (0 to 3) | 1 (0 to 3) | 1 (0 to 3) | 2 (0 to 3) |
| 1 comorbidity¶ | 45 (19·9) | 75 (19·8) | 44 (23·8) | 58 (20·1) | 222 (20·6) |
| 2+ comorbidities¶ | 104 (46·0) | 206 (54·5) | 92 (49·7) | 138 (47·9) | 540 (50·1) |
| Cardiovascular¶ | 74 (32·7) | 176 (46·6) | 82 (44·3) | 123 (42·7) | 455 (42·2) |
| Renal & Endocrine ¶ | 23 (10·2) | 48 (12·7) | 11 (5·9) | 31 (10·8) | 113 (10·5) |
| Respiratory ¶ | 56 (24·8) | 105 (27·8) | 54 (29·2) | 69 (24·0) | 284 (26·4) |
| Type II Diabetes ¶§ | 30 (13·3) | 80 (21·2) | 40 (21·6) | 63 (21·9) | 213 (19·8) |
| **Admission** | .. | .. | .. | .. | .. |
| Admission duration (days) | 2 (1 - 6) | 6 (4 - 9) | 10 (6 - 15) | 33 (21 - 53) | 9 (4 - 21) |
| PCR COVID-19 +ve ¶ | 176 (84·2) | 319 (90·6) | 156 (92·3) | 243 (90·3) | 894 (89·5) |
| *Missing PCR result* | *17* | *26* | *16* | *19* | *78* |
| Systemic steroids ¶ | 22 (10·2) | 115 (31·5) | 57 (32·8) | 107 (45·9) | 301 (30·5) |
| *Missing steroids n* | *11* | *13* | *11* | *55* | *90* |
| Antibiotic therapy ¶ | 103 (47·5) | 313 (85·1) | 165 (91·7) | 260 (96·7) | 841 (81·3) |
| *Missing antibiotic n* | *9* | *10* | *5* | *19* | *43* |
| Anti-coagulation ¶ | 32 (15·1) | 89 (24·5) | 63 (35·4) | 137 (56·8) | 321 (32·3) |
| *Missing anti-coagulation n* | *14* | *14* | *7* | *47* | *82* |

Number (%) unless †median [IQR], ††mean [SD], n, column proportions, % calculated by category after exclusion of missing data for that variable, § = type I diabetes n = 8 (of which n=6 in category 7-9) are within the Renal & Endocrine category, PCR= Polymerase Chain Reaction, IMD = Indices of multiple deprivation with 1 = most deprived and 5 =least deprived, Anticoagulation = therapeutic dose anticoagulation –does not include intermediate doses which were not recorded. WHO = World Health Organisation. Category 3-4 = no continuous supplemental oxygen needed, 5= continuous supplemental oxygen only, 6= Continuous or Bi-level Positive Airway Pressure ventilation or High Flow Nasal O<sub>2</sub>, 7-9 = Invasive Mechanical Ventilation or other organ support

### Follow-up assessments at five months stratified by severity of acute illness

Study assessments were conducted a median 5 [IQR 4-6] months after discharge from hospital (Table 2). Over 50% of the cohort were obese (BMI categories Table SR3). Only 28.8% of the cohort described themselves as ‘fully recovered’. Of those working before COVID-19 17.8% (113/641) were no longer working, and 19.3% (124/641) experienced a health-related change in their occupational status. 47.8% (54/113) were no longer working and 55% (68/124) experiencing a health-related change in occupational status were in WHO category 7-9 (Table SR2b).

**Table 2.**
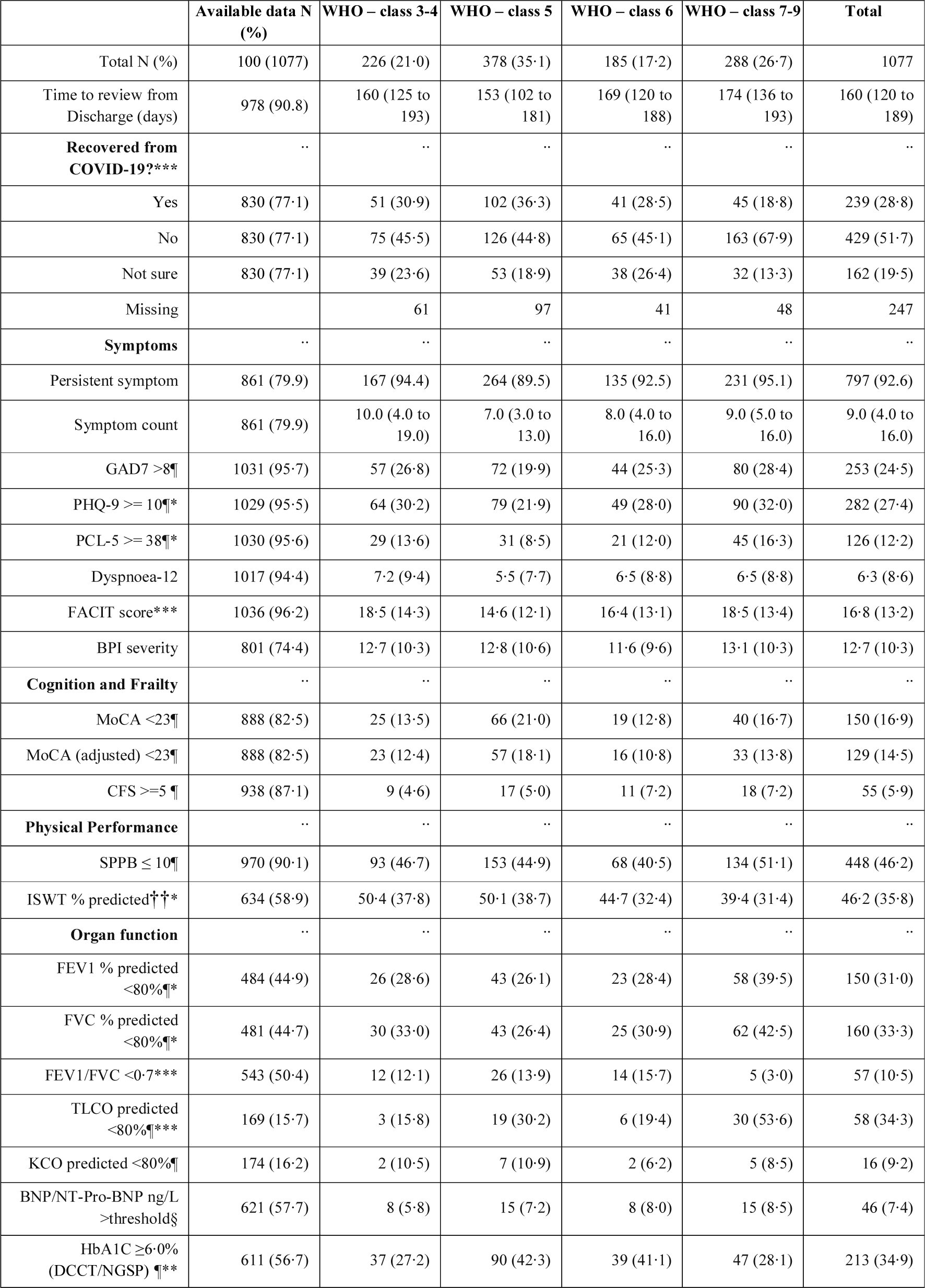
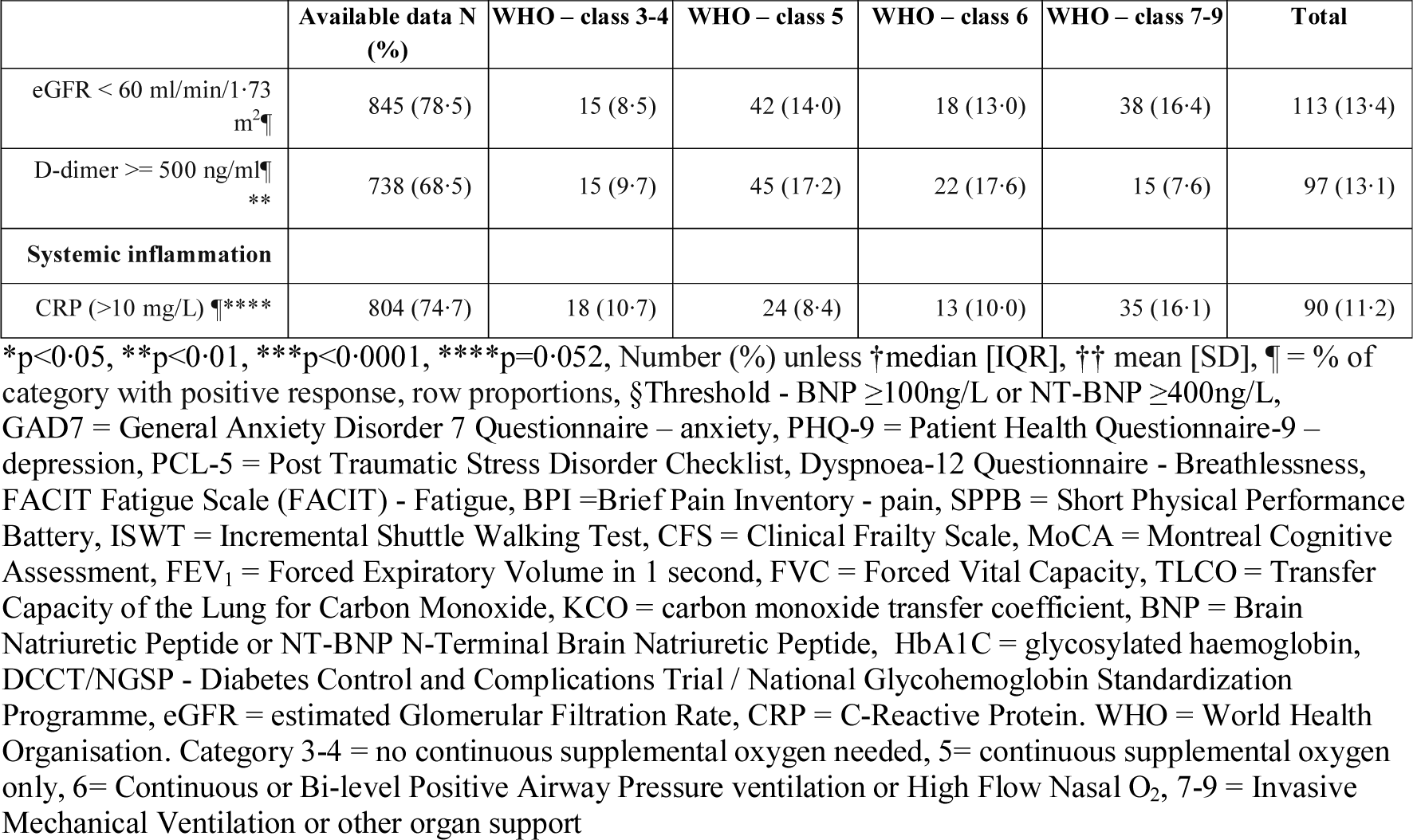
Patient reported outcome measures, physiological and biochemical tests stratified by acute illness severity (WHO clinical progression scale)

|  | Available data N (%) | WHO – class 3-4 | WHO – class 5 | WHO – class 6 | WHO – class 7-9 | Total |
| --- | --- | --- | --- | --- | --- | --- |
| Total N (%) | 100 (1077) | 226 (21.0) | 378 (35.1) | 185 (17.2) | 288 (26.7) | 1077 |
| Time to review from Discharge (days) | 978 (90.8) | 160 (125 to 193) | 153 (102 to 181) | 169 (120 to 188) | 174 (136 to 193) | 160 (120 to 189) |
| **Recovered from COVID-19\*\*\*** | .. | .. | .. | .. | .. | .. |
| Yes | 830 (77.1) | 51 (30.9) | 102 (36.3) | 41 (28.5) | 45 (18.8) | 239 (28.8) |
| No | 830 (77.1) | 75 (45.5) | 126 (44.8) | 65 (45.1) | 163 (67.9) | 429 (51.7) |
| Not sure | 830 (77.1) | 39 (23.6) | 53 (18.9) | 38 (26.4) | 32 (13.3) | 162 (19.5) |
| Missing |  | 61 | 97 | 41 | 48 | 247 |
| **Symptoms** | .. | .. | .. | .. | .. | .. |
| Persistent symptom | 861 (79.9) | 167 (94.4) | 264 (89.5) | 135 (92.5) | 231 (95.1) | 797 (92.6) |
| Symptom count | 861 (79.9) | 10.0 (4.0 to 19.0) | 7.0 (3.0 to 13.0) | 8.0 (4.0 to 16.0) | 9.0 (5.0 to 16.0) | 9.0 (4.0 to 16.0) |
| GAD7 >8¶ | 1031 (95.7) | 57 (26.8) | 72 (19.9) | 44 (25.3) | 80 (28.4) | 253 (24.5) |
| PHQ-9 >= 10¶* | 1029 (95.5) | 64 (30.2) | 79 (21.9) | 49 (28.0) | 90 (32.0) | 282 (27.4) |
| PCL-5 >= 38¶* | 1030 (95.6) | 29 (13.6) | 31 (8.5) | 21 (12.0) | 45 (16.3) | 126 (12.2) |
| Dyspnoea-12 | 1017 (94.4) | 7.2 (9.4) | 5.5 (7.7) | 6.5 (8.8) | 6.5 (8.8) | 6.3 (8.6) |
| FACIT score*** | 1036 (96.2) | 18.5 (14.3) | 14.6 (12.1) | 16.4 (13.1) | 18.5 (13.4) | 16.8 (13.2) |
| BPI severity | 801 (74.4) | 12.7 (10.3) | 12.8 (10.6) | 11.6 (9.6) | 13.1 (10.3) | 12.7 (10.3) |
| **Cognition and Frailty** | .. | .. | .. | .. | .. | .. |
| MoCA <23¶ | 888 (82.5) | 25 (13.5) | 66 (21.0) | 19 (12.8) | 40 (16.7) | 150 (16.9) |
| MoCA (adjusted) <23¶ | 888 (82.5) | 23 (12.4) | 57 (18.1) | 16 (10.8) | 33 (13.8) | 129 (14.5) |
| CFS >=5 ¶ | 938 (87.1) | 9 (4.6) | 17 (5.0) | 11 (7.2) | 18 (7.2) | 55 (5.9) |
| **Physical Performance** | .. | .. | .. | .. | .. | .. |
| SPPB ≤ 10¶ | 970 (90.1) | 93 (46.7) | 153 (44.9) | 68 (40.5) | 134 (51.1) | 448 (46.2) |
| ISWT % predicted††* | 634 (58.9) | 50.4 (37.8) | 50.1 (38.7) | 44.7 (32.4) | 39.4 (31.4) | 46.2 (35.8) |
| **Organ function** | .. | .. | .. | .. | .. | .. |
| FEV1 % predicted <80%¶* | 484 (44.9) | 26 (28.6) | 43 (26.1) | 23 (28.4) | 58 (39.5) | 150 (31.0) |
| FVC % predicted <80%¶* | 481 (44.7) | 30 (33.0) | 43 (26.4) | 25 (30.9) | 62 (42.5) | 160 (33.3) |
| FEV1/FVC <0.7*** | 543 (50.4) | 12 (12.1) | 26 (13.9) | 14 (15.7) | 5 (3.0) | 57 (10.5) |
| TLCO predicted <80%¶*** | 169 (15.7) | 3 (15.8) | 19 (30.2) | 6 (19.4) | 30 (53.6) | 58 (34.3) |
| KCO predicted <80%¶ | 174 (16.2) | 2 (10.5) | 7 (10.9) | 2 (6.2) | 5 (8.5) | 16 (9.2) |
| BNP/NT-Pro-BNP ng/L >threshold§ | 621 (57.7) | 8 (5.8) | 15 (7.2) | 8 (8.0) | 15 (8.5) | 46 (7.4) |
| HbA1C ≥6.0% (DCCT/NGSP) ¶*** | 611 (56.7) | 37 (27.2) | 90 (42.3) | 39 (41.1) | 47 (28.1) | 213 (34.9) |
| eGFR < 60 ml/min/1.73 m<sup>2</sup>¶ | 845 (78.5) | 15 (8.5) | 42 (14.0) | 18 (13.0) | 38 (16.4) | 113 (13.4) |
| D-dimer >= 500 ng/ml¶<br>** | 738 (68.5) | 15 (9.7) | 45 (17.2) | 22 (17.6) | 15 (7.6) | 97 (13.1) |
| **Systemic inflammation** |  |  |  |  |  |  |
| CRP (>10 mg/L) ¶**** | 804 (74.7) | 18 (10.7) | 24 (8.4) | 13 (10.0) | 35 (16.1) | 90 (11.2) |

\*p<0.05, \*\*p<0.01, \*\*\*p<0.0001, \*\*\*\*p=0.052, Number (%) unless †median [IQR], †† mean [SD], ¶ = % of category with positive response, row proportions, §Threshold - BNP ≥100ng/L or NT-BNP ≥400ng/L, GAD7 = General Anxiety Disorder 7 Questionnaire – anxiety, PHQ-9 = Patient Health Questionnaire-9 – depression, PCL-5 = Post Traumatic Stress Disorder Checklist, Dyspnoea-12 Questionnaire - Breathlessness, FACIT Fatigue Scale (FACIT) - Fatigue, BPI =Brief Pain Inventory - pain, SPPB = Short Physical Performance Battery, ISWT = Incremental Shuttle Walking Test, CFS = Clinical Frailty Scale, MoCA = Montreal Cognitive Assessment, FEV<sub>1</sub> = Forced Expiratory Volume in 1 second, FVC = Forced Vital Capacity, TLCO = Transfer Capacity of the Lung for Carbon Monoxide, KCO = carbon monoxide transfer coefficient, BNP = Brain Natriuretic Peptide or NT-BNP N-Terminal Brain Natriuretic Peptide, HbA1C = glycosylated haemoglobin, DCCT/NGSP - Diabetes Control and Complications Trial / National Glycohemoglobin Standardization Programme, eGFR = estimated Glomerular Filtration Rate, CRP = C-Reactive Protein. WHO = World Health Organisation. Category 3-4 = no continuous supplemental oxygen needed, 5= continuous supplemental oxygen only, 6= Continuous or Bi-level Positive Airway Pressure ventilation or High Flow Nasal O<sub>2</sub>, 7-9 = Invasive Mechanical Ventilation or other organ support

Factors associated with worse recovery, defined by patient-perceived recovery, were female sex, white ethnicity, the presence of two or more pre-existing co-morbidities and WHO category 7-9 during the acute illness (Figure 1). Age had a non-linear association, with age groups <30 years and >70 years perceiving better recovery than those aged 50-59 years (Figure 1). Recovery was associated with a BMI < 30 kg/m^2^ in the univariable and multivariable analyses but not in imputed models (Table SR4). There was no association between receiving systemic steroids during admission and recovery. The findings between the imputed and non-imputed models were similar (Table SR4).

**Figure 1.**
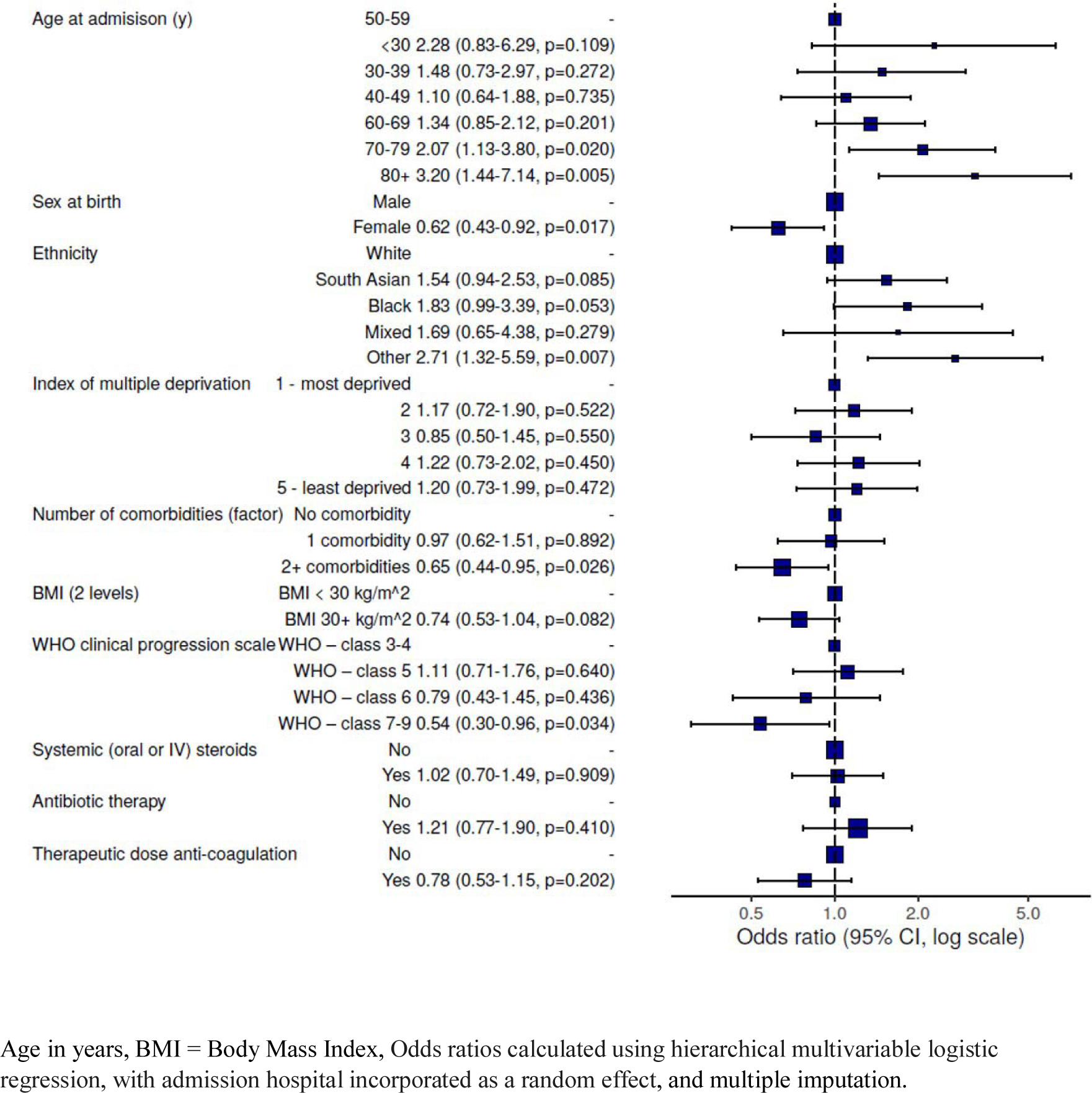
Forest plot of the patient and admission characteristics associated with recovery using multi-variable logistic regression and multiple imputation

92.8 % had at least one persistent symptom with a median (IQR) number of 9 (4 to 16) symptoms (Figure SR2). The ten most commonly reported symptoms were ‘aching …muscles (pain)’, ‘fatigue’, ‘physical slowing down’, ‘impaired sleep quality’, ‘joint pain or swelling’, ‘limb weakness’, ‘breathlessness’, ‘pain’, ‘short-term memory loss’ and ‘slowing down in …thinking’. The number of persistent symptoms was highest in those with pre-existing co-morbidities (10 [IQR 5 to 17], but high also in those without pre-existing co-morbidity (median of 7 [IQR 2 to 13]) (Table SR5).

Patient reported outcome measures alongside measures of physical performance, pulmonary physiology and biochemistry are shown in Table 2 (further details Table SR3) for the cohort, stratified by WHO clinical progression scale. Over 25% of the cohort had clinically significant symptoms of anxiety and depression, and 12.2% had symptoms of post-traumatic stress disorder. Physical performance measured by the incremental shuttle walking distance was 46.2% predicted for the cohort and 46.2% scored ≤10 on the SPPB, a marker of functional impairment. The severity of acute illness and patient reported outcomes of mental health, breathlessness, fatigue or pain or cognitive impairment were mostly unrelated (Table 2). The percent predicted ISWT distance was lower in WHO category 7-9, and there was a higher proportion of individuals with a TLCO <80% predicted, but otherwise there were no clear relationship between measures of organ function at five months and the spectrum of acute severity of illness. A greater proportion of people in WHO category 7-9 were no longer working after hospitalisation from COVID-19 and were more likely to experience change in occupation due to health after COVID-19 (Table SR2b).

### Change in symptoms, health-related quality of life and disability since hospital admission

Patients rated their EQ5D-5L VAS 0-100 worse than before the hospital admission with the greatest decrement seen in WHO category 7-9 (p<0.001) with a similar association with the EQ5D-5L utility index and all the domains of the EQ5D (Figure 2 and Table SR6a). At least a fifth of the population reached the threshold for a new disability with at least one domain coded as “a lot of difficulty” or “cannot do it all” on the WG-SS (Table SR6b). 56.2%, 48.1%, 41.8%, and 38.7% of the cohort reported significant worse symptoms of fatigue, breathlessness, sleep, and pain, respectively (Table SR6c).

**Figure 2.**
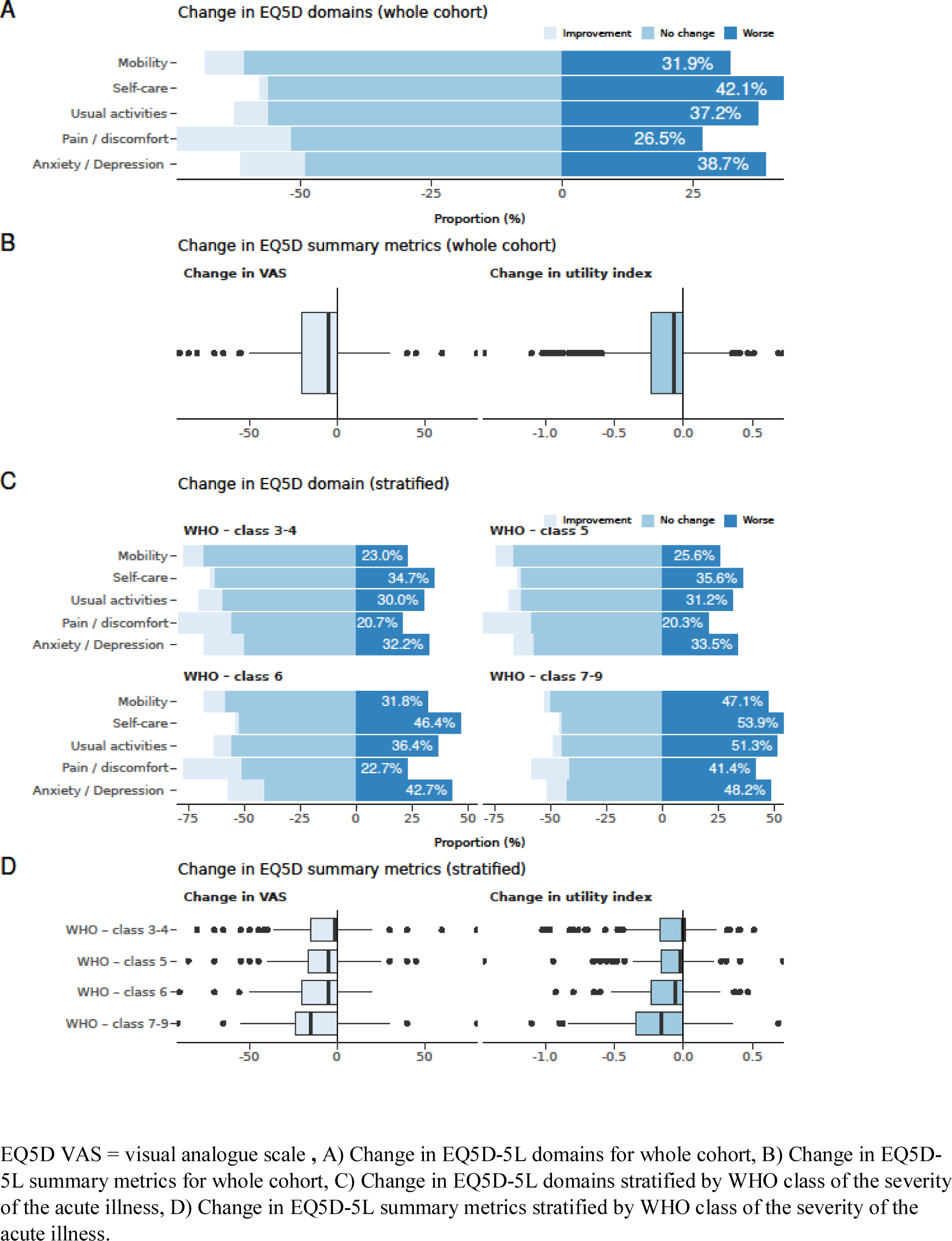
Health-related quality of life measured by the EQ5D-5L at follow-up compared to prior to admission with COVID-19

### Investigating phenotypes of recovery

Using patient reported outcome measures for symptoms, mental health including PTSD, MoCA for cognitive impairment, and SPPB for physical performance, four clusters were identified (Figure SR3 and Table SR7). Of the outcomes used in the cluster analysis all were closely related except for cognition (Figure 3 and Figure SR3). A comparison between clusters is shown in Table 3 for demographics. Figure 3 illustrates the four clusters with the predominant associated demographics, symptoms, and physical function. Cluster 1: very severe mental and physical health impairment (17%) - most obese, co-morbid, Cluster 2: severe mental and physical health impairment (21%) - obese, co-morbid, Cluster 3: moderate mental and physical health impairment with pronounced cognitive impairment (17%) - older, male predominance, overweight, and less co-morbid, Cluster 4: mild mental and physical health impairment (46%) – male predominance, overweight, less co-morbid. Respiratory and neuropsychiatric co-morbidities were more common in cluster 1 and 2 and rheumatological co-morbidities in cluster 1 than the other clusters. The indices of multiple deprivation were worse in patients in cluster 1 and 3 than cluster 2 and 4. There was no association between clusters and WHO clinical progression scale during the acute admission.

**Figure 3.**
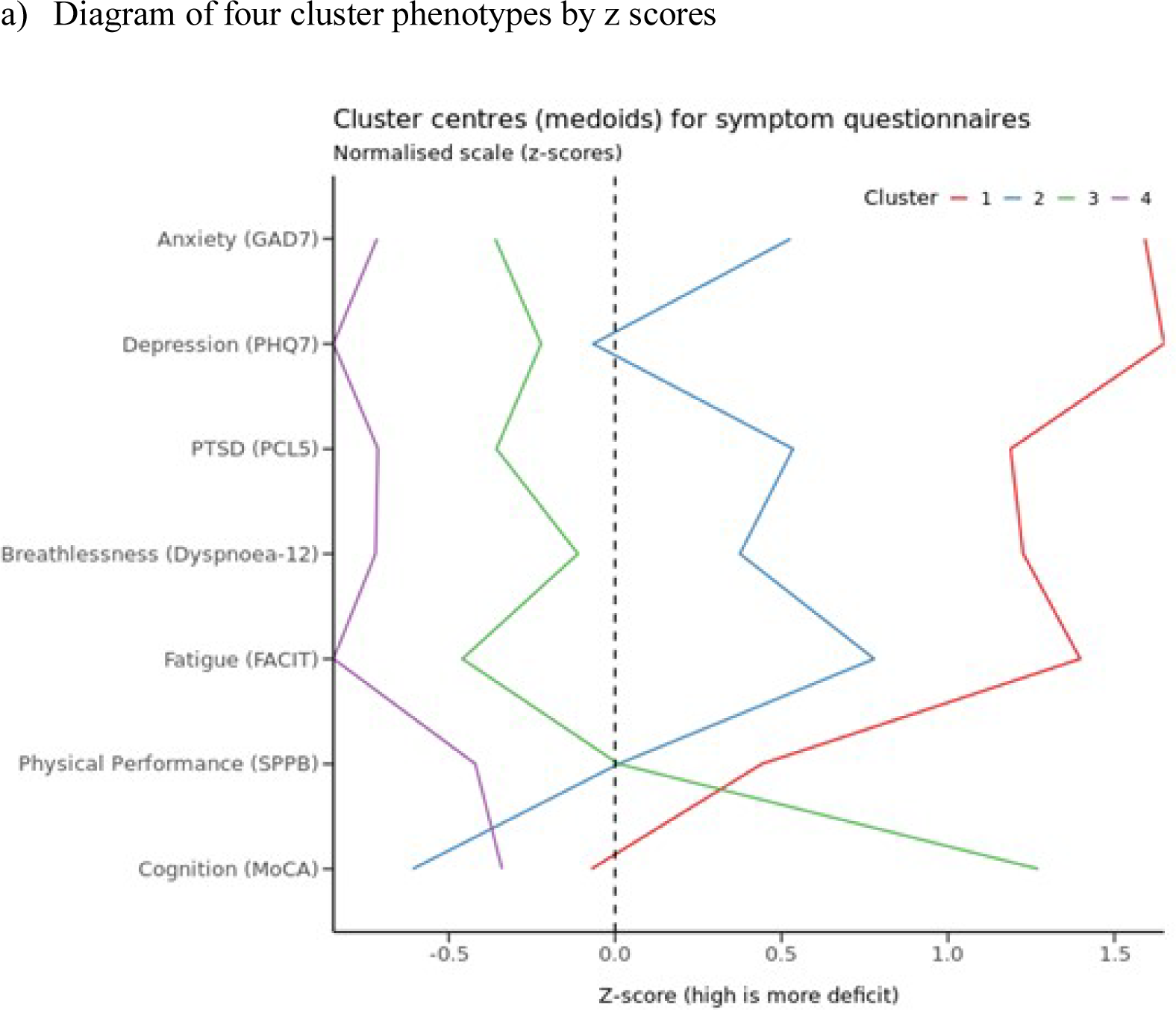
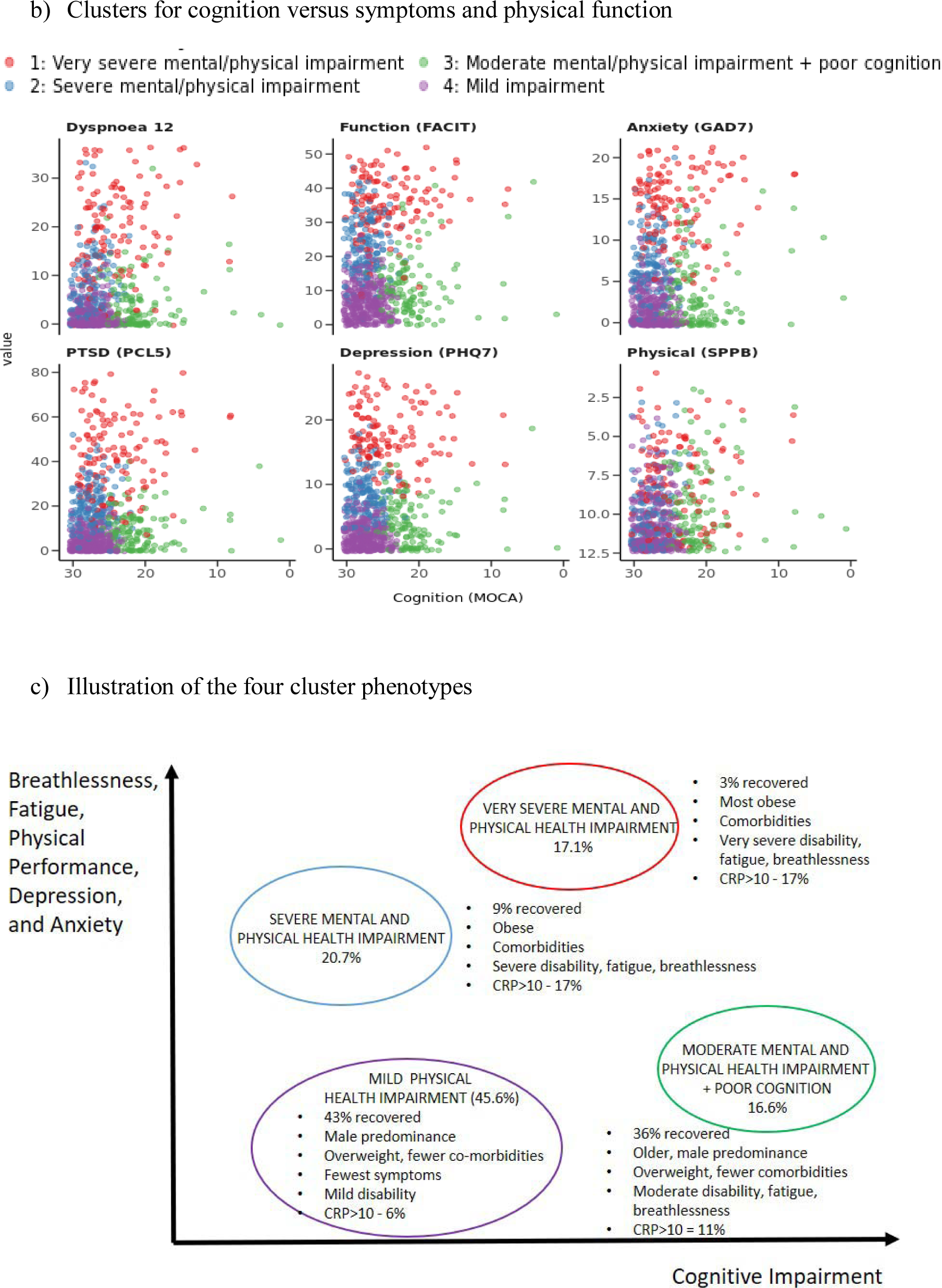
Clusters of mental, cognitive, and physical health impairments

**Table 3.** Patient and admission characteristics of the four recovery clusters

|  | Cluster 1 | Cluster 2 | Cluster 3 | Cluster 4 | Total |
| --- | --- | --- | --- | --- | --- |
| Total N (%) | 131 (17.1) | 159 (20.7) | 127 (16.6) | 350 (45.6) | 767 |
| Age (y)††*** | 55.0 (10.3) | 55.0 (11.2) | 63.2 (13.2) | 57.0 (13.2) | 57.2 (12.6) |
| Female *** | 60 (45.8) | 73 (45.9) | 46 (36.2) | 99 (28.3) | 278 (36.2) |
| Ethnicity White | 94 (74.6) | 118 (78.1) | 77 (61.1) | 242 (71.2) | 531 (71.5) |
| South Asian | 12 (9.5) | 20 (13.2) | 25 (19.8) | 60 (17.6) | 117 (15.7) |
| Black | 11 (8.7) | 6 (4.0) | 15 (11.9) | 17 (5.0) | 49 (6.6) |
| Mixed | 2 (1.6) | 2 (1.3) | 3 (2.4) | 6 (1.8) | 13 (1.7) |
| Other | 7 (5.6) | 5 (3.3) | 5 (4.0) | 8 (2.4) | 25 (3.4) |
| Missing ethnicity n | 5 | 8 | 1 | 10 | 24 |
| IMD 1 *** | 37 (29.4) | 24 (15.2) | 31 (25.0) | 53 (15.2) | 145 (19.2) |
| 2 | 32 (25.4) | 36 (22.8) | 39 (31.5) | 67 (19.3) | 174 (23.0) |
| 3 | 19 (15.1) | 29 (18.4) | 19 (15.3) | 80 (23.0) | 147 (19.4) |
| 4 | 20 (15.9) | 32 (20.3) | 22 (17.7) | 72 (20.7) | 146 (19.3) |
| 5 | 18 (14.3) | 37 (23.4) | 13 (10.5) | 76 (21.8) | 144 (19.0) |
| Missing IMD n | 5 | 1 | 3 | 2 | 11 |
| BMI kg/m<sup>2</sup>†*** | 33.5 (29.0 to 38.5) | 31.4 (28.0 to 35.7) | 29.2 (25.9 to 32.3) | 29.6 (26.5 to 33.5) | 30.3 (27.1 to 34.9) |
| BMI ≥30 kg/m<sup>2</sup>*** | 79 (68.1) | 86 (60.6) | 42 (38.2) | 136 (47.2) | 343 (52.3) |
| Missing BMI n | 15 | 17 | 17 | 62 | 111 |
| Never smoker* | 61 (53.0) | 70 (51.5) | 65 (60.2) | 181 (62.2) | 377 (58.0) |
| Ex-smoker | 50 (43.5) | 61 (44.9) | 43 (39.8) | 109 (37.5) | 263 (40.5) |
| Current smoker | 4 (3.5) | 5 (3.7) | 0 (0.0) | 1 (0.3) | 10 (1.5) |
| Missing smoking status n | 16 | 23 | 19 | 59 | 117 |
| Number of comorbidities*** | 2 (1 to 4) | 2 (1 to 3) | 1 (0 to 3) | 1 (0 to 2) | 1 (0 to 3) |
| Cardiovascular | 65 (49.6) | 68 (42.8) | 51 (40.2) | 128 (36.6) | 312 (40.7) |
| Neurology & psychiatry** | 53 (40.5) | 37 (23.3) | 12 (9.4) | 23 (6.6) | 125 (16.3) |
| Respiratory*** | 56 (42.7) | 47 (29.6) | 23 (18.1) | 80 (22.9) | 206 (26.9) |
| Rheumatology** | 25 (19.1) | 16 (10.1) | 15 (11.8) | 27 (7.7) | 83 (10.8) |
| Type II Diabetes | 26 (19.8) | 32 (20.1) | 24 (18.9) | 57 (16.3) | 139 (18.1) |
| Hospital Stay (days) | 12.0 (4.5 to 27.0) | 7.0 (3.0 to 25.5) | 10.0 (5.0 to 17.0) | 8.0 (4.0 to 18.0) | 9.0 (4.0 to 20.0) |
| WHO 3-4 | 27 (20.6) | 38 (23.9) | 22 (17.3) | 67 (19.1) | 154 (20.1) |
| WHO 5 | 33 (25.2) | 53 (33.3) | 56 (44.1) | 132 (37.7) | 274 (35.7) |
| WHO 6 | 21 (16.0) | 24 (15.1) | 20 (15.7) | 61 (17.4) | 126 (16.4) |
| WHO 7 -9 | 50 (38.2) | 44 (27.7) | 29 (22.8) | 90 (25.7) | 213 (27.8) |
| Discharge to review time | 150.0 (104.0 to 191.5) | 155.0 (127.5 to 184.0) | 168.5 (118.8 to 190.2) | 164.0 (124.5 to 189.0) | 159.0 (120.0 to 189.0) |
| PCR COVID +ve | 108 (89.3) | 127 (85.8) | 107 (89.2) | 292 (90.1) | 634 (88.9) |
| Missing PCR test n | 10 | 11 | 7 | 26 | 54 |
| Systemic steroids* | 47 (41.2) | 35 (24.0) | 34 (28.1) | 99 (30.7) | 215 (30.6) |
| *Missing steroids n* | 17 | 13 | 6 | 28 | 64 |
| Antibiotic therapy | 103 (80.5) | 123 (78.3) | 105 (83.3) | 273 (81.2) | 604 (80.9) |
| *Missing antibiotics n* | 3 | 2 | 1 | 14 | 20 |
| Anticoagulation | 39 (32.2) | 35 (23.6) | 37 (30.1) | 117 (36.1) | 228 (31.8) |
| *Missing anti-coagulation n* | 10 | 11 | 4 | 26 | 51 |

\*p<0.05, \*\*p<0.01, \*\*\*p<0.0001. Number (%) unless † = median [IQR], †† = mean [SD]. Column proportions. There was no relationship between other categories of co-morbidities and cluster classification. IMD = Indices of Multiple Deprivation, PCR= Polymerase Chain Reaction, Anticoagulation = therapeutic dose anticoagulation and did not include intermediate dose anticoagulation. WHO = World Health Organisation. Category 3-4 = no continuous supplemental oxygen needed, 5= continuous supplemental oxygen only, 6= Continuous or Bi-level Positive Airway Pressure ventilation or High Flow Nasal O<sub>2</sub>, 7-9 = Invasive Mechanical Ventilation or other organ support

The patient reported outcomes and physical performance were different between the clusters (Table 4). Impairments in lung physiology and biochemical tests used in the assessment of heart failure, renal failure, diabetes and pre-diabetes, were observed in all the clusters, but none were discriminatory across clusters. In the whole group, 11.4% had persistent systemic inflammation measured by a CRP >10mg/L. CRP was weakly correlated with BMI (r=0·247; p<0·001) (Figure SR4) and was higher in the moderate (11·4%), severe (18·0%) and very severe (16·5%) compared to mild (6.0%) cluster.

**Table 4.** Patient reported outcome measures, exercise capacity, lung physiology and biochemical tests stratified by four recovery clusters

|  | Available data | Cluster 1 | Cluster 2 | Cluster 3 | Cluster 4 | Total |
| --- | --- | --- | --- | --- | --- | --- |
| Total N (%) |  | 131 (17.1) | 159 (20.7) | 127 (16.6) | 350 (45.6) | 767 |
| Time to review from Discharge (days) | 767 (100.0) | 150 (104 to 192) | 155 (128 to 184) | 169 (119 to 190) | 164 (126 to 189) | 159 (120 to 189) |
| Persistent symptoms*** | 628 (81.9) | 114 (100.0) | 131 (98.5) | 97 (91.5) | 230 (83.6) | 572 (91.1) |
| Symptom count*** | 628 (81.9) | 20 (16 to 25) | 13 (8 to 17) | 7 (3 to 12) | 5 (2 to 8) | 8 (4 to 16) |
| GAD-7 >8*** | 767 (100.0) | 118 (90.1) | 44 (27.7) | 16 (12.6) | 4 (1.1) | 182 (23.7) |
| PHQ-9 >= 10*** | 767 (100.0) | 128 (97.7) | 60 (37.7) | 9 (7.1) | 1 (0.3) | 198 (25.8) |
| PCL-5 >= 38*** | 767 (100.0) | 79 (60.3) | 9 (5.7) | 2 (1.6) | 0 (0.0) | 90 (11.7) |
| Dyspnoea-12 score*** | 767 (100.0) | 18.2 (9.9) | 6.7 (6.4) | 4.2 (5.3) | 1.6 (2.5) | 5.9 (8.2) |
| FACIT fatigue*** | 767 (100.0) | 34.9 (8.9) | 23.8 (8.6) | 11.2 (8.4) | 7.0 (5.2) | 15.9 (12.9) |
| BPI severity*** | 587 (76.5) | 21.5 (8.4) | 13.9 (8.6) | 11.7 (10.1) | 7.3 (7.6) | 12.1 (9.9) |
| BPI interference*** | 574 (74.8) | 39.9 (17.1) | 21.2 (14.9) | 14.1 (15.8) | 6.7 (9.7) | 17.5 (18.3) |
| SPPB (mobility disability) *** | 767 (100.0) | 94 (71.8) | 72 (45.3) | 67 (52.8) | 101 (28.9) | 334 (43.5) |
| ISWT Distance (m) *** | 463 (60.4) | 278 (190) | 432 (258) | 415 (226) | 536 (262) | 452 (261) |
| CFS >=5 *** | 702 (91.5) | 26 (22.8) | 2 (1.3) | 4 (3.5) | 4 (1.2) | 36 (5.1) |
| MoCA <23*** | 767 (100.0) | 40 (30.5) | 2 (1.3) | 82 (64.6) | 0 | 124 (16.2) |
| Adj MoCA <23*** | 767 (100.0) | 36 (27.5) | 1 (0.6) | 70 (55.1) | 0 | 124 (16.2) |
| FEV<sub>1</sub> % predicted* | 359 (46.8) | 79.8 (20.9) | 91.2 (35.5) | 89.5 (20.8) | 91.6 (17.5) | 89.3 (24.2) |
| FEV<sub>1</sub> <80% predicted** | 359 (46.8) | 27 (26.5) | 23 (22.5) | 18 (17.6) | 34 (33.3) | 102 (100) |
| FVC <80% predicted** | 358 (46.7) | 30 (26.1) | 25 (21.7) | 20 (17.4) | 40 (34.8) | 115 (100) |
| FEV<sub>1</sub>/FVC <0.7 | 403 (52.5) | 3 (4.7) | 10 (11.0) | 8 (11.1) | 12 (6.8) | 33 (8.2) |
| TLCO <80% predicted | 141 (18.4) | 8 (40.0) | 12 (36.4) | 7 (30.4) | 18 (27.7) | 45 (31.9) |
| KCO <80% predicted* | 144 (18.8) | 2 (9.5) | 7 (21.2) | 3 (12.5) | 2 (3.0) | 14 (9.7) |
| BNP/NT-BNP % >threshold§ | 365 (47.6) | 5 (7.9) | 4 (4.9) | 5 (8.5) | 13 (8.0) | 27 (7.4) |
| HbA1C ≥6.0% DCCT/NGSP | 433 (56.5) | 27 (41.5) | 26 (27.4) | 29 (38.2) | 65 (33.0) | 147 (33.9) |
| eGFR < 60 ml/min/1.73 m<sup>2</sup> | 538 (70.1) | 16 (17.2) | 11 (9.5) | 14 (16.7) | 35 (14.3) | 76 (14.1) |
| D-dimer (>= 500 ng/ml) | 464 (60.5) | 8 (10.5) | 14 (13.9) | 11 (14.3) | 30 (14.3) | 63 (13.6) |
| D-Dimer (mg/L) | 455 (59.3) | 270.0 (345.1) | 240.6 (207.1) | 279.2 (290.9) | 293.5 (354.1) | 275.8 (315.4) |
| CRP (>10 mg/L) ** | 499 (65.1) | 15 (16.5) | 20 (18.0) | 9 (11.4) | 13 (6.0) | 57 (11.4) |

\*p<0.05, \*\*p<0.01, \*\*\*p<0.0001. Number (%) unless †median [IQR], †† mean [SD], §Threshold - BNP ≥100ng/L or NT-BNP ≥400ng/L GAD7 = General Anxiety Disorder 7 Questionnaire, PHQ-9 = Patient Health Questionnaire-9, PCL-5 = Post Traumatic Stress Disorder Checklist, Dyspnoea-12 Questionnaire, FACIT Fatigue Scale (FACIT), BPI = Brief Pain Inventory, SPPB = Short Physical Performance Battery, ISWT = Incremental Shuttle Walking Test, CFS = Clinical Frailty Scale, MoCA = Montreal Cognitive Assessment, Adj MoCA – MoCA adjusted for education, FEV<sub>1</sub> = Forced Expiratory Volume in 1 second, FVC = Forced Vital

The patient-perceived recovery was lowest in the very severe (2.7%) and severe (7.0%) severe clusters compared to the moderate (36.4%) and mild (42.7%), p<0.001 (Table 5). The EQ5D-5L VAS and utility index was lowest in the ‘very severe’ cluster 1 pre-COVID with the greatest change between pre-COVID and follow-up in the ‘severe and very severe’ clusters 1 and 2 (p<0.001). There were higher rates of new disability in Cluster 1 (51.8%) compared to 20%, 11.5%, and 4.6% across clusters 2-4, respectively (p<0.001). Cluster 1 also had a greater proportion of people no longer working after hospitalisation with COVID-19 (50.0%) versus 10.0-16.1% across the other clusters (p<0.001), and the greater proportion of people who experienced a change in occupation due to health reasons after COVID-19 (60.0%) versus 8.7-19.4% across the other clusters (p=0.001) (Table SR2c and Table SR2d).

**Table 5.** Change in health-related quality of life and disability after COVID-19 stratified by four recovery clusters

|  | Cluster 1 | Cluster 2 | Cluster 3 | Cluster 4 | Total |
| --- | --- | --- | --- | --- | --- |
| Total N (%) | 131 (17.1) | 159 (20.7) | 127 (16.6) | 350 (45.6) | 767 |
| **Do you feel fully recovered from COVID-19? \*\*\*** | .. | .. | .. | .. | .. |
| Yes^ | 3 (2.7) | 9 (7.0) | 36 (36.4) | 114 (42.7) | 162 (26.6) |
| No^ | 95 (84.1) | 86 (66.7) | 47 (47.5) | 98 (36.7) | 326 (53.6) |
| Not sure^ | 15 (13.3) | 34 (26.4) | 16 (16.2) | 55 (20.6) | 120 (19.7) |
| Missing | 18 | 30 | 28 | 83 | 159 |
| **How good or bad is your health overall (EQ5D-5L VAS 0-100)?** | .. | .. | .. | .. | .. |
| Pre-COVID†† *** | 74 (18) | 80 (15) | 84 (15) | 86 (13) | 81 (17) |
| Post-COVID † | 54 (20) | 69 (16) | 76 (17) | 81 (15) | 72 (20) |
| **EQ5D-5L Utility Index (UI)** | .. | .. | .. | .. | .. |
| EQ5D-5L UI Pre-COVID ¥*** | 0.67 (0.30) | 0.83 (0.19) | 0.87 (0.17) | 0.92 (0.13) | 0.84 (0.23) |
| EQ5D-5L UI Post-COVID ††*** | 0.43 (0.27) | 0.68 (0.17) | 0.76 (0.18) | 0.87 (0.14) | 0.71 (0.26) |
| Change EQ5D-5L UI†††*** | -0.25 (0.36) | -0.17 (0.20) | -0.10 (0.18) | -0.05 (0.15) | -0.11 (0.22) |
| **EQ5D Mobility\*\*\*** | .. | .. | .. | .. | .. |
| No change | 32 (40.0) | 49 (47.6) | 54 (65.9) | 195 (78.0) | 425 (60.8) |
| Improvement | 5 (6.2) | 3 (2.9) | 9 (11.0) | 15 (6.0) | 51 (7.3) |
| Worse | 43 (53.8) | 51 (49.5) | 19 (23.2) | 40 (16.0) | 223 (31.9) |
| **Self-Care\*\*\*** |  |  |  |  |  |
| No change | 20 (25.0) | 41 (39.4) | 54 (65.9) | 192 (77.1) | 392 (56.2) |
| Improvement | 3 (3.8) | 1 (1.0) | 1 (1.2) | 2 (0.8) | 12 (1.7) |
| Worse | 57 (71.2) | 62 (59.6) | 27 (32.9) | 55 (22.1) | 294 (42.1) |
| **Usual Activities\*\*\*** | .. | .. | .. | .. | .. |
| No change | 22 (27.5) | 47 (45.2) | 54 (65.9) | 190 (76.3) | 392 (56.2) |
| Improvement | 8 (10.0) | 3 (2.9) | 4 (4.9) | 12 (4.8) | 46 (6.6) |
| Worse | 50 (62.5) | 54 (51.9) | 24 (29.3) | 47 (18.9) | 260 (37.2) |
| **Pain/ Discomfort\*\*\*** | .. | .. | .. | .. | .. |
| No change | 33 (41.2) | 45 (43.3) | 40 (49.4) | 166 (66.7) | 361 (51.8) |
| Improvement | 14 (17.5) | 23 (22.1) | 23 (28.4) | 48 (19.3) | 151 (21.7) |
| Worse | 33 (41.2) | 36 (34.6) | 18 (22.2) | 35 (14.1) | 185 (26.5) |
| **Anxiety/Depression \*\*\*** | .. | .. | .. | .. | .. |
| No change | 9 (11.2) | 37 (35.9) | 44 (53.7) | 175 (70.0) | 342 (49.0) |
| Improvement | 20 (25.0) | 13 (12.6) | 11 (13.4) | 25 (10.0) | 86 (12.3) |
| Worse | 51 (63.8) | 53 (51.5) | 27 (32.9) | 50 (20.0) | 270 (38.7) |
| ‘alot of difficulty’ | 63 (55.8) | 33 (26.2) | 15 (15.0) | 18 (6.8) | 204 (24.7) |
| ***§ |  |  |  |  |  |
| 'new disability'***§ | 57 (51.8) | 25 (20.0) | 11 (11.5) | 12 (4.6) | 158 (19.6) |

\*p<0.05, \*\*p<0.01, \*\*\*p<0.0001. Number (%) unless †median [IQR] or ††mean [SD], ¶ = % of category with positive response, § Washington Group Short Set Functioning (WGSS) – 'a lot of difficulty' or 'cannot do at all' score for any of the seven problems, 'new disability' a new score of 'a lot of difficulty or 'cannot do at all' persisting after COVID-19, ^ % calculated after exclusion of missing individuals, EQ5D-5L VAS = Visual Analogue Scale, EQ5D-5L UI = Utility Index

## Discussion

This is the largest study to report in detail on prospectively assessed outcomes across multiple UK centres to describe the impact of COVID-19 on medium term health of survivors. The majority had not fully recovered, had persistent symptoms and 20% had a new disability. In the two-thirds that were working prior to admission, 19% had changed working status predominately due to ill health. Failure to fully-recover was associated with female sex, white ethnicity, middle age, two or more co-morbidities, and more severe acute illness. Treatment with systemic corticosteroids was not associated with recovery. The magnitude of the ongoing mental and physical health burden was substantial, but perhaps surprisingly were largely unrelated to acute severity. Cluster analysis determined by patient reported outcomes and tests of physical performance, depression, anxiety, cognition, breathlessness and fatigue, identified four recovery clusters that tracked with severity of both mental and physical health impairment, except for poor cognition that was largely independent. Whether these clusters have different underlying mechanisms, and warrant different treatments and clinical pathways needs to be determined.

We have undertaken a comprehensive and holistic assessment of patients post-hospitalisation for COVID-19. Patients rated their perceived recovery, current symptoms, health status and disability compared to their status prior to admission. The majority of patients had not recovered over five months and this patient-perceived recovery was consistent with the proportion that had persistent symptoms and worsening in health status. Recovery at five months following hospitalisation for COVID-19 of 20-30% is consistent with previous reports^10, 30^ and for those mechanically ventilated is similar to prior studies of intensive care survivorship.^31^ In contrast, in the community recovery following COVID-19 is 70-90% in most studies at 3 months.^32^ This does suggest that severity of the acute illness warranting hospitalisation is associated with a lower likelihood of recovery.

Here we describe features that were associated with failure to achieve patient-perceived recovery. Age had a non-linear relationship with recovery with those that were younger or older having a higher likelihood of recovery whereas poor recovery was associated with middle age. Consistent with a previous report for community managed COVID-19, which found that persistent symptoms were more common in women,^32^ we found that female sex was associated with failure to recover. Autoimmunity is more common in women over forty years old and has been implicated in post-COVID syndromes^33^; whether this is one possible explanation of this association needs to be further investigated. Intriguingly, we found that white ethnicity was associated with failure to recover in contrast to the consistent greater morbidity and mortality following the acute COVID-19 infection in ethnic minorities^34, 35^ and likewise increased risk post-discharge of cardiometabolic and respiratory events in this group.^36^ Two or more co-morbidities were associated with both increased risk of severe acute illness and subsequent poor recovery post-hospitalisation. Severity of the acute illness specifically mechanical ventilation and additional organ support was associated with patient-perceived poor recovery in keeping with similar findings following ITU-survivorship.^3^ Systemic corticosteroid therapy for the acute illness reduces mortality in those with more severe acute disease,^37^ but was not associated with post-discharge medium term recovery nor was acute treatment with antibiotics.

The physical, cognitive and mental health burden experienced by COVID-19 survivors was considerable. This included symptoms of anxiety and depression in a quarter, post-traumatic stress disorder in 12%, cognitive impairment in 17%, reduced exercise capacity and functional performance in 35-50%, impaired lung function and elevated HbA1c each in approximately a third, abnormal renal function in 13% and increased BNP in 7%. Lung diffusion was also abnormal in one-third of patients although this was undertaken in a minority of subjects. Taken together the restrictive spirometry with reduced transfer factor, but relatively preserved transfer coefficient is suggestive of predominately discrete parenchymal loss rather than diffuse fibrosis or pulmonary vascular disease. Importantly, the pattern and magnitude of the ongoing burden on mental and physical health was largely unrelated to the severity of the acute illness. This led us to define further possible recovery phenotypes using cluster analysis including validated tools for the domains breathlessness, fatigue, mental health, cognition and physical function.

We describe here four clusters that showed impairment in mental and physical health were related with clusters reflecting their severity, whereas cognitive impairment was largely distinct. The ‘mild’ cluster had the largest number of patients and the most that felt recovered. The numbers of patients in each of the other clusters moderate, severe and very severe were similar. In cluster one ‘very severe’ and cluster two ‘severe’ a small minority felt recovered. The number of co-morbidities were increased in these clusters compared to the ‘moderate’ and ‘mild’ clusters. Similarly, BMI was also higher in clusters one and two compared to clusters three and four. This suggests that in COVID-19 obesity is both associated with morbidity and mortality during the acute-phase of infection, and greater physical and mental health impact longer term. Cognitive impairment was a particular feature of cluster three even after taking into consideration education level. Impaired cognition is reported in those requiring mechanical ventilation for critical illness.^38^ Intriguingly, the clusters were not closely related to acute severity in our study further supporting the view that severity of persistent physical and mental ill health and poor cognition are due to mechanisms largely independent of mechanical ventilation. Pre-COVID-19 poor health status and comorbidities were particularly a feature of cluster one and to a lesser extent cluster two compared to clusters three and four. However, change in EQ5D-5L VAS and utility index showed the greatest impact on health status was in cluster one and two even accounting for poorer health status pre-COVID-19. Interestingly, even though there were abnormalities in physiological and biochemical tests of respiratory, cardiac and renal function, diabetes and prediabetes within each cluster, these objective tests were not discriminatory across the clusters except for forced expiratory volume in one-second percent predicted (FEV_1_ %) and CRP levels. The FEV_1_ % was lower in the very severe group without evidence of airflow obstruction suggestive of airflow restriction possibly due to lung fibrosis or in part due to extra-thoracic restriction secondary to obesity. The CRP was particularly elevated in the severe and very severe clusters and to a lesser extent in the moderate group compared to the mild cluster possibly due to post-COVID-19 systemic inflammation. The positive association between CRP and BMI is well-established. However, we found only a weak correlation between these parameters suggesting that although the elevated CRP in clusters 1-3 might be partly due to increased BMI that this is unlikely to fully explain the increased systemic inflammation. Persistent systemic inflammation is associated with poor physical recovery after critical illness. Therefore, the magnitude of the physical and mental health impact, the heterogeneity of the poor cognition between and within these clusters, and the impact of persistent inflammation and its impact on the immunological response require more understanding of possible underlying mechanisms.

Beyond the impact on health, 68% of participants were working prior to hospital admission. For 1-in-5 patients working status changed and a similar proportion experienced a health-related change in occupation. This impact on occupation was most marked in the group that had required mechanical ventilation and was similar to previous reports in ITU-survivorship studies.^39^ In the recovery clusters, impact on occupation was most striking in cluster one with over 60% working prior to COVID-19 infection and now ∼50% having changed occupation almost entirely due to poor health. This societal impact is clearly substantial in those hospitalised but also highly relevant for non-hospitalised cases of COVID-19 infection.

This study has a number of limitations. The patients represent a small sample of the total number of patients discharged from hospital following COVID-19 infection in the UK. The study population is younger than the whole population in the UK that survived hospitalisation for COVID-19 infection^40^ and only included those able to attend clinic visits and undertake the study procedures. This acquisition bias might under-represent the most severely affected patients but conversely those patients with ongoing symptoms might have been more willing to participate. The patient-reported outcomes, physical and biological tests assessed cross-sectionally does not allow for the clear differentiation between the contribution from premorbid disease versus emergent impaired health status and symptoms. Further analysis of the trajectory of recovery and linkage to primary and secondary health records within PHOSP-COVID will enable further discrimination. Notwithstanding this limitation, the magnitude of burden of physical and mental health is substantial in this group. Our definition for recovery in this report is a subjective definition based on patient perception and will fail to identify pathological changes that have not yet led to clinical expression, but might become overt in later follow-up. However, patient-perceived recovery did correspond well to the overall burden of disease identified in the recovery phenotypes. Further comparisons are also required with recovery following acute respiratory infections leading to hospitalisation and with those infected with COVID-19 but not hospitalised to understand the specificity of our findings to COVID versus other critical illnesses and between those hospitalised versus managed in the community.

The present report is the first from the PHOSP-COVID study, which includes biosampling to enable further immunology, and multi-omics and imaging endpoints including multi-modality magnetic resonance imaging. These will enable further careful analysis of systemic and organ-specific inflammation and possible organ damage. Further study of the trajectory of recovery coupled with this greater mechanistic understanding will further inform a precision medicine approach to the clinical management of hospitalised COVID-19survivors.

In conclusion, the majority of survivors of a hospital admission with COVID-19 are not fully recovered at five months and have substantial diverse physical and mental health burden and negative impact on employment. We identified key factors associated with recovery and four distinct recovery phenotypes using cluster analysis according to mental, cognitive, and physical health. Our findings support the need for a proactive approach to clinical follow-up with a holistic assessment to include symptoms of mental and physical health, and validated assessment of cognition. The four severity clusters highlight potential to stratify care, and also the need for wide-access to COVID-19 holistic clinical services to include mental health, memory and cognition, and rehabilitation services.

## Data Availability

The protocol, consent form, definition and derivation of clinical characteristics and outcomes, training materials, regulatory documents, requests for data access and other relevant study materials are available online at www.phosp.org.

## Supplement – Tables and Figures

### Supplement Tables

**Table SM1.** Description of the outcome measures

**Table SM2.** Methodology for the outcome measures including conduct

**Table SR1.** Co-morbidities for the cohort stratified by severity of acute illness using the WHO clinical progression scale.

**Table SR2.** Occupation data at baseline, and change in employment by severity of acute illness and by cluster severity

**Table SR3**. Patient reported outcomes, physiological and biochemical tests stratified by severity of acute illness.

**Table SR4.** Comparison between imputed and non-imputed logistic regression of predictors of failure to recover (multi-variable and multi-level).

**Table SR5.** Ongoing symptoms recorded at follow-up for the cohort stratified between those with and without pre-existing co-morbidities.

**Table SR6.** Proportion unchanged, worse or better in terms of a) Symptoms, b) Health-related quality of life (EQ5D-5L) c) Disability (WG-SS) at follow-up compared to prior to hospitalisation stratified by severity of acute illness.

### Supplement Figures

**Figure SR1.** Consort flow diagram for participants

**Figure SR2.** Histogram of number of symptoms reported at five months after discharge in survivors of a hospital admission due to COVID-19 Change in disability (WG-SS) and symptoms stratified by severity of illness

**Figure SR3.** Clusters of Patient reported outcome measures and physical function

**Figure SR4.** Correlation of C-Reactive Protein Level and Body Mass Index by clusters

## Writing Group (on behalf of the PHOSP-COVID Collaborative Group)

Rachael A Evans*, Hamish McAuley, Ewen M Harrison, Aarti Shikotra, Amisha Singapuri, Marco Sereno, Omer Elneima, Annemarie B Docherty, Nazir I Lone, Olivia C Leavy, Luke Daines, J Kenneth Baillie, Jeremy S Brown, Trudie Chalder, Anthony De Soyza, Nawar Diar Bakerly, Nicholas Easom, John R Geddes, Neil J Greening, Nicholas Hart, Liam G Heaney, Simon Heller, Luke Howard, Joesph Jacob, R Gisli Jenkins, Caroline Jolley, Steven Kerr, Onn Min Kon, Keir Lewis, Janet M Lord, Gerry P McCann, Stefan Neubauer, Peter J M Openshaw, Paul Pfeffer, Matthew Rowland, Malcolm G Semple, Sally J Singh, Aziz Sheikh, David Thomas, Mark Toshner, James D Chalmers, Ling Pei Ho, Alex Horsley, Michael Marks, Krisnah Poinasamy, Louise V Wain* Christopher E Brightling*,

*Joint first and last authors and contributed equally

## Contributors

The manuscript was initially drafted by CEB, RAE, LVW further developed by the writing committee. CEB, RAE, LVW, OE, HM, AShi, ADS, RGJ, LH MR, JRG, GPM, DT, SJS, JML, SN, JJ, MGS, AH, OMK, NH, MT, PEP, LPH, JDC, MM, JKB, ABD, EMH, ASh, NIL, NG, LGH, KEL made substantial contributions to the conception and design of the work. RAE, AH, OMK, CJ, NH, MT, JB, NE, PEP, NDB, LPH, JDC, MM, ASi, made substantial contributions to the acquisition of data. CEB, RAE, LVW, HM, OE, EMH, ABD, JKB, Ash, LD, SK, NIL, MS, OCL, AH, LPH, JDC, MM, KP, NG, LGH, KEL, ADS, RGJ, LH, MR, JRG, GPM, SH, DT, PJMO, SJS, JML, TC, SN, JJ, MGS, KP, made contributions to the analysis, or interpretation of data for the work. All authors contributed to data interpretation and critical review and revision of the manuscript. Final approval of the version to be published and agreement to be accountable for all aspects of the work in ensuring that questions related to the accuracy or integrity of any part of the work are appropriately investigated and resolved.

## Acknowledgments

This study would not be possible without all the participants who have given their time and support. We thank all the participants and their families. We thank the many research administrators, health-care and social-care professionals who contributed to setting up and delivering the study at all of the 40 NHS Trusts and 25 Research Institutions across the UK as well as all the supporting staff at the at the NIHR Clinical Research Network, Health Research Authority, Research Ethics Committee, Department of Health and Social Care Public Health Scotland, Public Health England and support from the ISARIC Coronavirus Clinical Characterisation Consortium (ISARIC4C). At the NIHR Office for Clinical Research Infrastructure (NOCRI) we thank Kate Holmes (for her support in coordinating the charities group) Ivana Poparic and Peter Sargent and Sheuli Porkess at the Association of the British Pharmaceutical Industry for their advice in commercial discussions. We are very grateful to all the charities that have provided insight to the study-Action Pulmonary Fibrosis, Alzheimer’s Research UK, Asthma UK /British Lung Foundation UK/BLF, British Heart Foundation, Diabetes UK, Cystic Fibrosis Trust, Kidney Research UK, MQ Mental Health, Muscular Dystrophy UK, Stroke Association Blood Cancer UK, McPin Foundations, Versus Arthritis. We thank the NIHR Leicester Biomedical Research Centre patient and public involvement group and the Long Covid Support Group.

