## Supplementary Appendix for "Physical, cognitive and mental health impacts of COVID-19 following hospitalisation – a multi-centre prospective cohort study"

#### Contents

Details of the PHOSP-COVID Collaborative Group ........ ii

Study Organisation ........ ix

Appendix 1- PHOSP-COVID Study Protocol Version 5 24 Nov 2020 ........ x

#### Details of the PHOSP-COVID Collaborative Group

##### Writing Committee

R A Evans*, H McAuley, E M Harrison, A Shikotra, A Singapuri, M Sereno, O Elneima, A B Docherty, N I Lone, O C Leavy, L Daines, J K Baillie, J S Brown, T Chalder, A De Soyza, N Diar Bakerly, N Easom, J R Geddes, N J Greening, N Hart, L G Heaney, S Heller, L Howard, J Jacob, R G Jenkins, C Jolley, S Kerr, O M Kon, K Lewis, J M Lord, G P McCann, S Neubauer, P J M Openshaw, P Pfeffer, M Rowland, M G Semple, S J Singh, A Sheikh, D Thomas, M Toshner, J D Chalmers, L P Ho, A Horsley, M Marks, K Poinasamy, L V Wain* C E Brightling*,

##### Steering Committee

*Co-chairs* D Lomas, E Sapey, *Institution representatives* C Berry, C E Bolton, N Brunskill, E R Chilvers, R Djukanovic, Y Ellis, D Forton, N French, J George, N A Hanley, N Hart, L McGarvey, N Maskell, H McShane, M Parkes, D Peckham, P Pfeffer, A Sayer, A Sheikh, R Thompson, N Williams and core management group representation

##### Executive Board

*Chair* C E Brightling, representation from the core management group, each working group and platforms

##### Core Management Group

*Chief Investigator* C E Brightling, *Members* J D Chalmers, R A Evans (Lead Co-I), L V Wain (Lead Co-I), L P Ho, A Horsley, M Marks, K Poinasamy, A Singapuri, A Shikotra,

#### Working Groups

##### Airways

L G Heaney (*Co-Lead*), A De Soyza (*Co-Lead*), D Adeloye, C E Brightling, J S Brown, J Busby, J D Chalmers, L Daines, O Elneima, J Hurst, P Novotny, P Pfeffer, K Poinasamy, J Quint, I Rudan, E Sapey, A Sheikh, S Siddiqui, S Walker

##### Brain

M Hotopf *(Co-Lead),* J R Geddes (*Co-Lead*), K Abel, R Ahmed, L Allan, C Armour, D Baguley, D Baldwin, C Ballard, J Bambrough, K Bhui, G Breen, M Broome, T Brugha, E Bullmore, D Burn, J Cavanagh, T Chalder, D Clark, A David, B Deakin, H Dobson, B Elliott, J Evans, R Francis, E Guthrie, P Harrison, M Henderson, A Hosseini, N Huneke, M Husain, T Jackson, I Jones, T Kabir, P Kitterick, A Korszun, I Koychev, J Kwan, A Lingford-Hughes, C Mackay, P Mansoori, H McAllister-Williams, K McIvor, B Michael, L Milligan, R Morriss, E Mukaetova-Ladinska, T Nicholson, S Paddick, C Pariante, J Pimm, K Saunders, M Sharpe, G Simons, R Upthegrove, S Wessely

##### Cardiac

G P McCann (*Co-Lead*), C Antoniades, R Bell, A Bularga, C Berry, K Channon, P Chowienczyk, J Greenwood, A Hingorani, A Hughes, K Khunti, C Kotanidis, J Mayet, N Mills, A J Moss, S Neubauer, D Newby, B Raman, N Samani, A Saratzis, N Sattar, A Shah, C Sudlow, M Toshner, R Touyz, B Williams, C Xie

**Immunology**

T Hussell (*Co-Lead*), P J M Openshaw (*Co-Lead*), D Altmann, J K Baillie, R Batterham, H Baxendale, N Bishop, C E Brightling, P C Calder, R A Evans, J L Heeney, P Klenerman, J M Lord, P Moss, S L Rowland-Jones, W Schwaeble, M G Semple, R S Thwaites, L Turtle, L V Wain, S Walmsley, D Wraith

**Intensive Care**

M J Rowland (*Co-Lead*), A Rostron (Co-Lead), J K Baillie, B Connolly, A B Docherty, N I Lone, D F McAuley, D Parekh, J Simpson, C Summers

**Lung Fibrosis**

R G Jenkins (*Co-Lead*), J Porter (*Co-Lead*), R Allen, R Aul, J K Baillie, S Barratt, P Beirne, J Blaikley, R Chambers, N Chaudhuri, C Coleman, L Fabbri, P M George, M Gibbons, F Gleeson, B Gooptu, I Hall, N A Hanley, L P Ho, E Hufton, J Jacob, S Johnson, M Jones, S Jones, F Khan, J Mitchell, P L Molyneaux, P R Ortega, J E Pearl, K Piper Hanley, K Poinasamy, J Quint, M G Semple, J Simpson, M Spears, L Spencer, S Stanel, I Stewart, D Thickett, R Thwaites, L V Wain, S Walker, S Walsh, J Wild, D Wootton, L Wright

**Metabolic**

S Heller (*Co-Lead*), H Atkins, S Bain, M Davies, J Dennis, K Ismail, D Johnston, P Kar, K Khunti, C Langenberg, P McArdle, A McGovern, T Peto, J Petrie, E Robertson, N Sattar, K Shah, J Valabhji, B Young

**Pulmonary and Systematic Vasculature**

L Howard (*Co-Lead*), Mark Toshner (*Co-Lead*), C Berry, P Chowienczyk, D Lasserson, A Lawrie, O C Leavy, J Mitchell, L Price, J Quint, J Rossdale, N Sattar, C Sudlow, R Thompson, J M Wild, M Wilkins

**Rehabilitation, Sarcopenia and Fatigue**

T Chalder (*Co-Lead*), J M Lord (*Co-Lead*), N J Greening (*Co-Lead*), W Man (*Co-Lead*), S J Singh (*Co-Lead*), N Armstrong, E Baldry, N Basu, M Beadsworth, L Bishop, C E Bolton, A Briggs, M Buch, G Carson, J Cavanagh, H Chinoy, E Daynes, S Defres, R A Evans, P Greenhaff, S Greenwood, M Harvie, M Husain, A McArdle, A McMahon, M McNarry, C Nolan, J Pimm, J Sargent, J Scott, L Sigfrid, M Steiner, D Stensel, A L Tan, J Whitney, D Wilkinson, D Wilson, M Witham, T Yates

**Renal**

N Brunskill (*Co-Lead*), S Francis (*Co-Lead*), S Greenwood (*Co-Lead*), C Laing (*Co-Lead*), D Thomas (*Co-Lead*), K Bramham, P Chowdhury, A Frankel, L Lightstone, S McAdoo, K McCafferty, M Ostermann, N Selby, C Sharpe, M Willicombe

#### Platforms

**Bioresource**

W Greenhalf (*Co-Lead*), M G Semple (*Co-Lead*), M Ashworth, H Hardwick, M Sereno, R Saunders, A Singapuri, V Shaw, A Shikotra, L V Wain

**Data Hub**

J K Baillie (*Co-Lead*), A B Docherty (*Co-Lead*), E M Harrison (*Co-Lead*), A Sheikh (*Co-Lead*), C E Brightling, L Daines, S Dunn, R A Evans, S Kerr, O C Leavy, N I Lone, H McAuley, R Pius, M Richardson, M Sereno, L V Wain

**Imaging Alliance**

C Bloomfield (*Co-Lead*), M Halling-Brown (*Co-Lead*), F Gleeson (*Co-Lead*), J Jacob (*Co-Lead*), S Neubauer (*Co-Lead*) B Raman (*Co-Lead*) S Siddiqui (*Co-Lead*) J Wild (*Co-Lead*), P Jezzard, H Lamlum, E Tunnicliffe

**Omics**

L V Wain (*Co-Lead)*, J K Baillie (*Co-Lead*), H Baxendale, C E Brightling, M Brown, J D Chalmers, R A Evans, B Gooptu, W Greenhalf, H Hardwick, R G Jenkins, D Jones, I Koychev, C Langenberg, A Lawrie, P Molyneaux, A Shikotra, J Pearl, M Ralser, N Sattar, R Saunders, J Scott, T Shaw, D Thomas, D Wilkinson

**PHOSP-COVID Study Central Coordinating Team**

C E Brightling (Chief Investigator), R A Evans (*Lead Co-I*), L V Wain (*Lead Co-I*), S Diver, R Dowling, C Edwardson, O Elneima, S Finney, R Free, N J Greening, B Hargadon, V Harris, L Houchen, W Ibrahim, O C Leavy, A Ient, H McAuley, P Novotny, C Overton, T Plekhanova, R Saunders, M Sereno, A Singapuri, M Sharma, A Shikotra, C Taylor, S Terry, E Turner, C Tong, A J Yousuf, B Zhao

#### Local Clinical Centre PHOSP-COVID trial staff

(listed in descending order of the number of Tier 2 participants recruited per Trust at the 1000 Tier 2 timepoint)

**1. University Hospitals of Leicester NHS Trust & University of Leicester**

C E Brightling (CI), R A Evans (PI), M Aljaroof, N Armstrong, H Arnold, M Bakali, M Bakau, M Bourne, C Bourne, N Brunskill, P Cairns, L Carr, A Charalambou, C Christie, M Davies, S Diver, S Edwards, C Edwardson, O Elneima, H Evans, J Finch, S Glover, N Goodman, B Gootpu, N J Greening, K Hadley, P Haldar, B Hargadon, L Houchen, W Ibrahim, L Ingram, K Khunti, A Lea, D Lee, G P McCann, H McCauley, P McCourt, T Mcnally, A Moss, W Monteiro, M Pareek, S Parker, A Rowland, A Prickett, R Russell, J Skeemer, S Siddiqui, S J Singh, M Soares, E Stringer, T Thornton, M Tobin, L V Wain, F Woodhead, T Yates, A Yousuf

**2. Manchester University NHS Foundation Trust & University of Manchester**

B Al-Sheklly (PI), A Horsley (PI), C Avram, J Blaikely, M Buch, N Choudhury, D Faluyi, T Felton, T Gorsuch, N A Hanley, T Hussell, Z Kausar, N Odell, R Osbourne, K Piper Hanley

**3. Imperial College Healthcare NHS Trust & Imperial College London**

L Howard (PI), O Kon (PI), D C Thomas (PI), B Card, E Calvelo, E R Chilvers, P Cullinan, R G Jenkins, C King, M Mariveles, U Munawar, J Nunag, R Parvin, B Pathmanathan, E Russell

**4. King's College Hospital NHS Foundation Trust & Kings College London**

A Shah (PI), C J Jolley (PI), O Adeyemi, H Assefa-Kebede, J Breeze, M Brown, S Byrne, T Chalder, P Dulawan, N Hart, A Hayday, A Hoare, A Knighton, M Malim, S Patale, I Peralta, N Powell, A Ramos, A Shah, K Shevket, F Speranza, A Te

**5. Guy's and St Thomas' NHS Foundation Trust**

N Hart (PI), F Adeyemi, G Arbane, S Betts, A Dewar, G Kaltsakas

**6. Cambridge University NHS Foundation Trust & University of Cambridge**

J Fuld (PI), I Cruz, K Dempsey, A Elmer, H Jones, S Jose, S Marciniak, M Parkes, M Toshner, L Watson, J Worsley

**7. University College London Hospital & University College London**

M Marks (PI), J S Brown (PI), R Chambers, A Checkley, R Evans, M Heightman, T Hillman, J Hurst, J Jacob, S Janes, M Lipman, S Logan, D Lomas, J Porter, K Roy, E Wall

**8. Hull University Teaching Hospitals NHS Trust & University of Hull**

N Easom (PI), P Atkin, K Brindle, M G Crooks, R Flockton, L Holdsworth, A Richards, D L Sykes, S Thackray-Nocera, K Vellore, C Wright

**9. Barts Health NHS Trust & Queen Mary University of London**

P Pfeffer (PI), K Chong-James, C David, W Y James, A Martineau, O Zongo

**10. Royal Free London NHS Foundation Trust**

J Hurst (PI), H Jarvis (PI), S Mandal (PI), S Ahmad, S Brill, L Lim, D Matila, E Murali, O Olaosebikan, C Singh

**11. Salford Royal NHS Foundation Trust**

N Diar Bakerly (PI), A Atkins, P Dark, D Evans, E Gourlay, E Hardy, A Harvey, D Holgate, S Knight, N Mairs, N Majeed, L McMorrow, J Oxton, J Pendlebury, C Summersgill, R Ugwuoke, S Whittaker

**12. Oxford University Hospitals NHS Foundation Trust & University of Oxford**

L P Ho (PI), N M Rahman (PI), M Ainsworth, A Alamoudi, A Bloss, P Carter, J Chen, T Dong, R I Evans, E Fraser, J R Geddes, F Gleeson, P Harrison, M Havinden-Williams, P Jezzard, I Koychev, P Kurupati, H McShane, S Neubauer, D Nicoll, G Ogg, E Pacpaco, M Pavlides, Y Peng, N Petousi, N Rahman, B Raman, M J Rowland, K Saunders, M Sharpe, N Talbot, E Tunnicliffe

**13. University Hospital Birmingham NHS Foundation Trust & University of Birmingham**

D Parekh (PI), R Baggott, A Botkai, P Clift, B Cooper, J Dasgin, N Gautam, N Ghatum, T Hiwot, T Jackson, S Johnson, V Kamwa, J M Lord, S Madathil, A Newton Cox, J Nyaboko, H Qureshi, E Sapey, J Short, J Stockley, Z Suleiman, T Thompson, S Walder, C Welch, D Wilson, S Yasmin, K P Yip

**14. Newcastle upon Tyne Hospitals NHS Foundation Trust & University of Newcastle**

A De Soyza (PI), C Echevarria (PI), J Brown, G Burns, G Davies, H Fisher, C Francis, A Greenhalgh, P Hogarth, J Hughes, K Jiwa, G Jones, G MacGowan, D Price, A Sayer, J Simpson, H Tedd, S West, M Witham, S Wright, A Young

**15. University Hospital Southampton NHS Foundation Trust & University of Southampton**

M Jones (PI), R Djukanovic, S Fletcher, M Harvey, B Marshall, R Samuel, T Sass, T Wallis, H Wheeler

**16. Nottingham University Hospitals NHS Trust & University of Nottingham**

C E Bolton (PI), J Bonnington, C Dobson, P Greenhaff, A Gupta, L Howard, S Linford, L Matthews, A K Thomas

**17. St George's University Hospitals NHS Foundation Trust**

R Aul (PI), M Ali, A Dunleavy, D Forton, N Msimanga, M Mencias, S Siddique, V Tavoukjian

**18. Liverpool University Hospitals NHS Foundation Trust & University of Liverpool**

D G Wootton (PI), L Turtle (PI), J Brown, A Cross, S L Dobson, N French, W Greenhalf, H Hardwick, K Hainey, J Hawkes, AL Key, N Lewis-Burke, G Madzamba, M J Noonan, L Poll, M G Semple, V Shaw, K A Tripp, L O Wajero, S A Williams-Howard

**19. Sheffield Teaching NHS Foundation Trust & University of Sheffield**

S Rowland-Jones (PI), A Alli, A Angyal, L Armstrong, M Bayley, E Bradley, R Brown, K Chapman, L Chetham, C Clark, Z Coburn, J Cole, T De Silva, M Dixon, A Fairman, D Foote, A Ford, R Gregory, K Harrington, S Heller, L Hesselden, A Holbourn, B Holroyd-Hind, A Howell, A Lawrie, E Lee, R Lenagh, I Macharia, S Megson, J Meiring, H Newell, J Rodgers, I Smith, J Smith, L Smith, R Thompson, H Turton, J Watson, L Watson, J Wild, I Wilson, A Zawia

**20. Belfast Health and Social Care Trust & Queen's University Belfast**

L G Heaney (PI), C Armour, V Brown, T Craig, S Drain, B King, N Magee, D McAulay, E Major, L McGarvey, J McGinness, R Stone

**21. NHS Tayside & University of Dundee**

J D Chalmers (PI), D Connell, J George, C J Tee, J Rowland, D Sutherland, A Elliott

**22. Aneurin Bevan University Health Board**

S Fairbairn (PI), A Dell, N Hawkings, J Haworth, M Hoare, A Lucey, V Lewis, G Mallison, H Nassa, C Pennington, A Price, C Price, A Storrie, G Willis, S Young

**23. Royal Brompton and Harefield NHS Foundation Trust**

W Man (PI), B Patel (PI), R E Barker, D Cristiano, N Dormand, M Gummadi, S Kon, K Liyanage, C M Nolan, S Patel, O Polgar, P Shah, S J Singh, J A Walsh

**24. NHS Lothian & University of Edinburgh**

G Choudhury (PI), J K Baillie, S Clohisey, A Deans, A B Docherty, J Furniss, E M Harrison, S Kelly, N I Lone, A Sheikh

**25. Hywel Dda University Health Board**

M Andrews (PI), K E Lewis (PI), A Mohamed (PI), G Ross (PI), K Davies, R Hughes, R Loosley, L O'Brien, Z Omar, H McGuinness, E Perkins, J Phipps, A Taylor, H Tench, R Wolf-Roberts

**26. Royal Papworth Hospital NHS Foundation Trust**

M Toshner (PI), H Baxendale, L Garner, C Johnson, A Michael, H Parfrey, J Parmar

**27. Bradford Teaching Hospitals NHS Foundation Trust**

D Saralaya (PI), L Brear, K Regan, K Storton

**28. North Bristol NHS Trust & University of Bristol**

N Maskell (PI), D Arnold, S Barrett, H Adamali, A Dipper, S Dunn, A Morley, L Morrison, L Stadon, H Welch

**29. Betsi Cadwaladr University Health Board**

A Haggar (PI), A Bolger, F Davies, J Lewis, A Lloyd, E McIvor, D Menzies, W Saxon, D Southern, C Subbe, V Whitehead

**30. Cardiff and Vale University Health Board**

R Sabit (PI), L Broad, A Buttress, T Evans, L Knibbs, A McQueen, C Oliver, K Paradowski, J Williams

**31. Leeds Teaching Hospitals & University of Leeds**

P Beirne (PI), A Ashworth, J Clarke, C Coupland, M Dalton, E Wade, C Favager, J Greenwood, J Glossop, L Hall, A Humphries, J Murira, D Peckham, S Plein, J Rangeley, A L Tan, B Whittam, N Window, J Woods

**32. London North West University Healthcare NHS Trust**

S N Diwanji (PI), P Papineni (PI), S Gurram, S Quaid, G F Toingson, E Watson

**33. NHS Dumfries and Galloway**

M J McMahon (PI), P Neill

**34. NHS Greater Glasgow and Clyde Health Board & University of Glasgow**

D Anderson (PI), H Bayes (PI), C Berry (PI), D Grieve (PI), I B McInnes (PI), N Basu, A Brown, A Dougherty, K Fallon, L Gilmour, K Mangion, A Morrow, K Scott, R Sykes

**35. NHS Highland**

E K Sage (PI), F Barrett, A Donaldson

**36. NHS Lanarkshire**

M Patel (PI), D Bell, A Brown, M Brown, R Hamil, K Leitch, L Macliver, J Quigley, A Smith, B Welsh

**37. North Middlesex Hospital NHS Trust**

B Jayaraman (PI), T Light

**38. Swansea Bay University Health Board**

G A Davies (PI), L Connor, A Cook, T Rees, F Thaivalappil

**39. The Hillingdon Hospitals NHS Foundation Trust**

S S Kon (PI), H Lota, G Landers, S Portukhay

**40. Whittington Health NHS**

R Dharmagunawardena (PI), E Bright, P Crisp, M Stern

**Health and Care Research Wales**
Y Ellis

**London School of Hygiene & Tropical Medicine (LSHTM)**

M Marks, A Briggs

**NIHR Office for Clinical Research Infrastructure**
K Holmes

**Patient Public Involvement Leads**
Asthma UK and British Lung Foundation Partnership - K Poinasamy, S Walker

**Royal Surrey NHS Foundation Trust**

M Halling-Brown

**South London and Maudsley NHS Foundation Trust & Kings College London**

G Breen, M Hotopf

**Swansea University & Swansea Welsh Network**
K Lewis, N Williams

#### Study Organisation

The PHOSP-COVID Study was set up to provide researchers with a comprehensive understanding of the impact on the health of those that have been hospitalised with COVID-19. The full protocol can be viewed www.phosp.org and is provided in appendix 1. The study was conducted at 40 National Health Service (NHS) Trusts in the United Kingdom. The study was coordinated centrally by the team at University of Leicester, the trial sponsor. Support for local site activities was provided by the National Institute for Health Research Clinical Research Network.

**Appendix 1- PHOSP-COVID Study Protocol Version 5 24 Nov 2020**

Full Study Title:

### Post-hospitalisation COVID-19 study: a national consortium to understand and improve long-term health outcomes

**Short title**: PHOSP-COVID Study

Sponsor Reference No: 0785

Ethics Ref: 20/YH/0225

Date and Version No: Version 5, 24-November-2020

| | |
|---|---|
| Chief Investigator: | Professor Christopher Brightling |
| Investigators: | Dr Rachael Evans and Professor Louise Wain on behalf of the PHOSP-COVID Study consortium |
| Sponsor: | University of Leicester |
| Funder: | NIHR UK Research and Innovation |
| Signatures: | The approved protocol should be signed by author(s) and/or person(s) authorised to sign the protocol |

Confidentiality Statement

All information contained within this protocol is regarded as, and must be kept, confidential. No part of it may be disclosed by any Receiving Party to any Third Party, at any time, or in any form without the express written permission from the Chief Author/Investigator and / or Sponsor.

#### Authors

The PHOSP-COVID Study Consortium.

*Some wording adapted from ISARIC/WHO Clinical Characterisation Protocol for Severe Emerging Infections Version 3.1*

#### Signature Page

**Chief Investigator Name:** ____________________________________

**Chief Investigator signature:** __________________________________

**Date:** __________________________________________________

**Sponsor Representative Name:** _________________________________

**Sponsor Representative signature:** ______________________________

**Date:** __________________________________________________

**Principal Investigator Name:** __________________________________

**Principal Investigator signature:** _______________________________

**Date:** __________________________________________________

(In cases of Multi-centre studies, this must be replicated for each site)

9. STATISTICS ... 39
9.1 Description of Statistical Methods ... 39
9.2 The Number of Participants ... 39
9.3 The Level of Statistical Significance ... 40
9.4 Procedure for Accounting for Missing, Unused, and Spurious Data. ... 40
10. QUALITY CONTROL AND QUALITY ASSURANCE PROCEDURES ... 41
11. CODES OF PRACTICE AND REGULATIONS ... 42
11.1 Ethics ... 42
11.2 Standard Operating Procedures ... 43
11.3 Declaration of Helsinki ... 43
11.4 ICH Guidelines for Good Clinical Practice ... 43
11.5 Approvals ... 43
11.6 Participant Confidentiality ... 44
11.7 Other Ethical Considerations ... 44
12. DATA HANDLING AND RECORD KEEPING ... 45
13. Data and sample Access and Sharing ... 46
14. REFERENCES ... 47

#### 1. AMENDMENT HISTORY

| Amendment No. | Protocol Version No. | Date issued | Author(s) of changes | Details of Changes made |
|---|---|---|---|---|
| 1 | 2 | 10-Aug-20 | PHOSP Team | 1. Minor clarifications and corrections.<br>2. Addition of 2 new participating NHS Organisations.<br>3. Introduction of a new questionnaire<br>4. Change to radiation research exposure<br>5. Update to the OID<br>6. Clarification of PIs and sites<br>7 Replacement of ISARIC Independent Data And Materials Access Committee with External Scientific Advisory Board for study oversight<br>8. The section on muscle biopsy sampling has been changed to allow for centres to use either a forceps or microbiopsy technique. |
| 2 | 3 | 01-Sep-2020 | PHOSP Team | 1. Addition of transfer of radiology reports with imaging data.<br>2. Addition of consultee declaration for Tier 1<br>3. Update to consent procedures:<br>a. Participants can consent on the same day as receiving study information |

| | | | | |
|---|---|---|---|---|
| | | | | b. Separate consent for postal, face to face and telephone consent<br>c. Telephone consent does not require participant counter signature<br>4. Addition of a demographic questionnaire<br>5. Addition of Participant Identification Centres<br>6. Minor Clarifications. |
| 3 | 4 | 02-Nov-2020 | PHOSP Team | 1. Section 5.3: Additional upper airway sampling techniques added to research sampling.<br>2. Clarification on adverse event / serious adverse event reporting |
| 4 | 5 | 24-Nov-2020 | PHOSP Team | 1. Addition of e-consent as a method of consenting to the study. Update to figure 3.<br>2. Addition of the collection of patient reported outcome measures via True Colours App for Tier 1 patients<br>3. Addition of 'basic tier 2' level for patients in Wave 2 recruited from 01.12.20. There is a minimum list of elements included in this 'basic' Tier 2.<br>4. For patients (those admitted Feb – April |

| | | | | |
|---|---|---|---|---|
| | | | | 2020) who have been unable to participate in Tier 2 for the first research visit at '3 months' post discharge, if they are willing, they will be invited to a Tier 2 12 month visit.<br>5. Minor clarifications and updates to figures<br>6. Clarification that recruitment to Tier 1 and Tier 2 may be split over more than 1 hospital<br>7. No cost extension |

#### 2. ABBREVIATIONS

| | |
|---|---|
| AE | Adverse event |
| AR | Adverse reaction |
| ARC | Applied Research Collaboration |
| BAME | Black, Asian and Minority Ethnic |
| BRC | Biomedical Research Centre |
| CI | Chief Investigator |
| COVID-19 | Beta-Coronavirus Disease |
| CPAP | Continuous Positive Airway Pressure |
| CRA | Clinical Research Associate (Monitor) |
| CRC | Clinical Research Collaboration |
| CRF | Case Report Form |
| CRO | Contract Research Organisation |
| CT | Clinical Trials |
| EC | Ethics Committee (see REC) |
| ECMO | Extra-Corporeal Membrane Oxygenation |
| GCP | Good Clinical Practice |
| GP | General Practitioner |
| GTAC | Gene Therapy Advisory Committee |
| IAPT | Improving Access to Psychological Therapies |
| ICF | Informed Consent Form |
| NHS | National Health Service |
| NRES | National Research Ethics Service |
| PI | Principal Investigator |
| PIL/S | Participant/ Patient Information Leaflet/Sheet |
| R&D | NHS Trust R&D Department |
| REC | Research Ethics Committee |
| SAE | Serious Adverse Event |
| SAR | Serious Adverse Reaction |
| SOP | Standard Operating Procedure |
| SUSAR | Suspected Unexpected Serious Adverse Reactions |
| TMF | Trial Master File |
| TRC | Translational Research Collaboration |

#### 3. BACKGROUND AND VISION

The peak of the first wave of COVID-19 is over. There is an urgent public health need to understand the underlying mechanisms driving the sequelae of the COVID-19 infection especially in the post-hospitalisation survivors to improve care for future waves of patients.

Our vision is to urgently create a national platform that integrates research and clinical service to understand and improve long-term outcomes for survivors of a hospitalisation with beta-coronavirus (SARScoronavirus 2, or SARS-CoV-2) disease (COVID-19). All patients discharged from hospital with a diagnosis of COVID-19 from recruiting sites will be invited to join a large national cohort at a variety of time points, maximising recruitment (at discharge from hospital, follow-up telemedicine appointments relating to their COVID-19 hospitalisation, or upon subsequent presentation at follow-up hospitalisations and out-patient clinics).

The long-term sequelae of COVID-19 after discharge from hospital are unknown and the trajectories are likely to be heterogeneous. Whilst some survivors will have a full recovery, others will have or will develop organ-based chronic disease, deconditioning, mental health issues and often a combination of these multi-morbidities. The challenges are thus to understand the magnitude of the burden of this post-COVID-19 chronic disease as a whole and in specific at-risk groups; the underlying mechanisms and biomarkers to predict future risk, the most efficacious and cost-effective pharmacological and non-pharmacological interventions and how to construct long-term integrated clinical and research pathways required to tackle future waves of COVID-19 and other pandemics.

Patients hospitalised with COVID-19 most commonly suffer pneumonitis, which can lead to respiratory failure requiring supplemental oxygen, respiratory support such as continuous positive airway pressure (CPAP), invasive ventilation or extra-corporeal membrane oxygenation (ECMO). The acute sequelae include a systemic inflammatory response, myocarditis, acute kidney injury, liver impairment, and haematological disturbances (1;2). The short-, medium- and long-term consequences of COVID-19, and the factors that influence resolution of symptoms and disease are not yet understood. Recovery may be influenced by different factors including: 1) individual characteristics such as age, gender, ethnicity (black, asian and minority ethnic background - BAME), socioeconomic status, 2) pre-existing co-morbidities (3-6) and clusters of co-morbidities including mental health, 3) lifestyle factors such as tobacco smoking, obesity, cardiorespiratory fitness and physical activity levels, 4) severity of the acute COVID-19 illness and duration of hospitalisation, 5) in-hospital intervention such as invasive ventilation or pharmacological treatment via COVID-19 Investigational Medicinal Product (IMP) studies, 6) genetic, and 7) environmental factors.

Previous cohorts of survivors of adult respiratory distress syndrome report persistent physical function and psychological impairment five years post admission (7-10). Effects of COVID-19 infection, hospitalisation and recovery on a range of health-related traits are therefore expected, including long-term symptoms such as breathlessness, multi-morbidity, physical inactivity and deconditioning, psychological and social well-being. Evidence based rehabilitation programmes are available for a wide range of long term conditions (11-13) in stable disease and immediately post-hospitalisation (14) with an evolving literature around post-intensive care for adult respiratory distress syndrome (ARDS) (15). The established cardiac rehabilitation model most closely mirrors the hospitalisation for COVID-19., i.e phase 1- in-hospital, phase 2- support post discharge, phase 3- exercise and education programme, phase IV – maintenance. However, further adaptation and new partnerships are needed to incorporate the needs of heterogeneous group of survivors particularly around methods of delivery incorporating digital technology, behaviour change strategies, mental health, and reducing future cardiometabolic risk.

Through the new clinical COVID-19 recovery care pathway, we will implement a UK research clinical cohort with a work-stream approach to the research programme. The aim is to recruit from sites that were involved in the acute COVID19 IMP trials, which included core centres from the Respiratory-Translational Research Collaborations (TRCs) network of ten centres of excellence, as they demonstrate research capacity and agreement in principle is in place for at least ten further sites to recruit with enhanced data capture (Mental Health TRCs, NHS trusts with Biomedical Research Centres, Birmingham, Newcastle, Scotland, Wales and Northern Ireland). The European Respiratory Society has recently established a Clinical Research Collaboration (CRC) for COVID-19 and the aim is for this and clinical service specification across other countries to be aligned where possible through national respiratory societies. We have agreed support from integrated infrastructure across the NIHR with Biomedical Research Centres (BRCs), Translational Research Collaborations, and Applied Research Collaborations (ARCs).

The PHOSP-COVID study is aligned to the clinical follow-up provided but does not alter clinical care. Figure 1 provides an overview of the clinical care and research pathway.

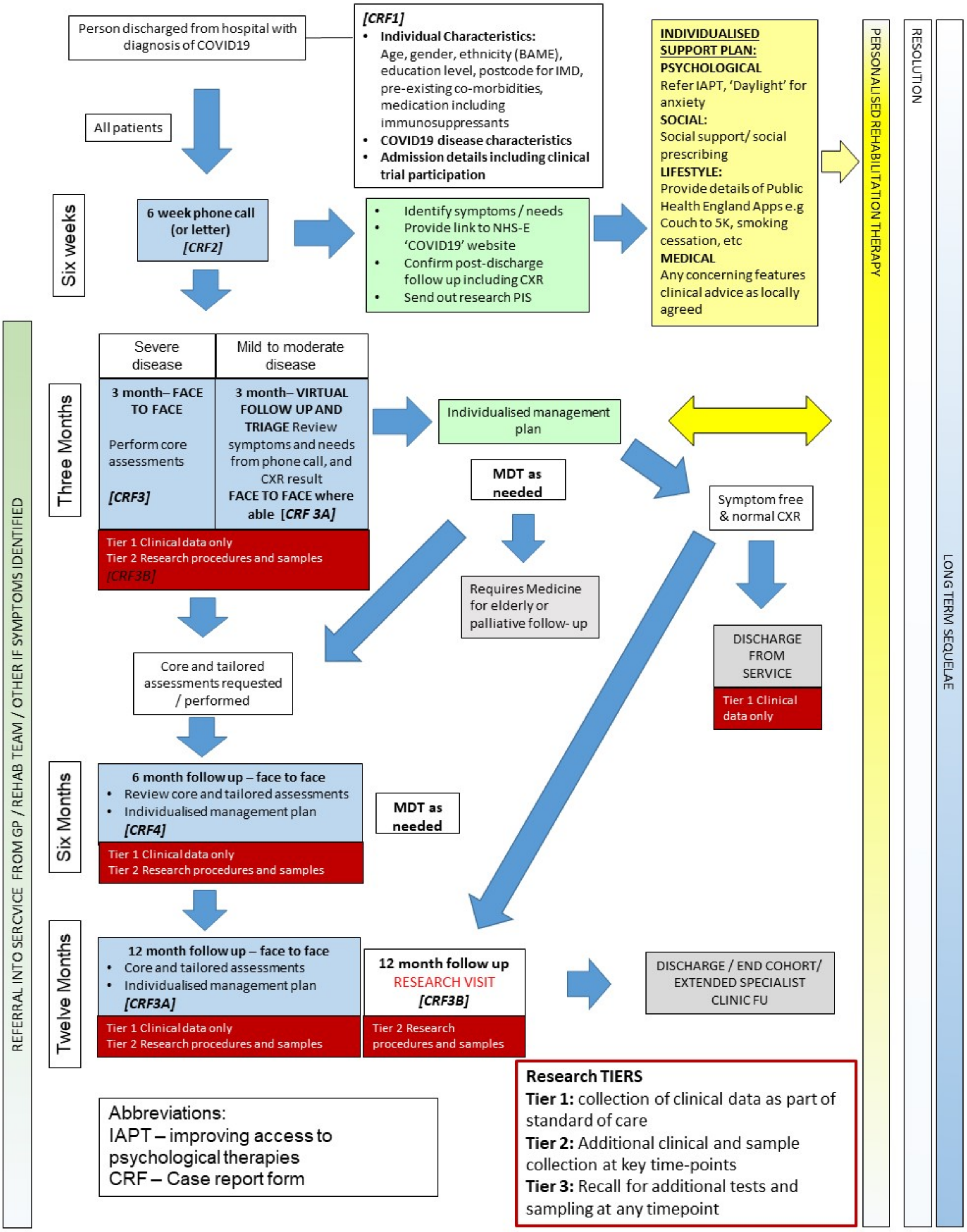

**Figure 1: Clinical pathway with data collection for research purposes indicated in red.**

#### 4. STUDY AIMS

We have established a national consortium to develop a platform to integrate research and clinical service to understand and improve long-term outcomes for survivors of a hospitalisation with COVID-19. In concert with addressing the specific aims outlined below, we shall form a stakeholder group (see Figure 7 for detail) from the outset through the **"Coordination and Data Management"** work stream (WS1) including a UK post COVID-19 Patient and Public Involvement (PPI) group. The stakeholder group will enable and support further hypothesis-led research that will utilise samples and data from the clinical study as well as enable the development of additional data modules where necessary to address our understanding of and to improve the long-term outcomes for survivors of a hospitalisation with COVID-19. The undertaking of the "**Clinical Study**" (WS2), the analyses of the "**Long-term Sequelae of COVID-19**" (WS3), impacts of interventions (WS4) and the development of **Health Services Design** through new interventions and models of care to support future precision medicine for COVID-19 (WS5) will be underpinned by enabling technologies such as multi-omics and imaging (WS6, "**Platform Science**").

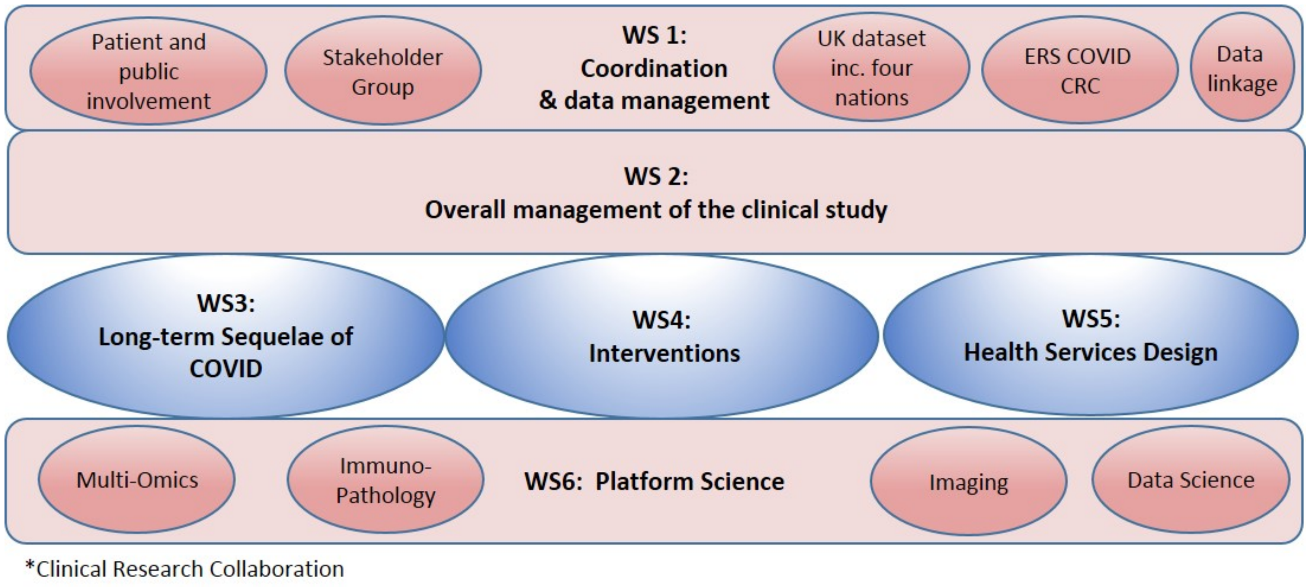

**Figure 2: Work-stream Model for Post-hospitalisation COVID-19 research programme**

**Aim 1:** To determine the short to long-term chronic health (and health economic) sequelae of COVID-19 infection in post-hospitalisation survivors; to define demographic, clinical and molecular biomarkers of the susceptibility including severity of the acute illness, development, progression and resolution of these health sequelae.

- Determine the short- (0-6 months), medium- (6-12 months) and long- (12 months and onward) term sequelae to COVID-19 including re-hospitalisation and mortality, symptoms, organ impairment, impact on physical function, psychological profile, and social consequences.
- Identify the association between short- to long- term outcomes and demographics (with a focus on Black, Asian and Minority Ethnic (BAME) background, socioeconomic status, lifestyle factors), genetics and other biomarkers, pre-existing co-morbidities and clusters of co-morbidities including mental health, and severity of acute illness stratified by requirement of supplemental oxygen, respiratory support such as continuous positive airway pressure

(CPAP), invasive respiratory support, and invasive ventilation and renal support such as central veno-venous haemofiltration or haemodialysis

- Where possible aim to link with other studies to evaluate (and compare with) a control group of individuals with COVID-19 with milder disease not hospitalised. We would aim to link with community testing programmes, potentially invite individuals to attend as comparison group (outside of this protocol) and utilise ongoing general population studies for comparison of sequelae in mild vs. severe disease (via new and existing longitudinal cohort studies and resources with COVID-19 research capability including EXCEED (13/EM/0226), Coronagenes, NIHR Bioresource and UK Biobank).

**Aim 2:** To understand the impact of acute and post-discharge, pharmacological and non-pharmacological interventions on long-term sequelae of COVID-19 in post-hospitalisation survivors; to define the determinants of response to inform future precision medicine as per Aim 1.

- To determine whether acute pharmacological interventions administered in hospital (e.g. RECOVERY and or REMAP-CAP trials) impact on the long-term sequelae of COVID-19 in post-hospitalisation survivors
- To understand retention and responder analysis of rehabilitation interventions
- To enable the study of novel interventions within the rehabilitation pathway to include self-management and behaviour change strategies, digital interventions and other modes of delivery, mental health interventions, wider multi-disciplinary team involvement to target obesity, diabetes and multimorbidity, and incorporating principles of reducing treatment burden

**Aim 3:** To build the foundation for multiple in-depth studies of emergent and worsening of premorbid disease to inform precision medicine in at risk groups by directing new clinical trials and care for current and future post-COVID-19 patients.

- Enable recall of COVID-19 survivors (based on demographics, health status or biomarkers including imaging and genetic variation) for further studies to address emerging research hypotheses related to physiological responses, health sequelae and effects on co-morbid conditions of COVID-19 (this will further enhance Aims 1 and 2)
- To map the treatments and care received by COVID-19 patients after discharge from hospital
- Understand the patient and healthcare professional experience to inform future integrated (both horizontal and vertical) care strategies
- Embed different ways of engaging patients, particularly BAME background representation, in the clinical cohort, rehabilitation and related research underpinned by behaviour change strategies and principles of reducing treatment burden particularly for multi-morbidity
- Understand the health economics of a holistic integrated model of care
- Inform long-term new models of care with integrated clinical and research pathways for further waves of COVID-19 and future pandemics
- To develop a master protocol for disease-specific intervention trials for post-COVID-19 patients
- To inform interventions and healthcare for future waves

The in-depth projects will include focussing on lung (fibrosis, pulmonary vasculature, bronchiolitis and bronchiectasis), systemic vascular, cardiometabolic, renal, and neurological disease, sarcopaenia, fatigue and rehabilitation, immunology, and mental health. Within our consortium we have existing established expert groups within each domain that have developed research plans. 3-4 exemplar projects will be prioritised based on early insights from the study).

For emergent disease, we shall determine the incidence of new disease based on extensive clinical investigation. The disease-specific research groups shall forensically phenotype these patients with access to the clinical meta-data, multiomic and imaging data (as per Aim 1). Biomarkers of specific interest can be analysed from the bioresource samples and additional quantitative analyses of the imaging. Whether treatments for the acute illness modify the disease incidence, severity or trajectory will be determined (Aim 2) and implemented in future waves. Insights from Aim 1 and 2 will inform recall studies for further subgroup characterisation and biosampling to support further mechanistic studies for the exemplar projects (Aim 3).

A master protocol will be developed for disease-specific precision medicine intervention trials for those post-COVID-19 that will require further funding (e.g. antifibrotic agents for interstitial lung disease, macrolides for bronchiectasis, rehabilitation interventions, anti-thrombotic agents for thromboembolic disease, mental health interventions for PTSD and severe psychiatric disease) and will inform interventions for future waves (Aim 3).

For worsening of premorbid conditions, we shall focus our attention on those where there is a step-change in their underlying disease. The approach will then be as for emergent disease. High-risk groups might include those with severe cardiorespiratory disease and chronic renal disease receiving dialysis, amongst others.

This study will provide control groups by comparing those that do versus do not develop a chronic condition or worsening of a premorbid condition, critical care studies for non-COVID-19 cases and will link to UK Biobank where relevant for population controls and especially for those with pre-COVID data including physiology and imaging, disease-specific cohort studies and epidemiology cohorts consented for recall.

The immunological response to COVID-19, immunoprotection and its relationship to the above conditions will be studied partly here and with a proposed British Society of Immunology Consortium.

The PHOSP-COVID programme will recruit individuals who were discharged from hospital following suspected COVID-19 and study the short (0-6 months), medium (6-12 months) and long-term (12 months and onward) effects of the disease. We propose to analyse routine clinical data (Tier 1) with enhanced clinical data at some sites (Tier 2), research-specific biomarkers (Tier 2) measured at routine follow-up clinic visits including rehabilitation, through linkage to retrospective and prospective health and social care records (including primary care, hospital episodes statistics, pathology records, prescribing records and specialist tertiary clinical databases), and through re-call of participants with particular characteristics for more detailed studies (Tier 3). The research will be embedded alongside the standard clinical care pathway outlined in Figure 1 and Table 1 as illustrated in Figure 3 below.

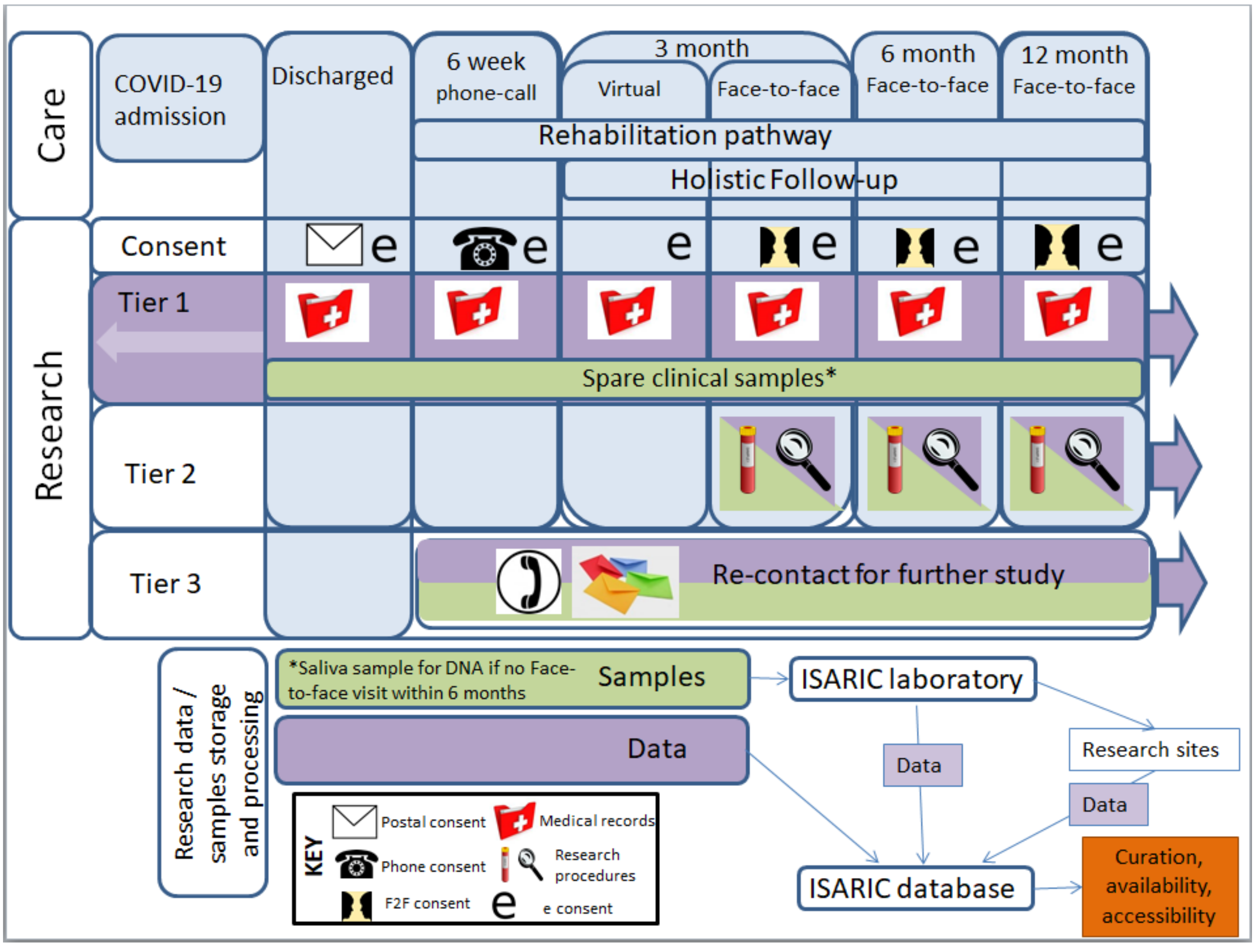

**Figure 3: Alignment of research data collection with clinical care pathway**

#### 5. STUDY DESIGN

##### *5. 1 Overview*

All patients discharged from hospital with clinician suspected COVID-19 from UK recruiting hospital sites will be invited to join the study.

Research data will be collected according to a Research Tier system to enable collection of data and samples to be tailored according to centre capacity and specific research questions to be addressed (**Figure 1, Figure 3** and **Figure 4**):

**Tier 1:** Clinical data only, healthcare records (to include primary care, secondary care (NHS digital / NHS-X), social care and prescriptions), saliva kit by post for DNA, consent for use of 'left-over' clinical samples, collection of patient reported outcome measures via True Colours App (pg 29).

**Tier 2:** Tier 1 plus extended data obtained from additional research procedures and samples

**Tier 3:** Research-specific recall for additional procedures and sampling including semi-structured interviews. Recruitment to external research studies requiring additional consent.

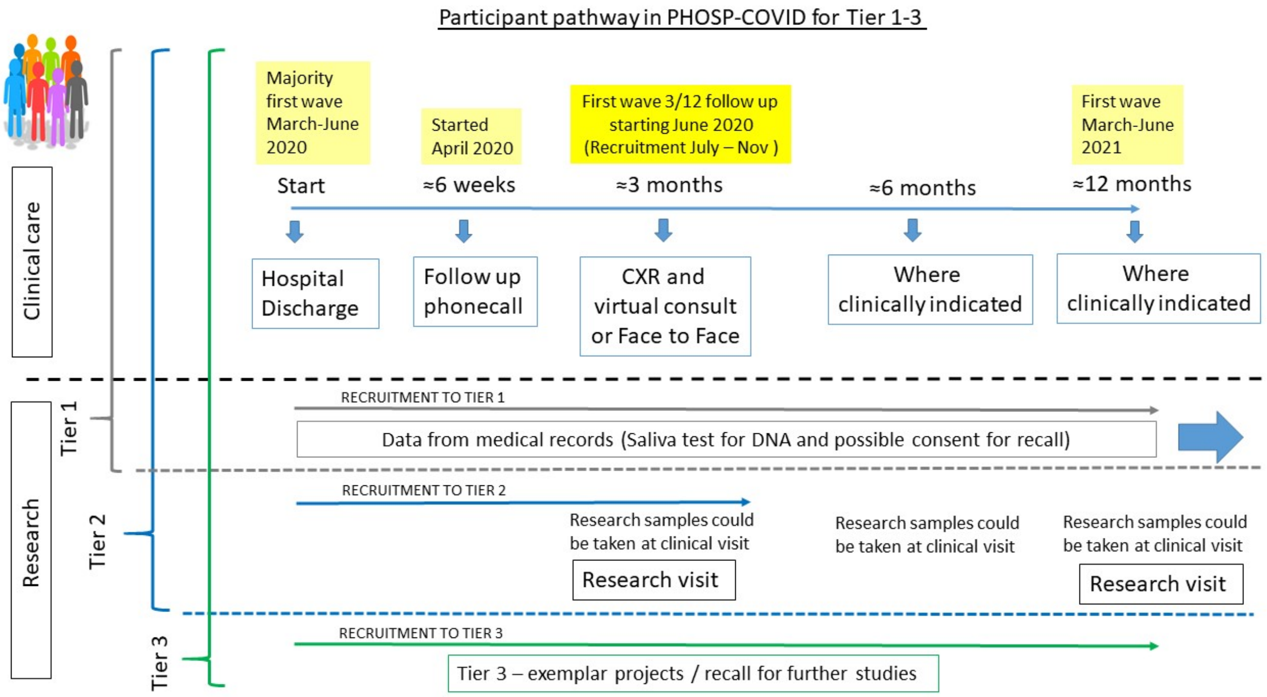

**Figure 4: Participant pathway for Tiers 1 to 3.**

All patients discharged from participating centres will be invited to participate in Tier 1 and patients can be recruited to Tier 1 at any time point over a year post-discharge. Recruitment to Tier 2 will be offered to all patients from discharge to their three month clinical review. There will not be explicit stratification for recruitment to Tier 2, but demographics of recruited patients will be reviewed at regular intervals (for example, at recruitment of 1000$^{th}$ participant) to ensure that important groups are well represented (BAME background – in relation to proportion of admissions, specific comorbidities, severity of acute illness etc).

The PHOSP-COVID study is aligned to clinical care but the study will neither alter nor depend on the clinical pathway. Consent for linkage to all results from assessments performed as part of clinical care will be requested for participants in Tier 1 and Tier 2, specifically:

- Any imaging (chest x-ray, CT imaging including CTPA, MR imaging, multi-organ MR) **including radiology reports** will be pseudonymised and transferred to Royal Surrey County Hospital and then to NCIMI and hosted securely.
- More detailed pulmonary function tests
- Echocardiogram
- Cardiopulmonary exercise testing
- Blood tests not already specified
- Urinary tests not already specified
- Further questionnaires or interview data as part of clinical consult
- Pre-post intervention data as part of clinical care

Clinical interventions

We will record any clinical interventions patients receive for example rehabilitation, mental health interventions such as Improving Access to Psychological Therapies (IAPT) and nutritional interventions. We will record any available results before and after the interventions.

##### *5.2 Research Tier 1:*

We will collect baseline information from all participants relating to their COVID-19 admission, demographics (via a demographics questionnaire which may be completed face-to-face, over the phone or self-completed following consent) including ethnicity and socioeconomic status, prior functional and physiological status. We will request participant consent or consultee declaration to link to all retrospective and prospective electronic healthcare records including primary care data, social care data, Hospital Episodes Statistics and Office of National Statistics (and others available via NHS Digital), and specialist clinic data obtained from out-patient and in-patient visits. Patient NHS number will be used to undertake linkage. The data will include but not be limited to investigations including imaging, physiological measures, questionnaires and blood test results obtained whether related to the participants' COVID-19 admission or not. We will perform data extraction from medical case files at regular intervals throughout the study. To assess the environmental contribution to COVID-19-related health factors, we will link to available environmental variables (including air pollution data) using the participant address.The True Colours App discussed Pg 29 will be available for patients in Tier 1 to complete the patient reported outcome measures.

We will request participant consent for collection, storage and testing of a DNA sample, obtained either from spare blood that remains after clinical testing or from a saliva kit that we will despatch when feasible to do so. Those participants recruited via Consultee Declaration will not be eligible for any study procedures including DNA/RNA sample collection. The appropriate statements on the consent form should be marked as 'no' as directed in the instructions on the informed consent form.

##### *5.3 Research Tier 2:*

Table 1 (Pg 18) shows the complete dataset for Tier 2. Participants may be asked to provide additional samples, undertake additional tests or provide additional information (for example via questionnaire) when they attend routine/clinical follow-up appointments to assess the resolution of their COVID-19 illness (see **Figure 1 and Table 1**) or as a research visit or as a combination of the two. ). A research visit is proposed as part of Tier 2 at 3 months (2-7 months +/-2 weeks) and 1 year post discharge (+/- 2 months). These procedures will be in addition to those undertaken as part of their standard clinical care (data from their standard clinical care will be extracted as described for Tier 1). Exactly which assessments are conducted as part of clinical care will vary according to the individual need and the clinical service at a particular centre. We describe an example of the scope and types of data that would be collected but is not exclusive. Participants will be contacted by phone or post to arrange visits, according to local site practice.

We have a no cost extension to our grant from UKRI to extend recruitment to wave 2. A 'basic' Tier 2 level is therefore being added for patients in Wave 2 recruited from 01.12.20 for at least six months. The minimum list of elements included in this 'basic' Tier 2 are highlighted in yellow in Table 1 (pg 18).

Some patients (those admitted Feb – April 2020) will have been unable to participate in Tier 2 for the first research visit at '3 months' post discharge where sites were not open in time,where participants are willing they will be invited to a Tier 2 12 month visit.

Sites will include Translational Research Centres (TRCs) and Biomedical Research Centres (BRCs) as well as Scottish, Welsh and Northern Ireland networks. Samples and tests required for clinical management will at all times have priority over samples taken for research tests. Wherever practical, taking research samples will be timed to coincide with clinical sampling. The research team will be responsible for sharing the sampling protocol with health care workers supporting patient management in order to minimise disruption to routine care and avoid unnecessary procedures.

Research samples will be minimally processed at site and transported to the ISARIC Outbreak laboratories in Liverpool for coordination and re-distribution to research sites for assays and analysis. Samples will be transported in line with Standard Operating Procedures for Category B shipping requirements where appropriate.

Table 1. Modular holistic follow up to enable identification of long term sequelae and personalised management. Minimal procedures for ‘basic’ Tier 2 for Wave 2 participants highlighted in yellow.

| Module | Tier 2 common dataset including detailed clinical phenotyping |
|---|---|
| **Symptoms** | *Report and track new or change in existing symptoms since COVID with numeric ratings scale for severity**<br>*MRC Dyspnoea Grade*,*<br>Q: Patient symptom questionnaire, Dyspnoea12 Questionnaire, FACIT – fatigue Questionnaire, Brief Pain Inventory |
| **Quality of life** | Q: Euroqol (EQ5D-5L) enabling calculation of quality-adjusted life years |
| **Respiratory** | *Resting pulse oximetry ($SpO_2$), Spirometry, Transfer Factor (DLco)*, max inspiratory pressure (MIP), max expiratory pressure (MEP) |
| **Cardiac** | *Resting BP & heart rate,*<br>*Electrocardiogram,*<br>*Blood tests: BNP/NT- BNP, Troponin* |
| **Renal / bone / liver** | *Blood tests: Urea & Electrolytes including eGFR, Bone chemistry, Vitamin D, Liver Function Tests**<br>*Urinalysis, albumin:creatinine ratio, protein:creatinine ratio* |
| **Haematological** | *Blood tests: Full blood count, D Dimer*, INR, Fibrinogen, Ferritin |
| **Systemic inflammation** | Blood test: CRP |
| **Physical function and activity** | Q: *GP Physical Activity questionnaire (GPPAQ)*,*<br>*Incremental Shuttle walking test to assess exercise capacity*,*<br>*Handgrip and quadriceps strength*,*<br>Objective Daily Physical Activity monitoring for 2 weeks using an activity monitor |
| **Frailty and activities of daily living (ADL)** | *Q: Clinical Frailty Scale*,*<br>*Short Physical Performance Battery (SPPB)*<br>Q: Nottingham ADL<br>Fried’s Frailty assessment – assessed using the information captured in Tier 2. |
| **Body composition** | *Body Mass index*,*<br>*Malnutrition Screening Tool (MUST)**<br>Q: Sarc-F (screening tool for those at risk of sarcopenia),<br>Body composition |
| **Mental Health** | *Q: Generalised Anxiety Disorder Assessment (GAD-7)*,*<br>*Q: Patient Health Questionnaire (PHQ)-9*,*<br>Q: Post-Traumatic Stress Disorder Checklist for *DSM-5* (PCL-5) |
| **Cognition** | Q: Montreal Cognitive Assessment (MOCA) |
| **Cardiometabolic risk** | Qrisk3 score, (fasting) lipids, fasting glucose & insulin,<br>HbA1C,<br>waist circumference<br>Homeostasis model assessment –insulin resistance (HOMA-IR) score and presence of metabolic syndrome to be calculated. |
| **Research samples** | Saliva<br>Blood<br>Urine<br>Sputum<br>Oral rinse<br>Upper Airway |

**may be performed as clinical care but when not completed will be performed as part of research to enable a complete dataset. Q= participant completed questionnaire*

The participant information sheet will ask for consent for these additional procedures to be requested. The participant will be provided with additional information and the opportunity to ask questions before proceeding with any additional procedures. They will be free to refuse to undertake any additional procedures and this will not affect their clinical care or continued involvement in the study.

Tier 2 research testing will be undertaken depending on the research sites capacity and capability. Also, participants may be required to attend more than one NHS Hospital in order to participate in both Tier 1 and Tier 2 of the study. This is due to local capacity and capability arrangements at different hospitals. In this scenario, the participant will be asked to sign a consent form at the Tier 1 hospital and then re-sign a part of the same consent form at the Tier 2 hospital. Therefore, a copy of the signed consent form will be securely transferred between the hospitals.

<u>Example Research visit (2.0 – 3.0 hrs)</u>

| | |
|---|---|
| *30 mins* | Consent |
| 15 mins | Blood tests (including fasting where possible), height, weight and bioelectrical impedance |
| Drink and snack | |
| 30-40 mins | Tests if not performed clinically: lung function, electrocardiogram |
| 40 mins | Walk tests (x2 with at least 20 mins rest between walks) |
| | Complete questionnaires during 20 min rest |
| 20 mins | Handgrip and quadriceps measurement |
| | Short Physical Performance Battery (SPPB) |
| 20-30 mins | Complete the remaining questionnaires |
| 5 min | Provide the activity monitor explanation |

Where possible, visits will be restricted to a single appointment as previous patient representatives with long term conditions have stated a preference for longer visits and to limit the amount of travel. However, we will personalise the approach to the research visit strategy and content based on an individual's preference and appreciate this might vary for patients suffering from fatigue or cognitive impairment for example.

Example Research visit for Tier 2 'basic' (1 – 1.5 hrs)

| | |
|---|---|
| *10 min* | Review Consent |
| 15 mins | Blood sample (including fasting where possible), height, weight, waist circumference |
| Drink and snack | |
| 30 mins | Tests if not performed clinically: spirometry, electrocardiogram |
| 10 mins | Handgrip strength |
| | Short Physical Performance Battery (SPPB) |
| 5 min | Discuss the activity monitor with explanation |
| 10min | Review True Colours App and collection of PROMS |

| | |
|---|---|
| Same day or different day | CXR if not performed clinically (rare) |

##### *Research Samples*

The specific assays to be undertaken using biological samples will be defined by the particular research question being addressed and informed by current and emerging literature.
Additional Tier 3 samples, which will be undertaken in subsets of participants, are also described here.

*Blood:* Blood samples will be up to 100ml within a 4 week period and the first blood draw would be no sooner than the 3 month visit after discharge (**Figure 1**). For DNA (genetic variation and telomere measurement), blood will be stored in K2-EDTA tubes until extraction and analysis. For RNA (gene expression), blood will be stored in tubes that contain preservatives for RNA (e.g. PAXgene or Tempus RNA tubes) until extraction and analysis. For proteomics assays, blood samples will be centrifuged to extract plasma and stored prior to analysis. For other individual biomarker measurements, the blood sample will be processed and stored prior to analysis as whole blood, serum or plasma. Blood samples will also be used to develop cell lines for laboratory experiments and to study individual cells. Whole blood cell counts will also be obtained from the blood samples.

Where not performed as part of clinical care blood samples will be analysed at 3 and 12 months for full blood count (FB), glomerular filtration rate (eGFR), liver function tests (LFTs), bone profile, lipid profile, brain natriuretic peptide, troponin, glycosylated haemoglobin, vitamin D levels, D dimer and c-reactive protein levels. These will all be used to assess the multi-organ impact of COVID19 infection.

Research bloods will be transferred from the Investigative Site to Liverpool Good Clinical Practice Laboratory (GCPLab).

*Saliva:* This will involve participants providing a saliva sample into a storage tube. Participants will be asked to refrain from smoking, eating, chewing gum or drinking for up to 30 minutes before providing their sample. Saliva samples will be used to obtain DNA from participants who do not attend further clinics.

*Urine:* Urine will be collected into a suitable container, processed and used to measure proteins and metabolites. Where not performed as part of clinical care, urine samples will be analysed at 3 and 12 months for urinalysis, albumin creatinine ratio and protein creatinine ratio.

*Sputum:* Sputum will be collected by spontaneous sputum sample collection. Participants will be provided a sterile sample container in which to provide a spontaneous sample.

(16)

*Upper airway sampling:* Upper airway samples will be collected using one or more of the following methods. This should take a few minutes and is successful in most subjects but may cause minor discomfort or bleeding.

Oral rinse: An oral rinse using sterile saline solution will be used to collect upper airway samples for cell and microbiome analysis.

Nasosorption: A small piece of synthetic absorptive matrix (SAM) that feels like paper will be inserted into the nostril to collect nasal fluid for approximately one minute.

Nasal Scrape: A small nasal brush and/or “rhinoprobe” will be inserted and gently rubbed inside the nostril in order to collect nasal cells.

Nasopharyngeal swab: A nasopharyngeal swab will be gently inserted through the nose, rotated and then removed. The same process can be performed in the throat.

##### *Imaging*

We have formed an Imaging Alliance for thoracic CT, cardiac MRI and multi-modality whole body MRI imaging which builds on expertise in artificial intelligence and radiomics. Any CT thorax performed as clinical care will be linked, alongside other clinically-indicated X-Ray and MRI imaging where available, via NHSX and the National COVID-19 Chest Imaging Database (NCCID).

For clinically indicated thoracic CT scans the PHOSP-COVID imaging alliance have developed a suggested imaging protocol for sites to use.

At least 500 patients recruited to PHOSP-COVID will be selected for advanced research imaging in at least 6 sites, where specialist imaging facilities are available (for example Glasgow, Leeds, Leicester, Bristol, Oxford and other sites will be added).

Royal Surrey County Hospital are responsible for data transfer and anonymisation from local sites using the IEP network and data linkage.

##### *Physical Tests at 3 and 12 months.*

*Electrocardiogram* if not performed as part of clinical care.

*Physical activity and sleep monitor:* Participants will be fitted with a blinded wrist worn accelerometer. The devices are well tolerated, and weigh less than 100g. Accelerometers will be worn for two weeks at the 3 month and 12 month visits. Monitors will be initialised to record triaxial acceleration (g) at 50 Hz. Participants will post the monitors back to centres and the researchers will download the data.

*Short physical performance battery (SPPB)*: This includes the 4 metre gait speed (the time it takes to walk 4 metres), a balance test, and sit to stand time. The outcome is associated with frailty (17).

*Muscle strength:* Handgrip strength will be recorded using a handheld dynamometer. Knee extensor isometric strength will be performed on an adapted chair with a strain gauge (18). From a seated position patients are asked to flex their knees at 90° over the end of the chair. A strap is placed around the ankle and connected to a measuring device. Patients are then asked to push as hard as they can against the strap. For handgrip and knee-extensor strength, measurements are performed until 3 measurements are within 5% of each other. Typically 3-6 manoeuvres are performed.

*Incremental Shuttle Walk Test (ISWT)*: The incremental shuttle walk test is a validated test of maximal exercise capacity (19). Participants will be given standardised instructions on how to complete the test via audiorecording. The course is 10m, set with two cones 9m apart giving a turning distance of 1m at either end. Participants are asked to walk along the course at a speed indicated by an audio signal (bleeps). The aim is to have walked around the cone by the time the audio signal is given. The pace is very slow to begin with (0.5m/sec) and the speed increases every minute. Participants are advised to continue until they are too breathless or too tired to continue or can no longer keep up with the required speed. The distance walked is then calculated. A shuttle is completed if the patient reached within 0.5 m of the cone. The operator gives no encouragement throughout the test. Auxillary measurements of heart rate and oxygen saturation level are monitored continuously throughout the test via pulse oximetry and non-invasive blood pressure recorded before and upon completion of the test. The Borg Scale for peak breathlessness (BS) and perceived exertion (PE) are measured at rest and at the end of the test.

In centres with the necessary expertise a full cardiopulmonary exercise test may be performed and in line with local Trust policy around PPE measures.

*Cardiopulmonary exercise test on a cycle ergometer*

A full cardiopulmonary exercise test allows an integrated assessment of a person’s cardiorespiratory fitness and the cardiovascular, respiratory, and muscular systems. Patients are monitored during rest, warm up (unloaded pedalling), followed by an incremental phase to exhaustion. During the incremental phase the work load increases by a tailored amount every minute. The test will be supervised by an appropriately qualified operator. Patients will be monitored throughout the test and for at least ten minutes during recovery.

- Heart - ten small monitoring leads are placed on the back and chest
- Breathing - patients will wear a tight-fitting but light mask that has sensors that measure air movement to and from their lungs.
- Oxygen in the blood - a small sensor is worn on the finger
- Blood pressure - a blood pressure cuff is wrapped around a patients arm and is inflated briefly before, during and after exercise.

*Lung function*: measures of lung function may be categorised as aerosol generating procedures (AGP) and may require full personal protective equipment (PPE). Please follow local Trust policy on infection control when performing lung function measurements.

The following lung function would be performed when safe to do so:

- Post-bronchodilator Spirometry and transfer factor according to European Respiratory Society guidelines.
- Inspiratory and Expiratory pressures (PiMax/PeMax)

*Assessment of cardio-metabolic risk.* HbA1C, and where possible a fasting insulin, glucose, and full lipid profile will be added to blood sampling and a waist circumference to calculate metabolic syndrome. Qrisk3 score will be calculated by the researcher. https://qrisk.org/three/

*Body composition*

*Bioelectrical impedance* will be measured to assess fat free mass in all centres but the equipment may vary in difference centres. The main measurements however can be collectively interpreted. Patients will be given the precise instructions by the operator.

*Dual energy x-ray absorptiometry (DEXA) scanning.*

In some centres DEXA will be performed for a more accurate assessment of body composition and to provide appendicular measurements. It is a similar procedure to having a chest x-ray, but requires the patient to lie down. The proportion of muscle, fat and bone density in the body is measured. The radiation dose from each DEXA scan is equivalent to radiation from less than 1/10$^{th}$ of a chest x-ray. Chest x-rays expose the patient to a minimum amount of radiation.

***Questionnaires***

Some questionnaires will be either recommended medically or as part of the rehabilitation assessment in order to individually tailor management. However, we also want to comprehensively capture data on symptoms and health-related quality of life data so seek to add these questionnaires where they are not part of routine clinical care.

All questionnaires will be collected at 3 and 12 months after discharge, and every three months if participant is using True Colours (see description Pg 27):

The ‘patient symptom questionnaire’ designed for use in clinical care as part of CRF2a will also be collected at the time points above.

The ‘demographics questionnaire’ will be provided to participants following informed consent. This additional questionnaire is to collect demographic details (self-reported ethnicity, household income, gender, education level, marital status, disabilities, and exposure details). This may be asked verbally face to face or over the telephone or provided as a self-completed paper questionnaire during visits or posted to participants for completion and return alongside postal consent forms.

The following questionnaires may be completed as part of clinical care, but we seek consent to complete where missing:

- Medical Research Council Dyspnoea Grade Scale which assesses the impact of breathlessness on exertion (21)
- General Practitioner Physical Activity Questionnaire (GPPAQ) (22) which is a brief physical activity questionnaire
- Symptoms of anxiety and depression will be assessed by the Generalised Anxiety Disorder 7 item scale (GAD7) and the Patient Health Questionnaire-9 (PHQ-9) (23).
- Rockwood Clinical Frailty Scale (24).
- We will use the Post Traumatic Stress Disorder (PTSD) checklist for Diagnostic and Statistical Manual of Mental Disorders (DSM-5) (PCL-5) questionnaire (https://www.ptsd.va.gov/professional/assessment/adult-sr/ptsd-checklist.asp to assess for PTSD.
- We will use the ‘SARC-F’ questionnaire to screen for sarcopenia (https://www.cgakit.com/sarc-f-questionnaire).

These additional questionnaires will be requested to be completed:

- Euroqol (EQ-5D-5L) (https://euroqol.org) as a measure of generic quality of life and to assess quality adjusted life years.
- Activities of daily living will be assessed using the Nottingham Extended Activities of Daily Living (ADL) scale (25).
- Brief Pain Inventory questionnaire
- To evaluate specific symptoms further, we will include the Fatigue by the FACIT questionnaire (26), breathlessness by the Dyspnoea-12 questionnaire (27), and cough with the Leicester Cough Questionnaire (28).
- We will assess any cognitive impairment using the Montreal Cognitive Assessment (MOCA) https://www.mocatest.org/

There will be other specific questionnaires which maybe requested at certain centres in sub-samples of patients for example the Pittsburgh Sleep Quality Index (PSQI) and Morningness-Eveningness Questionnaire (MEQ) to understand the relationship between shift work and covid19 infection and complications.

###### *Data for health economic analyses*

We will collect data on the impact of ongoing chronic morbidity amongst patients utilising standard tools (EQ-5D) and extract data on healthcare resource utilisation from routine clinical data, supplemented with data from patient questionnaires. We will develop a simple economic model to assess both the quality adjusted life-year (QALY) losses and the incremental costs due to chronic sequelae. We will extrapolate costs and outcomes over a lifetime time horizon and take a disaggregated societal perspective, examining net costs to the health service, households, and society as a whole.

##### *5.4 Research Tier 3:*

The research may identify findings that require further samples, measurements or information from patients for additional analysis. We will request consent to re-contact participants for further studies on the basis of their demographic information, health status, genetic variation or other test results. We are unable to state exactly which samples, measurements and/or information will be needed but any proposed additional analyses will be reviewed by the Executive Board, the Steering Committee and the External Scientific Advisory Board and subject to further ethical review if appropriate. Participants will be free to decide whether or not they wish to provide further samples, measurements or information following provision of further information and opportunities for discussion with a study team member and their doctor.

***Understanding the impact, facilitators and barriers to implementing holistic integrated care pathways in the context of COVID-19.*** *T*o inform future healthcare design, we will capture the experience of people hospitalised with COVID-19 during their recovery and the experience of a novel integrated care pathway. We will perform semi-structured interviews in approximately 100 individuals post-hospitalisation for COVID-19 over 12 months and a similar number of healthcare professionals across the multi-disciplinary team. Qualitative data from patients and healthcare professionals may help understand any prognostic factors, including any interventions received, identified in the quantitative data.

***Quadriceps ultrasound***
Quadriceps ultrasound will be performed as per Standard Operating Procedure (SOP). Measures will be taken at mid-point of the thigh and three measures (within 10%) of rectus femoris cross sectional area will be taken and the mean recorded. Three measures of quadriceps thickness will be taken and the mean recorded. Images of the quadriceps will be taken (linked anonymised) for later analysis of grayscale (muscle quality).

***Skeletal Muscle Biopsy***:
Muscle biopsy of vastus lateralis (quadriceps) or another appropriate muscle for sampling will be taken under local anaesthetic. An incision (approx. 5mm) will be made in the skin and a biopsy of

muscle taken, using either a core needle biopsy device or forceps. A steristrip and bandage are then applied. Alternatively, a single stitch may be required. Muscle biopsies using this technique are well tolerated with minimal complications. There is a small risk of haematoma (bruising) and the site of biopsy will ache for about 24 hours. Participants are able to perform all normal tasks following biopsy. Patients on anticoagulants (e.g. DOACs, warfarin, clopidogrel will be precluded from biopsy)

***Multi-organ imaging***

Where capability and funding exists patients will be invited to undergo optional multi-organ (Brain, heart, liver and kidney) imaging (either computed tomography [CT] or Magnetic Resonance [MR]) to assess the frequency and severity of non-pulmonary involvement and CT imaging and or perfusion imaging for pulmonary and vascular systems as well as coronary arteries. Patients would be provided with all the relevant information about the type of imaging, and would be provided with an additional patient information sheet and consent

***Other lung function***

Measures of lung function may be categorised as aerosol generating procedures (AGP) and may require full personal protective equipment (PPE). Please follow local Trust policy on infection control when performing lung function measurements.

The following lung function would be performed when safe to do so:

- Oscillometry: where possible oscillometry will be conducted according to international guidelines (20) to investigate presence of early airway disease where spirometry is normal.

***Breath collection***

This will involve tidal breathing into a breath collection device for a few minutes to measure volatile organic compounds (16).

***Additional questionnaires***

Consent will be sought to complete additional questionnaires for certain sub-groups of patients, or particular research questions. Patients will be able to fully participate in Tier 2 without completing any further questionnaires as we are mindful of participant burden. Example questionnaires are the Pittsburgh Sleep Quality Index (PSQI) and Morningness-Eveningness Questionnaire (MEQ) to understand the relationship between shift work and COVID-19 infection and complications. Where relevant, questionnaires validated in interstitial lung disease will be used for example the IPF Prognostic Assessment and Referral to Care, and Kings Brief ILD questionnaire. The COPD assessment test may also be completed. We are seeking patient consent for other questionnaires relevant to a sub-group of patients as part of Tier 3 research projects.

***5.5 Data collection, storage and analysis***

We will provide case-report forms (CRFs) to capture data from clinical consultations through CRN support for Tier 1. Please see the Table below to illustrate the different CRFS. For patient characteristics, we are using nationally agreed data capture from the equalities act.

| Timeline | Tier 1 | Tier 2 | | Tier 3 |
|---|---|---|---|---|
| Discharge | *CRF1.*<br>Individual characteristics:<br>*age, gender, ethnicity, socioeconomic status including education level, postcode for IMD, annual salary bracket, type of employment including details on shift work, medications, co-morbidities to allow calculation of Charlson Index and accurate call back for relevant Tier 3 studies, social history: living arrangements including carers, lifestyle factors, disability*<br>Admission details*:*<br>*Description of acute disease and other acute pathology* etc, blood test results and participation in any IMP studies<br>(data capture is aligned to ISARIC, but with additional detail) | | | Participants selected based on information from Tier 1 and Tier 2.<br>Bespoke CRFs for Tier 3 studies. |
| 6 week triage phone call | CRF2: *pre-existing conditions, changes to health and symptoms since discharge including mental health, self-care and activity assessments, nutrition, cognition, self-perceived health status, social & lifestyle.*<br>*Results of investigations performed (CXR, HRCT, CTPA, etc)* | | | |
| 3 month | CRF3A = clinical review | CRF3A if clinical review and<br>CRF3B research data | | |
| 6 month | CRF4 = clinical review | *Research samples only* | | |
| 12 month | CRF3A = clinical review | CRF3A if clinical review and CRF3B research data | | |

We will also provide CRFs to allow data collection for:

1) Clinical interventions (including a description of the intervention and results pre-post for example rehabilitation, Improving Access to Psychological Therapies (IAPT) and nutritional interventions),
2) Other clinical consultations related to COVID19 and
3) Unscheduled clinical care.

Research sample data will be stored using sample record forms and laboratory record forms.

The data will be stored as text and image files using unique codes to identify each participant (pseudonymised). The link between the unique codes and names will be stored separately and securely.  A copy of the data will be held in the ISARIC Oxford/Edinburgh database to facilitate further data curation, record linkage and analyses by approved researchers via a trusted secure environment. In the longer term, a copy of the data set will be held within a safe data haven such as the SAIL Data Bank (Swansea) of HDR UK Digital Innovation Hub BREATHE.

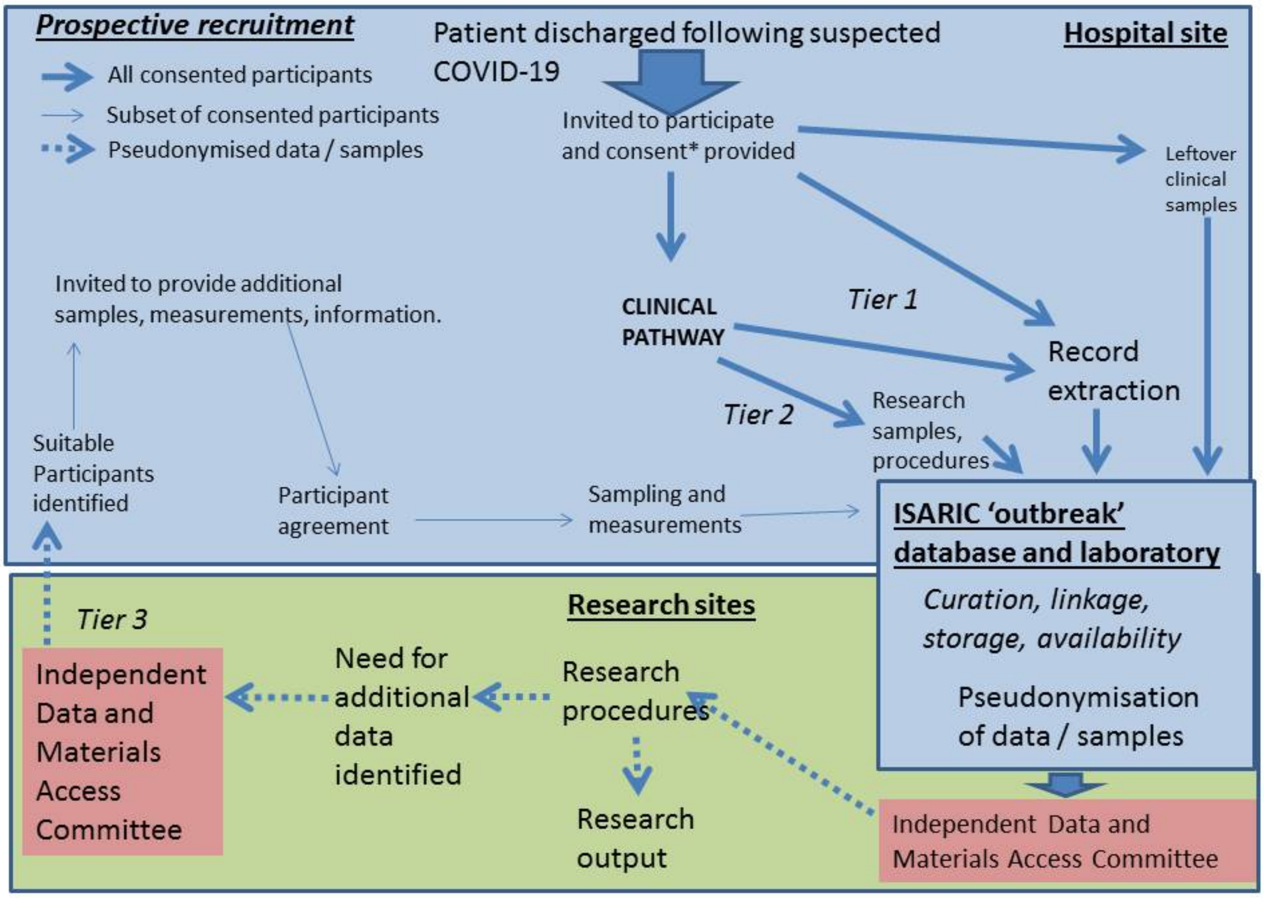

**Figure 5: Data extraction, curation, linkage and storage**

We will take appropriate steps to ensure that no other potentially identifiable data is included in the pseudonymised data set. For example, removal of identifiable meta-data from imaging scans, removal of names from free text fields.

A pseudonymised version of the clinical data set, and extra information or measurements undertaken for the study, will be reviewed by clinically-trained members of the research team prior to analysis. Variables will be analysed as quantitative, binary or categorical according to the specific research questions being addressed.

Participants who have already been recruited via ISARIC will re-consent to PHOSP-COVID19 and have their follow-up data linked to their acute phase data using ISARIC id and NHS number.

Established links with UK longitudinal population studies with epidemiological, biomarker (including COVID-19 serology) and genomic data (including but not limited to EXCEED, Coronagenes, UK Biobank) and national and global initiatives (International Complex Disease Alliance COVID-19 Host Genomics Initiative, ISARIC, NIHR Bioresource) will enable comparative studies and meta-analyses as outlined in Aim 2 above. Participants will provide consent to linkage of their data with other research studies that they have been recruited to.

‘TRUE COLOURS’ tool to record and monitor symptoms

We are working with the University of Oxford to use their ‘True Colours’ software. True Colours (TC) is a web and smartphone based digital tool to monitor a wide variety of symptoms and questionnaires including quality of life remotely. It’s been used as a clinical and research tool across a number of mental and physical health disorders (depression, bipolar disorder, inflammatory bowel diseases, psychosis, eating disorders, sleep disorders) developed over a decade ago by psychiatrists, software engineers, and researchers at the University of Oxford in partnership with Oxford health NHS Foundation Trust. TC prompts users to answer questionnaires rating their symptoms, via text or e-mail (participant preference) at set time points. Treating clinicians of TC users also have access to these plotted response summaries. TC is jointly governed between the University of Oxford and Oxford Health, with the University of Oxford currently responsible for information governance.

Using this software will enable participants to answer questions at a time more suited to them and with breaks rather than at research visits. Hard-copy questionnaires will be available for participants who are either not able or choose not to use the software.

##### *5.6 Analyses using the DNA*

Background to the DNA research methods: The human genome is made up of around 3 billion base pairs (commonly denoted by the letters A, C, G and T). Variations in the genome can involve (i) sequence variation, where there is a substitution of a base pair that usually occurs in a specific position by an alternative base pair (for example a substitution of A for G) or (ii) structural variation where a segment of DNA may be deleted, duplicated or inverted. Studies may focus on genes and genetic variants have been shown to be associated with traits that can occur as a consequence of COVID-19 or may analyse all genetic variation carried by each individual to identify new variants and genes of interest.

DNA will be extracted from cell fractions following plasma separation and/or from the saliva sample provided by the participant. Saliva should yield around 150-250ug of DNA (more than sufficient for the genetic assays proposed here) which will be stored as 3 aliquots.

The DNA samples will be genotyped using either microarray or re-sequencing technology. We will measure all genome-wide variation including, but not limited to, single base changes, structural changes, Human Leukocyte Antigen typing, Killer-cell Inhibitory Receptor (KIR) typing and epigenetic variation. During the period that the participants are recruited to the study it is likely that there will be further developments in genetic technologies and a reduction in prices of genetic assays. Similarly, the analytic strategies are keeping pace with the development of assays. The research team is involved in a range of large-scale genetic studies employing the latest technologies and will be well-placed to advise on modifications to the genetic assays (and to analysis approaches to the data generated by the assays) that will make best use of the samples and funding to maximise the prospects of discovery of causal genetic variants.

We will coordinate with other national studies (including Longitudinal Population Studies such as UK Biobank and dedicated COVID-19 cohorts such as Coronagenes and GenOMICC) to maximise funds available for whole genome sequencing, to link to additional data and to identify potential sample duplications in meta-analyses or replication studies.

These additional resources will also be a source of controls for specific research questions.

##### *5.7 Other samples*

All samples will be collected according to recognised guidelines, stored under appropriate conditions and in line with all relevant legislation, and tested and analysed using documented protocols and procedures. The Executive Board will regularly review the ongoing blood sample collection, storage, processing and analysis. Processing of samples for gene expression, epigenetics, telomere length, proteomics and other biomarkers will be subject to funding.

##### *5.8a Data-analysis of the semi-structured interviews*

Audio-recorded interviews will be conducted privately face to face or via telephone between the participant and an interviewer after informed consent. The interviews would be anticipated to be between 30 minutes and one in hour duration and will be professionally transcribed verbatim, with identifiable information removed. Interview prompts will be devised based upon relevant literature, experience of the team and consultation with patient representatives.

The interviews will be reviewed using thematic analysis supported by NVivo software. This approach follows six distinct stages: familiarisation with data; generating initial codes; searching for themes; reviewing themes; defining and naming themes and producing the report. Initial coding will be carried out and a sample of interviews will be coded by a second member of the team to ensure consistency and to enhance interpretive authenticity. Throughout the data analysis, an iterative approach will be undertaken with the research team meeting to discuss and review emerging themes and search for accounts that provide contesting views of the same phenomena or identify different phenomena. Patient representatives will be invited to comment on the emerging themes from the patient interviews to whether important issues may have been missed which could be included in subsequent interviews.

Audio recordings will be conducted using an encrypted Dictaphone. The recordings will be uploaded to secure drives on University of Leicester and University Hospitals of Leicester computers then deleted from the Dictaphone. Access to the files will be restricted and password protected. The transcription will be performed by an external company and a confidentiality agreement will be in place.

##### *5.8b Data-analysis of physical activity monitoring*

Data will be downloaded from the activity monitors at the recruiting site and then transferred to the NIHR Leicester BRC lifestyle team in line with their written standard operating procedure. Data will be processed to provide validated objective measures of sleep duration and quality, sedentary behaviour, time in different intensities of physical activity, as well as overall physical activity volume and intensity profiles using methods and standard operating procedures developed within the Lifestyle theme of NIHR Leicester BRC.

### 6. ELIGIBILITY

#### *6.1 Overall Description of Eligible Participants*

All patients who are admitted to an admissions unit or ward at UK recruiting hospital sites will be invited to join the study following discharge from hospital with clinician suspected COVID-19. This will include eligible individuals who were discharged from hospital before the start of the study (see Figure 4).

#### *6.2 Inclusion Criteria*

- Participant admitted to an admissions unit or ward at a UK hospital and discharged with suspected COVID-19
- Participant or consultee is willing and able to give informed consent for participation in the study
- Aged 18 years or above.

#### *6.3 Exclusion Criteria*

- Confirmed diagnosis of a pathogen unrelated to the objectives of this study and no indication or likelihood of co-infection with a relevant pathogen.
- Attendance at an A&E or emergency department only
- Refusal by participant, parent or appropriate representative.
- Other life limiting illness with life expectancy <6 months such as disseminated malignancy.

#### *6.4 Participation in other studies*

Patients who have already consented to COVID-19 related research (e,g., ISARIC-4c and RECOVERY trials) are eligible for inclusion. As this is an observational study participation in other studies is permitted, at the discretion of the investigators. The investigators will consider the burden of assessment of such participation for individual patients.

We will seek consent for linkage of all data and samples obtained for research purposes from participants who have or will provide consent to more than one COVID-19-related research study or longitudinal population study.

#### 7. STUDY PROCEDURES

##### *7.1 Informed Consent*

Patients will be provided with a copy of the Participant Information Sheet and consent form upon discharge from hospital or after healthcare professional contact within 12 months of discharge. Eligible participants will be identified by their normal clinical care team either directly by the participating site or via participant identification centres (PIC) sites. If identified via a PIC site and the patient is interested in taking part, then a member of the clinical care team will provide the patients details to the PHOSP study team at the local participating site, or the patient can contact the local study team directly using the contact details on the information sheet. Study information may be provided to potentially eligible participants either directly to the patient during a hospital/clinic visit or by post with an accompanying invitation letter. A phone number for contact for further information and the procedure for withdrawal is provided as part of the Participant Information Sheet. Study information, such as the study leaflet, may also be placed publicly available at participating sites and PIC sites (for example waiting areas).

Participants have the option to provide informed consent in one of four different ways:

- **Face-to-face** with a healthcare professional/member of the research team at a hospital appointment
- By **Telephone** with the support of a healthcare professional/member of the research team, or
- By **Post**
- By e-consent

We will not require potential participants to have been given 24 hours to review the Participant Information Sheet.

Patients choosing to provide informed consent by either the **face-to-face** or **postal** methods will sign a consent form that will be counter-signed by a member of the research team (at the same time or upon receipt if returned by post). A fully signed copy of the consent from will be provided to the participant for their records either at the time of their face-to-face consent, or by return post together with a copy of the withdrawal form.

NB. Postal Consent is the least preferred method of consent for PHOSP-COVID due to previously returned postal consent forms being incomplete and inaccurate. Any postal consent received at site must be checked thoroughly to ensure it is complete and accurate. Follow-up with participants may be required.

Patients choosing to provide consent by the **Telephone** method will have each statement on the Consent Form read to them by a member of the research team in conjunction with additional information about the study as required. The researcher will obtain verbal assent from the patient for each statement and then initial against each statement to confirm it has been understood and agreed to. The researcher will sign the consent form confirming that full verbal assent has been provided for the study, and a copy will be posted to the participant together with a copy of the withdrawal form.

Patients will also have the option to consent electronically via the e-consent system. All patients invited to join the study will receive details and instructions on the invitation letter, providing them with access to the e-Consent system. Patients choosing to provide consent by e-Consent will be asked to follow the URL: phosp.org/consent. There they will be asked to enter a 6 digit code (provided on the invitation letter), their first name, last name, year of birth and recruiting site (provided on the invitation letter). Error messages will appear if the invitation code/site is incorrect or the first name/last name is incorrect. All fields will need to be completed before they can proceed. This will take them to the e-consent platform where they will follow the instructions and complete the consent and some further questions. These additional questions are normally completed by the research team, but participants have the option to complete these via the e-consent platform. The electronic consent form will require the participant to confirm in the affirmative each mandatory statement from the consent form and provide an affirmative or negative response to the optional statements. At the end of the form the system will request the participant to provide an electronic signature (and witness/ translator signature if required) and then provide them with a downloadable pdf of their completed consent. If the patient chooses not to consent they will reach a page informing them that they chose not to take part. A section of the website will be available for participants to access using their 6 digit code where they can access a generic and current PIS and withdrawal form and their local hospital site contact details. Patients who complete the e-Consent will have their data automatically allocated to their appropriate recruiting site in the REDCap database, and an alert will go to a member of the appropriate study team notifying them that a patient has completed e-Consent, where it can be reviewed for completeness. If a patient is taking part, the patient can then be allocated a PHOSP ID by the recruiting site.

To generate the 6 digit codes, the PHOSP recruiting sites will go to phosp.org/redcap/phosp-econsent-invitation-code-generation. There they will upload an Excel file containing the details of patients who will be invited to use e-Consent for the PHOSP study. The Excel file requires patient first name, last name, and year of birth. The recruiting site also selects their site from the dropdown list and uploads the file. They will then be able to automatically download a csv file which contains a 6 digit code to be added to the patient's invitation letter.

Patients can opt-out of DNA collection and Tier 2 research data collection and still contribute Tier 1 research data.

At subsequent face-to-face visits, continued willingness to participate will be confirmed verbally by the research team and documented. Participants will have received a copy of the withdrawal form following informed consent, however, we will also host the withdrawal form on Phosp.org so it is accessible to all participants.

Patients who are unable to sign the consent form themselves either due to physical or language difficulties but are able to provide informed consent may do this through a **witness/translator** whose relationship to the participant should be documented on the consent form and who should sign the appropriate declaration on the consent form. A witness/translator can be a family member or trusted friend, member of the research team, a healthcare professional, or a routine hospital translator. For those participants who consented to the study via a witness/translator and are participating in any Tier 2 research visits, the study team will make every effort to ensure that the participant fully understands their involvement in the study and provides continued willingness to participate. This will be documented.

**Consultee Declarations** will be allowed for **Tier 1 recruitment only** for patients who do not have capacity (temporarily or permanently) to provide consent themselves. There are a number of different ways these participants will be identified:
1) a representative contacting the study team follow an invitation letter being sent to the patient

2) by a treating clinician contacting the study team
3) where permission is available to contact the patient directly but it becomes apparent to the study team that the patient may lack capacity (and this would then be clarified by a clinician)
A clinician experienced in assessing capacity will decide if a patient has capacity to provide consent. A Consultee can be a Personal Consultee or a Nominated Consultee. A **Personal Consultee,** who is someone who cares for the patient or is interested in their welfare, has a personal relationship with the patient and does not have a conflict of interest, such as being part of the research team or gaining financial benefit and is not acting in a professional capacity. Examples of suitable people who might act in this manner are a family member, caregiver or close friend. If a Personal Consultee is not available or is unable or unwilling to provide advice, then a **Nominated Consultee** can be approached. A Nominated Consultee is often a professional who is involved in the patient's care in a professional capacity but has no connection with the research study. These participants will not be eligible for any study procedures including DNA/RNA sample collection and relevant sections of the consent form will be marked as 'no' as directed by the instructions on the consent form. Only clinically collected data being used for participants who have not provided informed consent themselves. Participants who regain the capacity to consent for themselves will asked to provide full informed consent. Consultee Declaration will only occur for face to face or telephone consent.

##### *7.2 Baseline Assessments*

We will collect baseline information that will include, but not be limited to:

- Patient details (e.g. name, NHS number, date of birth, biological sex, address and postcode)
- Demographic details (self-reported ethnicity, household income, gender, education level, marital status, disabilities, and exposure details). This questionnaire will be provided to participants following informed consent. This may be asked verbally face to face or over the telephone or provided as a self-completed paper questionnaire during visits or posted to participants for completion and return alongside postal consent forms. Clinician details (e.g. name)
- COVID-19 symptom onset date
- COVID-19 results of throat, nasal swab, or sputum sample, or serology from blood tests
- Pre-existing comorbidities (e.g. heart disease, diabetes, chronic lung disease, chronic kidney disease)
- Pre-existing immunosuppressants Y/N
- COVID-19 severity as assessed by need for supplemental oxygen or ventilation/extracorporeal membrane oxygenation
- Other organ support required during hospitalisation such as renal replacement therapy
- Anti-coagulant during COVID-19 hospitalisation Y/N
- Antibiotic during COVID-19 hospitalisation Y/N
- Steroid during COVID-19 hospitalisation Y/N
- Date of hospitalisation
- Date of discharge
- Name of person completing the form

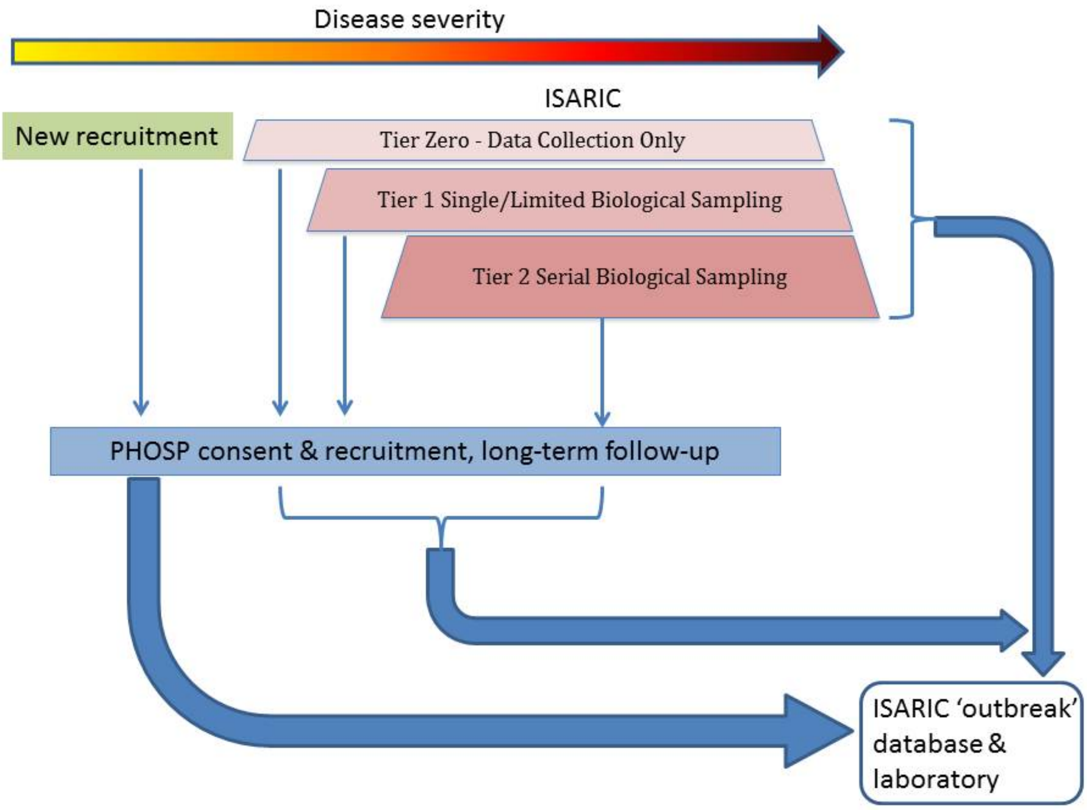

**Figure 6: PHOSP and ISARIC recruitment - linked data collection and repository**

##### *7.3 Subsequent Assessments*

We will undertake automated repeat extraction of routine healthcare records (for example, NHS Digital) for all participants every 3 months after enrolment into the study for the first 12 months and every 6 months thereafter. Extraction time-points might vary by up to 3 months. Participants who attend additional clinic visits will have information recorded from their clinical visits as described above (section 5.2). Participants who agree to additional research procedures and sampling (Tier 2) and to additional research studies (Tier 3) will have those data recorded (as described above – sections 5.3 and 5.4). See Figure 1.

##### *7.4 Recruitment and Follow-up*

Recruitment will continue for one year (although current funding will be for six months). For all participants, we will continue to link to their clinic data for at least 25 years or until the patient has deceased or withdrawn consent.

Following recruitment of the first 1000 participants, we will undertake a review to determine if key populations are under-represented. Strategies needed to address under-representation will be developed and agreed by the Executive Board (Figure 7).

##### *7.5 Discontinuation/Withdrawal of Participants from Study Treatment*

Each participant has the right to withdraw from the study at any time by completing a withdrawal form (this could be face to face, phone or post and online at Phosp.org). In addition, the investigator may discontinue a participant from the study at any time if the investigator considers it necessary for any reason including:

- Ineligibility (either arising during the study or retrospective having been overlooked at screening)
- Significant protocol deviation
- Consent withdrawn
- Lost to follow up (i.e. moved to a different geographical area and no longer attending the local clinics for their conditions).
- Death

The reason for withdrawal will be recorded in the CRF. The withdrawal form can be completed by someone acting on behalf of the participant.

If participants withdraw then any data and samples already collected will remain and be used in the study. Information and data will continue to be collected about participants health from central NHS records, hospital records and from participants GP unless participants state otherwise on the withdrawal form.

##### *7.6 Source Data*

The source data will be hospital records (from which medical history and previous and concurrent medication information will be extracted), demographic data, health and social care records (e.g. NHS Digital, primary care health records), clinical and office charts including 'True Colours', laboratory and pharmacy records, and imaging. Source data also includes data from linked research studies.

All documents will be stored safely in secure office environments with access restricted to delegated members of the research study teams at each of the participating sites. On all study-specific documents, other than the signed consent and withdrawal forms, the participant will be referred to by the study participant number/code, not by name. For any questionnaires that are posted out, the default will be for them to be labelled by study ID only. If there are two study participants in the household the questionnaires will be named for completion but the names would be removed before filing in their research file.

#### 8. SAFETY REPORTING

As this is an observational study an adverse event in this protocol is defined as any untoward medical occurrence in a patient or clinical investigation that has occurred from research interventions only. Therefore, we will only report adverse events and serious adverse events that relate directly to participation in the study (for example, as a consequence of providing additional blood samples or physical activity monitoring). All adverse events (AEs) and serious adverse events (SAEs) will be recorded on an AE/SAE log in the ISF to capture start and end dates.

Any clinically significant results as part of the research will be shared with the participant's lead clinician or general practitioner for further action as required with the exception of results from DNA analysis (see Section 11.1 below).

### 9. STATISTICS

#### *9.1 Description of Statistical Methods*

Analyses will be performed using standard epidemiological and statistical genetics methodology. This will include cross-sectional and longitudinal studies, and analyses of disease prevalence and incidence.

Analysis design and choice of controls will be dependent upon the precise nature of the research question. Identification of risk factors for a specific COVID-19 sequela would involve controls both from within PHOSP-COVID (without the sequela) and serology-positive controls with prospective questionnaire and healthcare record linkage from Longitudinal Population Studies (for example, UK Biobank, Coronagenes, EXCEED) and pre-existing disease cohorts.

Analyses aiming to characterise and understand the clinical features, subtypes and trajectories of sequelae (for example, sarcopaenia) would evaluate the cross-sectional and longitudinal clinical data and biomarkers of PHOSP participants who are presenting with the sequelae being studied.

Analyses aiming to understand the role and function of particular biomarkers (for example, genetic markers, raised cytokine levels) would conduct ‘phenome-wide’ analyses of that biomarker using the full PHOSP dataset.

The PHOSP-COVID study itself is also intended as a source of cases and controls for other studies. For example, as a comparator group for studies of critical care of non-COVID-19 disease, or for studies of sequelae identified amongst non-hospitalised COVID-19 survivors. Our research Tier 3 capability enables recall of PHOSP-COVID study participants for case-control studies or nested cohort studies based on either pre-existing risk factors (for example, age, demographics, biomarkers, comorbidities) or disease status itself (for example, post-COVID-19 cardiovascular complications). PHOSP-COVID study participants are asked for consent to linkage with other research studies to which they are recruited to reduce sample overlap.

We have extensive experience of development of, and collaborative use of, disease-specific and general population cohort studies both nationally and internationally enabling access to control populations and alignment of research strategies for rapid validation and replication of findings.

#### *9.2 The Number of Participants*

We aim to recruit 10,000 individuals, who were discharged from hospital with suspected COVID-19. This will include at least 4,000 individuals that participate in Tier 2 samples and data collection.

We aim to study the impact of COVID-19 upon respiratory, cardiovascular, deconditioning and mental health. Without prior knowledge of the incidence of new chronic disease as a consequence of COVID-19 in post-hospitalised survivors it is not possible to precisely estimate whether these number of participants are sufficient to identify key associations between the development and persistence of chronic disease and demographic, clinical and molecular biomarkers of susceptibility. However, data in the small cohorts reported do provide some insights, for example, of the possible development of lung fibrosis. In SARS, abnormal diffusion capacity (a physiological measure indicating possible underlying fibrosis) was identified in ~25% of hospitalised cases, and data from China suggests a similar decrement in diffusion capacity in COVID-19.

A sample size of 4,000 will allow an equal discovery and validation subsets in the same study to explore fibrosis risk versus non-risk using a binary regression model. This model will have 80% power to detect odds ratios >1.1 assuming the multiple correlation coefficient $\rho$ to be approximately 0.4, the event rate to be 0.25. A sample size of 500 will have 80% power to detect an odds ratio >1.5. Thus, we are confident that we will have sufficient power to identify important associations between risk biomarkers and development of different chronic diseases in important subgroups e.g. ethnicity or those receiving specific interventions during the acute infection.

Where recall studies are proposed (Research Tier 3), the Executive Board will review experimental design and power calculations to ensure adequate sample sizes are included and proposals will be approved by the External Scientific Advisory Board (Figure 7) and will undergo ethical review as appropriate.

##### *9.3 The Level of Statistical Significance*

Statistical significance thresholds will be defined in advance of each analysis and will take into account issues of multiple testing and *a priori* evidence.

##### *9.4 Procedure for Accounting for Missing, Unused, and Spurious Data.*

All data sets will undergo rigorous quality control prior to undertaking analyses. Outlying values will be investigated in the raw data and excluded as appropriate. Missing data will either be imputed, or individuals removed from analyses, as appropriate for the particular analysis being undertaken.

Direct access will be granted to authorised representatives from the sponsor, host institution, and the regulatory authorities to permit study-related monitoring, audits and inspections.

Data extraction will be performed by a study team member for all patients who have provided consent.

#### 10. QUALITY CONTROL AND QUALITY ASSURANCE PROCEDURES

The study will be conducted in accordance with the current approved protocol, ICH GCP, relevant regulations and standard operating procedures.

#### 11. CODES OF PRACTICE AND REGULATIONS

##### *11.1 Ethics*

###### ***Rapidly evolving genetic assay technologies***

During the period that the participants are recruited to the study it is likely that there will be further developments in genetic technologies and a reduction in prices of genetic assays. Similarly, the analytic strategies are keeping pace with the development of assays. The research team is involved in a range of large-scale genetic studies employing the latest technologies and will be well-placed to advise on modifications to the genetic assays (and to analysis approaches to the data generated by the assays) that will make best use of the samples and funding to maximise the prospects of discovery of causal genetic variants.

###### ***Success of the project and implications for care of participants***

There will be no implications for care of participants. Inclusion in the study is entirely voluntary and they can withdraw at any time. Our findings could advance clinical knowledge of COVID-19 sequelae, which could ultimately lead to the development of a new predictive tests or treatments. Participants can agree to be re-contacted for future research studies, which might include clinical trials. These future studies will be subject to appropriate ethical review prior to any re-contact.

###### ***Risk to subjects***

Participants may have blood drawn more often than is required for standard care. Phlebotomy can be associated with pain at the draw site and rarely with infection. Discomfort will be minimised by having qualified and experienced staff obtain blood samples, and by combining research sampling with routine clinical sampling.

###### ***DNA storage***

Stored DNA samples, and possibly cell lines to provide a renewable DNA source, will be stored indefinitely for future research use. Cell lines may be deposited at the end of the research in a cell-line bank so as to be available to other qualified researchers. The consent we seek and this application relates to (i) future research studies aimed at investigating how genetic variations can contribute to risk of developing respiratory disease (for example, if new improved laboratory methods become available) and; (ii) use of the samples or data for comparison purposes when we are studying different diseases. Further research proposals from members of the study team or from investigators and commercial companies both within and outside of the UK will be reviewed by the Executive Board and approved by the External Scientific Advisory Board (Figure 7) and will undergo ethical review as appropriate.

###### ***Sharing of genetic and clinical information and samples***

To improve research in this area, we may share the samples, genetic information and basic clinical information with *bona fide* researchers in different organisations, including those in other countries or in commercial organisations. Appropriate Data and Material Transfer Agreements will be put in place prior to any sharing of data and samples. All relevant organisational and technical measures will be taken to safeguard peoples information. Consent is the legal basis for the transfer of this information.

##### *Incidental findings in genetic testing*

This study includes genetic testing to identify host genetic variants associated with disease progression or severity. There is a very small chance that these tests may result in the incidental discovery of information that is relevant to the participant's health. Since the samples will be analysed anonymously in batches, and generally in non-clinical laboratories with investigational techniques, we will not attempt to identify and inform participants of any results from genetic tests. If we were to do so, there would be a considerable risk of accidental harm in the form of unnecessary anxiety and distress.

##### *Participant Burden*

Participants may be asked to undertake additional procedures that are not requested across all participants (for example, airway sampling). Moreover, Tier 3 research studies that re-contact participants for additional research procedures may be targeted towards specific subsets of participants (for example, the most severe). Our database will include a regularly updated mechanism to record additional requests made, and additional studies participated in, within the PHOSP-COVID study framework. The PHOSP-COVID study will provide a broad sampling frame for all studies, identify participants who are likely to become overburdened, and identify participants that may already have the required information available for subsequent studies. To monitor the research requests made on participants selected to be invited to join additional studies, (Tier 3), proposals will be reviewed by the Executive Board and approved by the External Scientific Advisory Board (Figure 7) and will undergo ethical review as appropriate.

#### *11.2 Standard Operating Procedures*

All relevant Sponsor SOPs will be followed to ensure that this study complies with all relevant legislation and guidelines. Where relevant, local NHS SOPs will be followed at participating sites

#### *11.3 Declaration of Helsinki*

The Investigator will ensure that this study is conducted in full conformity with the current revision of the Declaration of Helsinki (last amended October 2000, with additional footnotes added 2002 and 2004).

#### *11.4 ICH Guidelines for Good Clinical Practice*

The Investigator will ensure that this study is conducted in full conformity with relevant regulations and with the ICH Guidelines for Good Clinical Practice (CPMP/ICH/135/95) July 1996.

#### *11.5 Approvals*

Once Sponsor authorisation has been confirmed, the protocol, informed consent form, participant information sheet and any proposed advertising material will be submitted to an appropriate Research Ethics Committee (REC), Health Research Authority (HRA), and host institution(s) for written approval. Sponsor Green Light will be issued to each site upon receipt of all the relevant approvals.

Once Sponsor authorisation has been confirmed, the Investigator will submit and, where necessary, obtain approval from the above parties for all non-substantial and substantial amendments to the original approved documents. Sponsor Green Light will be issued to each site upon receipt of all the relevant approvals

##### *11.6 Participant Confidentiality*

The research staff at each participating site will ensure that confidentiality is preserved. Participants will be identified only by initials and a participants ID number on the CRF and any electronic database. All documents will be stored in secure office environments at each participating site, with access restricted to delegated members of the research team and authorised personnel from the Sponsor (University of Leicester) or their delegate, the host NHS organisation, regulatory authorities, or public health agencies and members of the coordinating study team from the participating site for monitoring and auditing purposes. The study will comply with the Data Protection Act (2018) and General Data Protection Regulation (2018) which requires data to be anonymised as soon as it is practical to do so.

##### *11.7 Other Ethical Considerations*

**Recruitment of ill patients** Patients will be asked to consent at the point they are deemed well enough to be discharged from hospital. Some discharged patients may still be physically unwell, but no longer requiring hospital treatment. Patients can provide consent upto 12 months after discharge for Tier 1 and 3, but only until 6 months post discharge for Tier 2.

**Perceived coercion because of individual responsibilities to society, and the implications of this research for public health.** We are sensitive to the fact that some patients, or their family and friends, may feel under an unusually strong moral obligation to participate (or encourage participation) given the nature of this research and the wide, and often inaccurate, publicity surrounding emerging infections. In view of this, we have tried to make both the potential benefits and limitations of this simple observational study clear in the information sheet and declining to participate in the study would not affect on going clinical care. In the informed consent form we also stress that participation is entirely voluntary.

**Balance between public health and research.** Patients with emerging infections are commonly the subject of public health investigations. The work proposed here is research and will be clearly presented as such. There is no primary gain to the patient from participating. In designing and describing this research we are clear that, in accordance with the guiding principles of Good Clinical Practice, the needs and autonomy of the individual are paramount and the potential benefits to wider society do not take precedence.

**Risks to clinical and research staff treating the participants** As all contact with participants for up to 3 months post-discharge are expected to be by telephone or under usual clinical circumstances with appropriate precautions, we do not expect additional risk to staff as a consequence of the research activities. We expect that participants will no longer be infectious 3 months after discharge. If new evidence is reported that suggests there is a longer infectious period after onset of symptoms, or that individuals may be re-infected within this time, we will take appropriate steps to ensure the safety of staff and participants.

#### 12. DATA HANDLING AND RECORD KEEPING

Paper records (e.g. CRFs, consent forms etc) will be kept at the recruiting sites in a secure physical location. Research data from each site will be transcribed into a single dedicated REDCap database system hosted on a secure systemThis will include eCRFs and a visit/event structure which are based on the study design described in section 5. A modular approach will be used to collect data from across the different tiers and disciplines by setting up additional REDCap projects. Day-to-day the REDCap system will be managed by the host site. At regular intervals, baseline data from consented ISARIC patients will be pre-loaded into the REDCap project to assist with the harmonisation of data across the two studies and to minimise collection of aligned data.

To manage the posting of sample kits to participants, minimal contact data (participant ID number, name and address) from tier 2 and tier 3 patients will be shared securely with the delivery partner. This will not include study data (e.g. clinical observations or linked health data). Contact data will be stored securely at the delivery partner with appropriate access control. In the lab information management system, samples will only be identified through their participant ID number and a unique sample ID. Full records of sample storage and shipment will be retained for monitoring and auditing purposes.

Data may be manually entered or uploaded using the facilities provided by REDCap (e.g. automated data import, bulk data upload). Where data import approaches are used, data will be reviewed/cleaned prior to initial run to ensure that identifiable information is not included in data transfers.

Data quality will be maintained using tools within the REDCap environment and through dashboards provided by the REDCap host site. These will ensure data quality at individual sites is maintained and allow early notification of problems. Regular reviews of the data quality will be undertaken by dedicated PHOSP team members..

Access to REDCap will be provided to users at an appropriate level, with roles setup accordingly. Data access groups (DAGs) will be used to maintain separation of data entry and exports across multiple sites. DAGs will be setup to mirror those used in the ISARIC study in order to make it easier to manage users and data across the two aligned studies.

On a regular basis, automated routines will extract eCRF data from REDCap and link it to routinely collected health care data (extracted using the NHS number if provided). For those participants for which it is available, data sets from the True Colours web site/mobile application will also be extracted and linked using participants' NHS number.

Images will be pseudonymised at the individual sites. Images will be archived by the University of Oxford and Royal Surrey County Hospital NHS Foundation Trust and made available to study members for download and linkage to other data sets.

In the short term, data will be accessed by bona fide researchers outside of the study team via application to the Executive Board and Steering Committee with oversight by the External Scientific Advisory Board.

Data will be retained for 25 years following the completion of the study, with an anonymised set stored indefinitely.

#### 13. DATA AND SAMPLE ACCESS AND SHARING

Requests for the use of data and samples will be reviewed by the Executive Board and Steering Committee with oversight by an External Scientific Advisory Board (ESAB).

The ESAB will comprise senior, independent people who will not be involved in the acquisition or analysis of PHOSP-COVID samples or data.

##### *Longer-term data availability*

In the longer term, a copy of the data set will be held by the HDR UK Digital Innovation Hub BREATHE at the SAIL Data Bank (Swansea) or a similar trusted research environment with access governed by a Data Access Committee.
