## Supplementary Material for "Physical, cognitive and mental health impacts of COVID-19 following hospitalisation – a multi-centre prospective cohort study"

**Supplement**

**Supplementary Methods**

**Table SM1. Outcome measures for participants included in the current analysis.**

| **Module** | **Common dataset including detailed clinical phenotyping** |
| --- | --- |
| **Symptoms** | Patient symptom questionnaire (PHOSP-COVID study specific questionnaire)  Dyspnoea12 Questionnaire  Fatigue scale Questionnaire (FACIT)  Brief Pain Inventory Questionnaire (BPI) |
| **Health-related Quality of life** | Euroqol EQ5D-5L |
| **Respiratory** | Pulmonary Function Tests Including: Spirometry (FEV_1_, FVC) and Transfer Factor (TLCO, KCO) |
| **Cardiac** | Blood tests: BNP / NT-Pro-BNP and Troponin I /Troponin T |
| **Renal** | Blood tests: eGFR |
| **Pre-diabetes/diabetes** | Blood tests: HbA1C levels |
| **Haematological** | Blood tests: D Dimer |
| **Systemic inflammation** | Blood test: CRP |
| **Physical performance** | Incremental Shuttle Walk Test (ISWT) to assess exercise capacity  Short Physical Performance Battery (SPPB) |
| **Frailty** | Rockwood Clinical Frailty Scale (CFS) |
| **Body composition** | Body Mass Index (BMI) calculation from Height and Weight Measurement |
| **Mental Health** | Generalised Anxiety Disorder Questionnaire (GAD-7)  Patient Health Questionnaire (PHQ-9)  Post Traumatic Stress Disorder Checklist for DSM-5 Questionnaire (PCL-5) |
| **Cognition** | Montreal Cognitive Assessment (MoCA) |

FEV_1_ = Forced Expiratory Volume in 1 second, FVC = Forced Vital Capacity, TLCO = Transfer Capacity of the Lung for Carbon Monoxide, KCO = carbon monoxide transfer coefficient, BNP = Brain Natriuretic Peptide or NT-BNP N-Terminal Brain Natriuretic Peptide, HbA1C = glycosylated haemoglobin, eGFR = estimated Glomerular Filtration Rate, CRP = C-Reactive Protein

**Table SM2. Methods and thresholds for processing of variables and outcome measures used in the current analysis.**

|  | **Method** |
| --- | --- |
| **Table 2** |  |
| Symptoms at 2 to 7 months | The total number of current symptoms reported were from the following list which were answered as binary Yes/No questions: Aching in your muscles (pain), Physical slowing down, Slowing down in your thinking, Joint pain or swelling, Limb weakness, Difficulty with concentration, Short term memory loss, Headache, Tingling feeling/pins and needles, Confusion/fuzzy head, Dizziness or light headedness, Chest tightness, Problems with balance, Altered personality/ behaviour, Chest pain, Palpitations, Leg/ankle swelling, Difficulty with communication, Skin rash, Diarrhoea, Problems seeing, Pain on breathing, Weight loss, Tremor/shakiness, Constipation, Erectile Dysfunction, Loss of sense of smell, Can’t fully move or control movement, Abdominal pain, Stomach pain, Loss of control of passing urine, Loss of appetite, Loss of taste, Nausea/vomiting, Bleeding, Can’t move and/or feel one side of your body or face, Loss of control of opening bowels, Lumpy lesions on toes, Fainting / blackouts, Seizures  Symptom severity was rated using a 0-10 visual analogue scale for Breathlessness, Cough, Fatigue, Sleep quality and Pain before COVID-19 illness and worst in last 24 hours. The results were dichotomised using cut off of ≤2 for no and ≥3 for Yes to combine the analysis with the longer list of symptoms. For the analysis shown in Table SR6 section a) a lower score by 1 point was reported as patient worsened. |
| Generalised Anxiety Disorder Questionnaire (GAD-7) (Anxiety) | The Generalised Anxiety Disorder (GAD-7) questionnaire is a patient reported outcome measure consists of 7 questions with total scores ranging from 0 to 21. We used a GAD7 threshold score of > 8 to suggest at least mild-moderate anxiety.^1^ |
| Patient Health Questionnaire (PHQ-9) (Depression) | The Patient Health Questionnaire (PHQ-9) is a patient reported outcome measure consisting of 9 questions with total scores ranging from 0 to 27. We used a PHQ-9 threshold score of ≥10 to suggest at least moderate depression.^2^ |
| Post-Traumatic Stress Disorder Checklist for DSM V (PCL-5) Questionnaire | The Post-Traumatic Stress Disorder Checklist for DSM V (PCL-5) questionnaire is a patient reported outcome measure consisting of 20 questions assessing evidence of post-traumatic stress disorder according to the DSM V criteria. Total scores range from 0-80. We used a PCL-5 threshold score of ≥38 suggestive of a provisional diagnosis of post-traumatic stress disorder.^3,4^ |
| Dyspnoea-12 | The Dyspnoea-12 questionnaire is a patient reported outcome measure consisting of 12 questions assessing breathlessness severity incorporating both “physical” and “affective” aspects.^5^ Scores range from 0 to 36 with higher scores correspond to greater severity of breathlessness. |
| FACIT fatigue subscale score (FACIT) | The Functional Assessment of Chronic Illness Therapy – Fatigue (FACIT-Fatigue) scale is a patient reported outcome measure consisting of 13 questions to assess self-reported fatigue and its impact on daily activities and function.^6^ Total scores range from 0-52, with lower scores corresponding to an increased burden of fatigue.^7^ |
| Brief Pain Inventory (BPI) severity and interference | The Brief Pain Inventory (BPI) is a patient reported outcome questionnaire consisting of 15 questions across domains of pain severity and pain interference. We have reported the BPI Severity score as the mean score from the 4 severity questions each with a range 0 – 10 anchored at 0 = “No Pain” and 10 = “Pain as bad as you can imagine”.^8,9^ |
| Short Physical Performance Battery (SPPB) | The Short Physical Performance Battery (SPPB) test is a researcher administer assessment of physical performance and frailty. It comprises 3 components; balance, gait speed and sit to stand tests. Tests were completed according to recommended standards and training was provided to site staff by the central study team via a recorded demonstration video. SPPB total scores range from 0-12. We have reported a total SPPB score of ≤10 suggestive of underlying frailty.^10^ |
| Incremental Shuttle Walk Test (ISWT) | The Incremental Shuttle Walk Test (ISWT) is a researcher administered assessment of maximal physical performance and was performed according to standardised instructions with two attempts performed by participants on the same day with a 20 minutes rest between them.^11^ Training was provided to site staff by the central study team via a recorded demonstration video. The best effort was reported in metres and the percent predicted value was calculated using the following reference formula accounting for gender, age and BMI.^12^ (ISWTpred = 1449.701 − (11.735 × age) + (241.897 × gender) − (5.686 × BMI), where male gender = 1 and female gender = 0) |
| Rockwood Clinical Frailty Scale (CFS) | The Rockwood Clinical Frailty Scale (CFS) is a researcher assessed scale of clinical frailty with scores ranging from 1-9 where lower scores correspond to increased frailty. We have reported CFS scores of <5 suggestive of frailty.^13^ |
| Montreal Cognitive Assessment (MoCA) | The Montreal Cognitive Assessment (MoCA) is a researcher administered cognitive function questionnaire across 8 domains. Training was provided to site staff using standardised resources supplied online by MoCA TEST Inc.^14^ The assessment was conducted in English with researchers applying their discretion to exclude participants whose command of English was insufficient to complete the test accurately. Total scores range from 0 to 30. We report total MoCA scores of <23 suggestive of at least Mild Cognitive Impairment.^15^ |
| Spirometry and Pulmonary Function Testing | Spirometry and Pulmonary function testing was completed as per ERS/ATS recommendations.^16^ Spirometry and Transfer factor values were converted to SI units if not reported as such by sites. ERS Reference values were used to calculate % predicted values.^17-19^ FEV_1_/FVC <0.7 was used to define airflow obstruction.^20^ % predicted TLCO <80% was considered indicative of impaired gas transfer. |
| BNP / NT-pro BNP | Brain Natriuretic Peptide (BNP) and N-terminal pro B-type Natriuretic Peptide (NT-pro BNP) were collected by according to each site’s routine clinically available assay as a biomarker of heart failure. Three sites submitted BNP results with all of the remaining sites submitting NT-pro BNP results. The threshold values used for BNP was ≥ 100 ng/litre^21^ and for NT-pro BNP ≥ 400ng/litre ^22^ as suggestive of heart failure. |
| Glycated haemoglobin (HbA1c) | Glycated haemoglobin (HbA1c) was collected as a biomarker of current glycaemic control. We have reported HbA1c levels ≥ 6.5% as suggestive of a diagnosis of diabetes.^23^ |
| D-Dimer | D-Dimer levels were collected as a biomarker of possible thromboembolic disease with sites reporting results either as mcg/mL Fibrinogen Equivalent Unit (FEU) or ng/mL D Dimer Unit (DDU) according to their clinically available assays. Conversion of mcg/mL FEU to ng/mL DDU used the following equation: (value in mcg/mL FEU)*500  We have reported a D-Dimer level ≥ 500ng/ml as suggestive of systemic inflammation and possible venous thromboembolic disease.^24^ |
| C-Reactive Protein (CRP) | C-Reactive Protein (CRP) levels were collected as a biomarker of current systemic inflammation. Values reported as below the lower or upper limit reportable range for the assay used at the site have been included at the stated less than or more than cut off value for calculation of mean (SD) results. We have reported CRP levels > 10mg/L as suggestive of systemic inflammation. |
| **Table 5** |  |
| EQ5D-5L VAS | The EQ5D Visual Analogue Scale is a patient reported outcome questionnaire recording the patient’s self-rated health and was completed for “before your COVID-19 illness” and “your own health state today.” Scores are presented as mean and standard deviation.^25^ |
| EQ5D-5L Utility Index | The EQ5D-5L is a five-dimension patient reported outcome questionnaire recording a patient’s self-rated health state for mobility, self-care, usual activities, pain/discomfort and anxiety/depression. These scores are then mapped to a United Kingdom specific Utility Index anchored at 1 for “perfect health” and 0 for “dead” calculated from reported EQ5D-5L scores across the five dimensions.^26^ |
| Washington Group Short Set of Functioning | The Washington Group Short Set of Functioning (WG-SS) is a patient reported outcome questionnaire using six questions to assess disability and function. A participant was considered to have a new disability if a response for any single domain changed from “no difficulty” or “some difficulty” to “a lot of difficulty” or “cannot do it at all” following the Washington Group guidelines.^27^ |

**Supplementary Data – Results**

**Table SR1.** **Co-morbidities for the cohort stratified by severity of acute illness using the WHO clinical progression scale.**

|  | WHO – class 3-4 | WHO – class 5 | WHO – class 6 | WHO – class 7-9 | Total |
| --- | --- | --- | --- | --- | --- |
| Total N (%) | 226 (21·0) | 378 (35·1) | 185 (17·2) | 288 (26·7) | 1077 |
| **CARDIOVASCULAR DISEASE** | ·· | ·· | ·· | ·· | ·· |
| Myocardial Infarction | 11 (5·0) | 20 (5·5) | 3 (1·7) | 7 (2·5) | 41 (3·9) |
| Ischaemic Heart Disease | 14 (6·4) | 27 (7·4) | 5 (2·9) | 16 (5·7) | 62 (6·0) |
| Atrial Fibrillation | 10 (4·5) | 17 (4·7) | 14 (8·1) | 7 (2·5) | 48 (4·6) |
| Hypertension | 56 (25·5) | 126 (34·5) | 62 (35·8) | 110 (39·3) | 354 (34·1) |
| Congestive Heart Failure | 4 (1·8) | 5 (1·4) | 2 (1·2) | 4 (1·4) | 15 (1·4) |
| Congenital Heart Disease | 0 (0·0) | 2 (0·5) | 1 (0·6) | 2 (0·7) | 5 (0·5) |
| Valvular Heart Disease | 1 (0·5) | 6 (1·6) | 5 (2·9) | 2 (0·7) | 14 (1·3) |
| Pacemaker / Implantable Defibrillator | 6 (2·7) | 5 (1·4) | 1 (0·6) | 2 (0·7) | 14 (1·3) |
| Peripheral Vascular Disease | 4 (1·8) | 5 (1·4) | 1 (0·6) | 2 (0·7) | 12 (1·2) |
| Hypercholesterolaemia/dyslipidaemia | 25 (11·4) | 74 (20·3) | 39 (22·7) | 45 (16·1) | 183 (17·7) |
| Cerebrovascular Accident/ Transient Ischaemic Attack | 8 (3·7) | 23 (6·4) | 7 (4·1) | 13 (4·7) | 51 (4·9) |
| **NEUROLOGICAL and PSYCHIATRIC** | ·· | ·· | ·· | ·· | ·· |
| Dementia | 0 (0·0) | 4 (1·1) | 0 (0·0) | 0 (0·0) | 4 (0·4) |
| Depression or Anxiety | 32 (14·6) | 45 (12·3) | 29 (16·8) | 40 (14·3) | 146 (14·1) |
| Chronic Fatigue Syndrome/fibromyalgia or chronic pain | 10 (4·6) | 11 (3·0) | 5 (2·9) | 12 (4·3) | 38 (3·7) |
| Previous treatment with antidepressant medication | 21 (9·6) | 34 (9·3) | 23 (13·4) | 24 (8·6) | 102 (9·9) |
| Previous treatment with a mental health professional for a mental health problem | 12 (5·5) | 14 (3·8) | 13 (7·6) | 13 (4·7) | 52 (5·0) |
| **RESPIRATORY** | ·· | ·· | ·· | ·· | ·· |
| COPD | 10 (4·6) | 23 (6·3) | 11 (6·4) | 5 (1·8) | 49 (4·7) |
| Asthma | 41 (18·6) | 64 (17·6) | 36 (20·8) | 47 (16·8) | 188 (18·1) |
| Interstitial Lung Disease | 1 (0·5) | 3 (0·8) | 1 (0·6) | 3 (1·1) | 8 (0·8) |
| Bronchiectasis | 7 (3·2) | 5 (1·4) | 4 (2·3) | 3 (1·1) | 19 (1·8) |
| Obstructive Sleep Apnoea | 8 (3·6) | 20 (5·5) | 11 (6·4) | 14 (5·0) | 53 (5·1) |
| Obesity hypoventilation syndrome | 0 (0·0) | 0 (0·0) | 2 (1·2) | 2 (0·7) | 4 (0·4) |
| Pleural Effusion | 1 (0·5) | 7 (1·9) | 0 (0·0) | 2 (0·7) | 10 (1·0) |
| **RHEUMATOLOGICAL** | ·· | ·· | ·· | ·· | ·· |
| Connective Tissue Disease | 1 (0·5) | 0 (0·0) | 0 (0·0) | 0 (0·0) | 1 (0·1) |
| Rheumatoid Arthritis | 8 (3·6) | 9 (2·5) | 5 (2·9) | 5 (1·8) | 27 (2·6) |
| Osteoarthritis | 18 (8·2) | 39 (10·7) | 14 (8·1) | 25 (8·9) | 96 (9·3) |
| **GASTROINTESTINAL** | ·· | ·· | ·· | ·· | ·· |
| Peptic Ulcer Disease | 2 (0·9) | 4 (1·1) | 0 (0·0) | 2 (0·7) | 8 (0·8) |
| Liver disease *- Mild* | 4 (1·8) | 4 (1·1) | 5 (2·9) | 3 (1·1) | 16 (1·5) |
| Liver disease *– mod/severe* | 5 (2·3) | 3 (0·8) | 5 (2·9) | 4 (1·4) | 17 (1·6) |
| GORD | 23 (10·5) | 38 (10·4) | 15 (8·7) | 27 (9·7) | 103 (10·0) |
| Inflammatory Bowel Disease | 3 (1·4) | 6 (1·6) | 2 (1·2) | 2 (0·7) | 13 (1·3) |
| Irritable Bowel Disease | 9 (4·1) | 3 (0·8) | 7 (4·1) | 7 (2·5) | 26 (2·5) |
| **METABOLIC/ENDOCRINE/RENAL** | ·· | ·· | ·· | ·· | ·· |
| Diabetes *Type 1* | 1 (0·5) | 6 (1·6) | 0 (0·0) | 1 (0·4) | 8 (0·8) |
| Diabetes *Type 2* | 30 (13·7) | 80 (21·9) | 40 (23·1) | 63 (22·7) | 213 (20·6) |
| *Uncomplicated (% of all Diabetes)* | *21 (67·7)* | *58 (67·4)* | *25 (62·5)* | *39 (60·9)* | *143 (64·7)* |
| *End-organ damage (% of all Diabetes)* | *0 (0·0)* | *2 (2·3)* | *2 (5)* | *3 (4·7)* | *7 (3·2)* |
| Hypothyroidism | 15 (6·8) | 16 (4·4) | 2 (1·2) | 14 (5·0) | 47 (4·5) |
| Hyperthyroidism | 1 (0·5) | 9 (2·5) | 1 (0·6) | 3 (1·1) | 14 (1·3) |
| Chronic kidney disease | 8 (3·6) | 21 (5·8) | 8 (4·6) | 14 (5·0) | 51 (4·9) |
| **MALIGNANCY** | ·· | ·· | ·· | ·· | ·· |
| Solid tumour malignancy | ·· | ·· | ·· | ·· | ·· |
| *Localised* | 5 (2·3) | 19 (5·2) | 5 (2·9) | 4 (1·4) | 33 (3·2) |
| *Metastatic* | 2 (0·9) | 3 (0·8) | 0 (0·0) | 2 (0·7) | 7 (0·7) |
| Leukaemia | 1 (0·5) | 5 (1·4) | 2 (1·2) | 3 (1·1) | 11 (1·1) |
| Lymphoma | 4 (1·8) | 4 (1·1) | 2 (1·2) | 1 (0·4) | 11 (1·1) |
| **CHRONIC INFECTIOUS DISEASE** | ·· | ·· | ·· | ·· | ·· |
| HIV | 0 (0·0) | 1 (0·3) | 1 (0·6) | 3 (1·1) | 5 (0·5) |
| Chronic Viral Hepatitis (B or C) | 5 (2·3) | 1 (0·3) | 3 (1·7) | 4 (1·5) | 13 (1·3) |
| Mycobacterium TB (previously treated active or latent) | 5 (2·3) | 3 (0·8) | 2 (1·2) | 3 (1·1) | 13 (1·3) |

Data are n (%). WHO = World Health Organisation. Category 3-4 = no continuous supplemental oxygen needed, 5= continuous supplemental oxygen only, 6= Continuous or Bi-level Positive Airway Pressure ventilation or High Flow Nasal O_2_, 7-9 = Invasive Mechanical Ventilation or other organ support

**Table SR2. Occupation data at baseline, and change in employment by severity of acute illness and by cluster severity**

**a. Occupation data at before hospitalisation for COVID-19 stratified by severity of acute illness**

|  | **WHO – class 3-4** | **WHO – class 5** | **WHO – class 6** | **WHO – class 7-9** | **Total** |
| --- | --- | --- | --- | --- | --- |
| Working full-time | 110 (54·5) | 172 (52·6) | 90 (53·9) | 175 (68·9) | 547 (57·6) |
| Working part-time | 23 (11·4) | 31 (9·5) | 19 (11·4) | 21 (8·3) | 94 (9·9) |
| Full time carer (children or other) | 5 (2·5) | 6 (1·8) | 0 (0·0) | 0 (0·0) | 11 (1·2) |
| Unemployed | 9 (4·5) | 11 (3·4) | 2 (1·2) | 6 (2·4) | 28 (2·9) |
| Unable to work due to chronic illness | 3 (1·5) | 6 (1·8) | 4 (2·4) | 7 (2·8) | 20 (2·1) |
| Student | 3 (1·5) | 2 (0·6) | 2 (1·2) | 3 (1·2) | 10 (1·1) |
| Retired | 43 (21·3) | 95 (29·1) | 47 (28·1) | 39 (15·4) | 224 (23·6) |
| Medically retired | 5 (2·5) | 2 (0·6) | 3 (1·8) | 3 (1·2) | 13 (1·4) |
| Prefer not to say | 1 (0·5) | 2 (0·6) | 0 (0·0) | 0 (0·0) | 3 (0·3) |
| (Missing) | 24 | 51 | 18 | 34 | 127 |

Variables are presented as n and % of total n in each severity category. % are out of a total of 950. WHO = World Health Organisation. Category 3-4 = no continuous supplemental oxygen needed, 5= continuous supplemental oxygen only, 6= Continuous or Bi-level Positive Airway Pressure ventilation or High Flow Nasal O_2_, 7-9 = Invasive Mechanical Ventilation or other organ support

**Table SR2b. Change in occupation status COVID stratified by severity of acute illness.**

|  | **WHO – class 3-4** | **WHO – class 5** | **WHO – class 6** | **WHO – class 7-9** | **Total** |
| --- | --- | --- | --- | --- | --- |
| Working full-time or part-time before COVID-19 | 133 | 203 | 109 | 196 | 641 |
| No longer working after COVID-19 | 15 (11.3) | 24 (11.8) | 20 (18.3) | 54 (27.6) | 113 (17.8) |
| Occupation change due to health after COVID-19 | 19 (14.3) | 19 (9.4) | 18 (16.5) | 68 (34.7) | 124 (19.3) |

Variables are presented as n and % of total n in each severity category. Percentages are the proportion of those who reported working full-time or part-time before COVID-19. Participants were classified as no longer working post-hospitalisation for COVID-19 if they reported working full or part-time before COVID-19, subsequently answered “different from before” when asked “What is your main occupation/working status today?” and answered “Unable to work due to chronic illness/ /Medically retired”. Participants who reported working full or part-time before COVID-19 were classified as experiencing an occupation change due to health if they answered “different from before” when asked “What is your main occupation/working status today?” and then answered “Poor health/Sick leave” when asked “If different, why did your occupation/working status change?”.

**Table SR2c Occupation data before hospitalisation for COVID-19 stratified by cluster.**

|  | **Cluster 1** | **Cluster 2** | **Cluster 3** | **Cluster 4** | **Total** |
| --- | --- | --- | --- | --- | --- |
| Working full-time | 59 (52·2) | 94 (67·1) | 45 (42·9) | 211 (66·6) | 409 (60·6) |
| Working part-time | 11 (9·7) | 14 (10·0) | 17 (16·2) | 20 (6·3) | 62 (9·2) |
| Full time carer (children or other) | 4 (3·5) | 1 (0·7) | 0 (0·0) | 2 (0·6) | 7 (1·0) |
| Unemployed | 9 (8·0) | 2 (1·4) | 1 (1·0) | 4 (1·3) | 16 (2·4) |
| Unable to work due to chronic illness | 8 (7·1) | 3 (2·1) | 0 (0·0) | 2 (0·6) | 13 (1·9) |
| Student | 1 (0·9) | 1 (0·7) | 1 (1·0) | 3 (0·9) | 6 (0·9) |
| Retired | 16 (14·2) | 22 (15·7) | 38 (36·2) | 74 (23·3) | 150 (22·2) |
| Medically retired | 5 (4·4) | 3 (2·1) | 1 (1·0) | 0 (0·0) | 9 (1·3) |
| Prefer not to say | 0 (0·0) | 0 (0·0) | 2 (1·9) | 1 (0·3) | 3 (0·4) |
| (Missing) | 18 | 19 | 22 | 33 | 92 |

Variables are presented as n and % of total n in each severity category.

**Table SR2d Change in occupation status after hospitalisation for COVID-19 stratified by cluster.**

|  | **Cluster 1** | **Cluster 2** | **Cluster 3** | **Cluster 4** | **Total** |
| --- | --- | --- | --- | --- | --- |
| Working full-time or part-time before COVID-19 | 70 | 108 | 62 | 231 | 471 |
| No longer working after COVID-19 | 35 (50.0) | 12 (11.1) | 10 (16.1) | 23 (10.0) | 78 (16.6) |
| Occupation change due to health after COVID-19 | 42 (60.0) | 21 (19.4) | 10 (16.1) | 20 (8.7) | 93 (19.7) |

Variables are presented as n and % of total n in each severity category. See footnote to Table SR2b

**Table SR3.** **Patient reported outcomes, physiological and biochemical tests stratified by severity of acute illness.**

|  | **N (%)** | **WHO –**  **class 3-4** | **WHO –**  **class 5** | **WHO –**  **class 6** | **WHO –**  **class 7-9** | **Total** | **p** |
| --- | --- | --- | --- | --- | --- | --- | --- |
| Total N (%) | ·· | 226 (21·0) | 378 (35·1) | 185 (17·2) | 288 (26·7) | 1077 | ·· |
| **PROMS** | ·· | ·· | ·· | ·· | ·· | ·· | ·· |
| Any symptom at 3 months | 1077 (100·0) | 167 (73·9) | 264 (69·8) | 135 (73·0) | 231 (80·2) | 797 (74·0) | 0·026 |
| Symptom count | 1077 (100·0) | 9·5 (9·7) | 6·8 (7·5) | 8·2 (7·9) | 9·3 (8·1) | 8·3 (8·3) | <0·001 |
| GAD7 total score | 1031 (95·7) | 5·8 (6·2) | 4·4 (5·3) | 5·4 (5·7) | 5·5 (6·0) | 5·2 (5·8) | 0·017 |
| Anxiety (GAD7 >8) | 1031 (95·7) | 57 (26·8) | 72 (19·9) | 44 (25·3) | 80 (28·4) | 253 (24·5) | 0·069 |
| Missing |  | 13 | 16 | 11 | 6 | 46 |  |
| PHQ-9 total score | 1029 (95·5) | 7·6 (7·0) | 5·7 (6·0) | 6·4 (6·1) | 7·3 (6·7) | 6·7 (6·5) | 0·002 |
| Depression (PHQ-9 ≥ 10) | 1029 (95·5) | 64 (30·2) | 79 (21·9) | 49 (28·0) | 90 (32·0) | 282 (27·4) | 0·024 |
| Missing |  | 14 | 17 | 10 | 7 | 48 |  |
| PCL-5 Total Severity Score | 1030 (95·6) | 15·9 (18·6) | 12·5 (14·7) | 15·5 (17·1) | 18·6 (19·1) | 15·4 (17·3) | <0·001 |
| PTSD (PCL-5 ≥38) | 1030 (95·6) | 29 (13·6) | 31 (8·5) | 21 (12·0) | 45 (16·3) | 126 (12·2) | 0·025 |
| Missing |  | 12 | 13 | 10 | 12 | 47 |  |
| Dyspnoea-12 score | 1017 (94·4) | 7·2 (9·4) | 5·5 (7·7) | 6·5 (8·8) | 6·5 (8·8) | 6·3 (8·6) | 0·107 |
| FACIT fatigue subscale score | 1036 (96·2) | 18·5 (14·3) | 14·6 (12·1) | 16·4 (13·1) | 18·5 (13·4) | 16·8 (13·2) | <0·001 |
| BPI severity | 801 (74·4) | 12·7 (10·3) | 12·8 (10·6) | 11·6 (9·6) | 13·1 (10·3) | 12·7 (10·3) | 0·558 |
| BPI interference | 777 (72·1) | 19·8 (20·8) | 17·6 (18·2) | 15·1 (16·7) | 20·4 (19·6) | 18·4 (19·0) | 0·059 |
| **Body composition by BMI** kg/m^2^ | 908 (84·3) | ·· | ·· | ·· | ·· | ·· | ·· |
| Underweight (<18·5) | ·· | 2 (1·1) | 2 (0·6) | 1 (0·6) | 1 (0·4) | 6 (0·7) | 0.007 |
| Normal weight (18·5 to 24·9) | ·· | 43 (23·5) | 42 (13·0) | 16 (10·1) | 27 (11·1) | 128 (14·1) | ·· |
| Overweight (25 to 29·9) | ·· | 58 (31·7) | 122 (37·8) | 45 (28·3) | 85 (35·0) | 310 (34·1) | ·· |
| Obese (30 to 39·9) | ·· | 67 (36·6) | 131 (40·6) | 76 (47·8) | 101 (41·6) | 375 (41·3) | ·· |
| Severe obesity (40+) | ·· | 13 (7·1) | 26 (8·0) | 21 (13·2) | 29 (11·9) | 89 (9·8) | ·· |
| Missing |  | 43 | 55 | 26 | 45 | 169 |  |
| **Physical performance** | ·· | ·· | ·· | ·· | ·· | ·· | ·· |
| SPPB total score (0-12) | 970 (90·1) | 10·0 (2·3) | 9·9 (2·5) | 9·9 (2·5) | 9·7 (2·4) | 9·9 (2·4) | 0·698 |
| SPPB <=10 (mobility disability) | 970 (90·1) | 93 (46·7) | 153 (44·9) | 68 (40·5) | 134 (51·1) | 448 (46·2) | 0·168 |
| Missing |  |  |  |  |  |  |  |
| ISWT Distance (m) †† | 634 (58·9) | 466 (270) | 445 (273) | 425 (255) | 411 (236) | 436 (260) | 0·296 |
| ISWT % predicted | 634 (58·9) | 50·4 (37·8) | 50·1 (38·7) | 44·7 (32·4) | 39·4 (31·4) | 46·2 (35·8) | 0·010 |
| **Frailty and Cognition** | ·· | ·· | ·· | ·· | ·· | ·· | ·· |
| Rockwood clinical frailty score ≥5 (n - %) | 938 (87·1) | 9 (4·6) | 17 (5·0) | 11 (7·2) | 18 (7·2) | 55 (5·9) | 0·502 |
| 1 = Very Fit | 938 (87·1) | 28 (14·4) | 50 (14·7) | 28 (18·3) | 21 (8·4) | 127 (13·5) | 0·449 |
| 2 = Well | ·· | 68 (34·9) | 109 (32·0) | 43 (28·1) | 76 (30·5) | 296 (31·6) | ·· |
| 3 = Managing Well | ·· | 58 (29·7) | 116 (34·0) | 54 (35·3) | 92 (36·9) | 320 (34·1) | ·· |
| 4 = Vulnerable | ·· | 32 (16·4) | 49 (14·4) | 17 (11·1) | 42 (16·9) | 140 (14·9) | ·· |
| 5 = Mildly Frail | ·· | 5 (2·6) | 11 (3·2) | 7 (4·6) | 12 (4·8) | 35 (3·7) | ·· |
| 6 = Moderately Frail | ·· | 4 (2·1) | 5 (1·5) | 4 (2·6) | 6 (2·4) | 19 (2·0) | ·· |
| 7 = Severely Frail | ·· | 0 (0·0) | 1 (0·3) | 0 (0·0) | 0 (0·0) | 1 (0·1) | ·· |
| 8 = Very Severely Frail | ·· | 0 (0·0) | 0 (0·0) | 0 (0·0) | 0 (0·0) | 0 (0·0) | ·· |
| 9 = Terminally Ill | ·· | 0 (0·0) | 0 (0·0) | 0 (0·0) | 0 (0·0) | 0 (0·0) | ·· |
| (Missing) | ·· | 31 | 37 | 32 | 39 | 139 | ·· |
| MoCA score | 888 (82·5) | 25·9 (3·4) | 25·1 (4·4) | 25·9 (2·9) | 25·8 (3·9) | 25·6 (3·9) | 0·063 |
| MoCA <23 | 888 (82·5) | 25 (13·5) | 66 (21·0) | 19 (12·8) | 40 (16·7) | 150 (16·9) | 0·074 |
| MoCA Adjusted | 888 (82·5) | 26·2 (3·4) | 25·5 (4·3) | 26·2 (2·9) | 26·0 (3·8) | 25·9 (3·8) | 0·099 |
| MoCA Adjusted <23 | 888 (82·5) | 23 (12·4) | 57 (18·1) | 16 (10·8) | 33 (13·8) | 129 (14·5) | 0·130 |
| **Lung physiology** | ·· | ·· | ·· | ·· | ·· | ·· | ·· |
| FEV_1_ (L) | 574 (53·3) | 2·6 (0·7) | 2·7 (0·9) | 2·7 (0·8) | 2·7 (0·9) | 2·7 (0·8) | 0·623 |
| FEV_1_ % predicted | 484 (44·9) | 88·6 (19·6) | 89·0 (18·3) | 90·9 (31·5) | 85·0 (23·9) | 88·0 (22·9) | 0·247 |
| FEV_1_ % predicted <80%¶ | 484 (44·9) | 26 (28·6) | 43 (26·1) | 23 (28·4) | 58 (39·5) | 150 (31·0) | 0·063 |
| FVC (L) | 571 (53·0) | 3·3 (0·9) | 3·5 (1·1) | 3·4 (1·0) | 3·3 (1·1) | 3·4 (1·0) | 0·172 |
| FVC % predicted | 481 (44·7) | 88·1 (19·3) | 89·5 (16·4) | 92·0 (36·0) | 80·7 (19·8) | 87·0 (22·7) | 0·001 |
| FVC % predicted <80%¶ | 481 (44·7) | 30 (33·0) | 43 (26·4) | 25 (30·9) | 62 (42·5) | 160 (33·3) | 0·026 |
| FEV_1_/FVC | 571 (53·0) | 0·8 (0·2) | 0·8 (0·1) | 0·8 (0·1) | 0·8 (0·2) | 0·8 (0·2) | 0·007 |
| FEV_1_/FVC <0·7¶ | 571 (53·0) | 13 (12·3) | 27 (13·8) | 15 (16·0) | 6 (3·4) | 61 (10·7) | 0·002 |
| TLCO mmol/KPa/min | 194 (18·0) | 7·0 (1·8) | 7·1 (1·5) | 7·3 (2·0) | 7·0 (2·5) | 7·1 (2·0) | 0·952 |
| TLCO % predicted | 169 (15·7) | 97·5 (16·4) | 89·6 (18·9) | 97·1 (39·4) | 83·3 (32·8) | 89·8 (28·6) | 0·099 |
| TLCO % predicted <80%¶ | 169 (15·7) | 3 (15·8) | 19 (30·2) | 6 (19·4) | 30 (53·6) | 58 (34·3) | 0·001 |
| KCO mmol/Kpa/min/L | 202 (18·8) | 1·5 (0·3) | 1·4 (0·3) | 1·4 (0·2) | 1·4 (0·3) | 1·4 (0·3) | 0·153 |
| KCO % predicted | 174 (16·2) | 104·8 (15·6) | 97·1 (16·4) | 98·9 (14·6) | 96·1 (20·4) | 97·9 (17·6) | 0·289 |
| KCO % predicted <80%¶ | 174 (16·2) | 2 (10·5) | 7 (10·9) | 2 (6·2) | 5 (8·5) | 16 (9·2) | 0·887 |
| **Biochemical Tests** | ·· | ·· | ·· | ·· | ·· | ·· | ·· |
| BNP Result (ng/L) | 51 (4·7) | 49·6 (53·9) | 40·9 (63·0) | 43·6 (53·3) | 38·1 (39·4) | 42·2 (52·0) | 0·960 |
| Pro-NT-BNP (ng/L) | 572 (53·1) | 129 (297) | 173 (382) | 177 (460) | 305 (1832) | 201 (1023) | 0·473 |
| BNP/NT-Pro-BNP above threshold*¶ | 621 (57·7) | 8 (5·8) | 15 (7·2) | 8 (8·0) | 15 (8·5) | 46 (7·4) | 0·825 |
| HbA1C % (DCCT/NGSP) | 611 (56·7 | 5·9 (1·1) | 6·3 (1·3) | 6·1 (1·1) | 6·0 (1·2) | 6·1 (1·2) | 0·010 |
| HbA1C ≥6.0 ¶ | 611 (56·7) | 37 (27·2) | 90 (42·3) | 39 (41·1) | 47 (28·1) | 213 (34·9) | 0·004 |
| eGFR Result (ml/min/1·73m2) | 845 (78·5) | 89·0 (76·6) | 79·5 (44·4) | 81·2 (62·6) | 84·2 (83·1) | 83·0 (66·7) | 0·493 |
| eGFR < 60 ml/min/1·73 m^2^¶ | 845 (78·5) | 15 (8·5) | 42 (14·0) | 18 (13·0) | 38 (16·4) | 113 (13·4) | 0·138 |
| D-Dimer Result (mg/L) | 738 (68·5) | 285·2 (430·4) | 332·5 (381·6) | 344·6 (369·0) | 233·2 (168·8) | 298·0 (349·7) | 0·008 |
| D-dimer ≥500 ng/ml¶ | 738 (68·5) | 15 (9·7) | 45 (17·2) | 22 (17·6) | 15 (7·6) | 97 (13·1) | 0·005 |
| **Systemic Inflammation** | ·· | ·· | ·· | ·· | ·· | ·· | ·· |
| CRP (>10 mg/L)¶ | 804 (74·7) | 18 (10·7) | 24 (8·4) | 13 (10·0) | 35 (16·1) | 90 (11·2) | 0·052 |

Missing not included in %, Number (%) unless †median [IQR], †† mean [SD], ¶ = % of category with positive response *Threshold - BNP ≥100ng/L or NT-BNP ≥400ng/L, column proportions, P values for Chi-squared test for differing proportions across WHO categories are presented, P values for Kruskal-Wallis tests for variables summarised as median (IQR) are presented and P values for ANOVA F-test for variables summarised as mean [SD] are presented. DCCT/NGSP - Diabetes Control and Complications Trial / National Glycohemoglobin Standardization Programme, PROM = Patient reported outcome measures, GAD7 = General Anxiety Disorder 7 Questionnaire, PHQ-9 = Patient Health Questionnaire-9, PCL-5 = Post Traumatic Stress Disorder Checklist, Dyspnoea-12 Questionnaire, FACIT Fatigue Scale (Facit), BPI =Brief Pain Inventory, SPPB = Short Physical Performance Battery, ISWT = Incremental Shuttle Walking Test, CFS = Clinical Frailty Scale, MoCA = Montreal Cognitive Assessment, FEV1 = Forced Expiratory Volume in 1 second, FVC = Forced Vital Capacity, TLCO = Transfer Capacity of the Lung for Carbon Monoxide, KCO = carbon monoxide transfer coefficient, BNP = Brain Natriuretic Peptide or NT-BNP N-Terminal Brain Natriuretic Peptide, HbA1C = glycosylated haemoglobin, eGFR = estimated Glomerular Filtration Rate, CRP = C-Reactive Protein. WHO = World Health Organisation. Category 3-4 = no continuous supplemental oxygen needed, 5= continuous supplemental oxygen only, 6= Continuous or Bi-level Positive Airway Pressure ventilation or High Flow Nasal O_2_, 7-9 = Invasive Mechanical Ventilation or other organ support

**Table SR4**. **Comparison between imputed and non-imputed logistic regression of predictors of failure to recover (multi-variable and multi-level).**

| **Dependent: ‘Fully Recovered’** | **No**  **n (%)** | **Yes**  **n (%)** | **OR**  **(univariable)** | **OR**  **(multivariable)** | **OR (multivariable imputation)** | **OR**  **(multilevel imputation)** |
| --- | --- | --- | --- | --- | --- | --- |
| **Age (y)** | ·· | ·· | ·· | ·· | ·· | ·· |
| 50-59 | 183 (78.2) | 51 (21.8) | - | - | - | - |
| <30 | 16 (61.5) | 10 (38.5) | 2.24 (0.93-5.18, p=0.062) | 1.61 (0.50-4.83, p=0.406) | 2.27 (0.83-6.21, p=0.111) | 2.28 (0.83-6.29, p=0.109) |
| 30-39 | 43 (69.4) | 19 (30.6) | 1.59 (0.84-2.93, p=0.147) | 1.30 (0.59-2.79, p=0.511) | 1.42 (0.72-2.80, p=0.314) | 1.48 (0.73-2.97, p=0.272) |
| 40-49 | 88 (75.2) | 29 (24.8) | 1.18 (0.70-1.98, p=0.529) | 0.66 (0.32-1.32, p=0.248) | 1.08 (0.63-1.83, p=0.789) | 1.10 (0.64-1.88, p=0.735) |
| 60-69 | 192 (72.7) | 72 (27.3) | 1.35 (0.89-2.04, p=0.158) | 1.19 (0.70-2.03, p=0.525) | 1.33 (0.85-2.08, p=0.215) | 1.34 (0.85-2.12, p=0.201) |
| 70-79 | 71 (59.7) | 48 (40.3) | 2.43 (1.50-3.93, p<0.001) | 2.78 (1.49-5.22, p=0.001) | 1.96 (1.09-3.53, p=0.026) | 2.07 (1.13-3.80, p=0.020) |
| 80+ | 16 (50.0) | 16 (50.0) | 3.59 (1.67-7.72, p=0.001) | 4.04 (1.57-10.60, p=0.004) | 2.86 (1.31-6.23, p=0.008) | 3.20 (1.44-7.14, p=0.005) |
| **Sex at birth** | ·· | ·· | ·· | ·· | ·· | ·· |
| Male | 372 (67.1) | 182 (32.9) | - | - | - | - |
| Female | 246 (78.3) | 68 (21.7) | 0.56 (0.41-0.78, p=0.001) | 0.50 (0.33-0.77, p=0.002) | 0.61 (0.42-0.89, p=0.012) | 0.62 (0.43-0.92, p=0.017) |
| **Ethnicity** | ·· | ·· | ·· | ·· | ·· | ·· |
| White | 442 (75.6) | 143 (24.4) | - | - | - | - |
| South Asian | 83 (62.9) | 49 (37.1) | 1.82 (1.22-2.72, p=0.003) | 1.68 (0.94-2.98, p=0.076) | 1.58 (1.00-2.52, p=0.052) | 1.54 (0.94-2.53, p=0.085) |
| Black | 46 (66.7) | 23 (33.3) | 1.55 (0.89-2.61, p=0.111) | 2.27 (1.12-4.54, p=0.021) | 1.82 (1.01-3.28, p=0.046) | 1.83 (0.99-3.39, p=0.053) |
| Mixed | 12 (60.0) | 8 (40.0) | 2.06 (0.79-5.08, p=0.121) | 1.56 (0.48-4.73, p=0.439) | 1.59 (0.62-4.06, p=0.333) | 1.69 (0.65-4.38, p=0.279) |
| Other | 20 (52.6) | 18 (47.4) | 2.78 (1.42-5.41, p=0.003) | 2.82 (1.05-7.33, p=0.035) | 2.69 (1.31-5.53, p=0.007) | 2.71 (1.32-5.59, p=0.007) |
| **IMD** | ·· | ·· | ·· | ·· | ·· | ·· |
| 1 | 128 (74.0) | 45 (26.0) | - | - | - | - |
| 2 | 129 (68.3) | 60 (31.7) | 1.32 (0.84-2.10, p=0.230) | 1.42 (0.78-2.58, p=0.252) | 1.17 (0.72-1.91, p=0.521) | 1.17 (0.72-1.90, p=0.522) |
| 3 | 129 (75.4) | 42 (24.6) | 0.93 (0.57-1.51, p=0.757) | 0.94 (0.50-1.76, p=0.845) | 0.88 (0.52-1.50, p=0.637) | 0.85 (0.50-1.45, p=0.550) |
| 4 | 109 (69.0) | 49 (31.0) | 1.28 (0.79-2.07, p=0.314) | 1.31 (0.69-2.46, p=0.408) | 1.24 (0.75-2.04, p=0.397) | 1.22 (0.73-2.02, p=0.450) |
| 5 | 112 (70.9) | 46 (29.1) | 1.17 (0.72-1.90, p=0.528) | 1.46 (0.78-2.75, p=0.234) | 1.21 (0.73-1.99, p=0.461) | 1.20 (0.73-1.99, p=0.472) |
| **No· of comorbidities**¶ | ·· | ·· | ·· | ·· | ·· | ·· |
| 0 | 150 (65.2) | 80 (34.8) | - | - | - | - |
| 1 | 123 (67.6) | 59 (32.4) | 0.90 (0.59-1.36, p=0.614) | 0.85 (0.49-1.47, p=0.557) | 0.99 (0.64-1.52, p=0.964) | 0.97 (0.62-1.51, p=0.892) |
| 2+ | 345 (75.7) | 111 (24.3) | 0.60 (0.43-0.85, p=0.004) | 0.44 (0.27-0.73, p=0.001) | 0.65 (0.45-0.95, p=0.026) | 0.65 (0.44-0.95, p=0.026) |
| **BMI**¶ |  |  |  |  |  |  |
| BMI <30 kg/m^2^ | 242 (64.9) | 131 (35.1) | - | - | - | - |
| BMI ≥30 kg/m^2^ | 296 (79.1) | 78 (20.9) | 0.49 (0.35-0.67, p<0.001) | 0.65 (0.43-0.97, p=0.035) | 0.74 (0.54-1.03, p=0.073) | 0.74 (0.53-1.04, p=0.082) |
| **WHO Class**¶ | ·· | ·· | ·· | ·· | ·· | ·· |
| 3-4 | 120 (68.2) | 56 (31.8) | - | - | - | - |
| 5 | 185 (63.4) | 107 (36.6) | 1.24 (0.84-1.85, p=0.289) | 1.04 (0.59-1.85, p=0.895) | 1.15 (0.74-1.79, p=0.540) | 1.11 (0.71-1.76, p=0.640) |
| 6 | 108 (72.5) | 41 (27.5) | 0.81 (0.50-1.31, p=0.399) | 0.58 (0.29-1.15, p=0.120) | 0.82 (0.45-1.48, p=0.500) | 0.79 (0.43-1.45, p=0.436) |
| 7-9 | 205 (81.7) | 46 (18.3) | 0.48 (0.31-0.75, p=0.001) | 0.27 (0.13-0.57, p=0.001) | 0.53 (0.30-0.94, p=0.029) | 0.54 (0.30-0.96, p=0.034) |
| **Steroids**¶ | ·· | ·· | ·· | ·· | ·· | ·· |
| No | 376 (69.1) | 168 (30.9) | - | - | - | - |
| Yes | 181 (73.6) | 65 (26.4) | 0.80 (0.57-1.12, p=0.204) | 1.22 (0.77-1.94, p=0.394) | 1.00 (0.69-1.44, p=0.994) | 1.02 (0.70-1.49, p=0.909) |
| **Antibiotics**¶ | ·· | ·· | ·· | ·· | ·· | ·· |
| No | 114 (72.6) | 43 (27.4) | - | - | - | - |
| Yes | 481 (71.0) | 196 (29.0) | 1.08 (0.74-1.61, p=0.696) | 1.49 (0.87-2.60, p=0.149) | 1.20 (0.77-1.87, p=0.416) | 1.21 (0.77-1.90, p=0.410) |
| **Anticoagulatio**n¶ | ·· | ·· | ·· | ·· | ·· | ·· |
| No | 353 (67.4) | 171 (32.6) | - | - | - | - |
| Yes | 207 (75.3) | 68 (24.7) | 0.68 (0.49-0.94, p=0.021) | 0.64 (0.40-1.02, p=0.063) | 0.80 (0.55-1.17, p=0.251) | 0.78 (0.53-1.15, p=0.202) |

OR =Odds Ratio, Data are n (%) unless otherwise stated. ¶ = % of category with positive response BMI = Body Mass Index, IMD = Indices of Multiple Deprivation, WHO = World Health Organisation. Category 3-4 = no continuous supplemental oxygen needed, 5= continuous supplemental oxygen only, 6= Continuous or Bi-level Positive Airway Pressure ventilation or High Flow Nasal O_2_, 7-9 = Invasive Mechanical Ventilation or other organ support

**Table SR5.** **Ongoing symptoms recorded at follow-up for the cohort stratified between those with and without pre-existing co-morbidities.**

|  | **Persisting Symptom** | **No comorbidity** | **1+ comorbidity** | **Total** |
| --- | --- | --- | --- | --- |
| Total N (%) | ·· | 315 (36·8) | 540 (63·2) | 855 |
| Any symptom | Yes | 201 (86.6) | 431 (96.0) | 632 (92.8) |
| Symptom count † |  | 7.0 (2.0 to 13.0) | 10.0 (5.0 to 17.0) | 9.0 (4.0 to 16.0) |
| Aching in your muscles (pain) | Yes | 105 (45·3) | 280 (63·5) | 385 (57·2) |
|  | (Missing) | 83 | 99 | 182 |
| Physical slowing down | Yes | 95 (41·1) | 242 (54·4) | 337 (49·9) |
|  | (Missing) | 84 | 95 | 179 |
| Slowing down in your thinking | Yes | 82 (36·0) | 201 (45·7) | 283 (42·4) |
|  | (Missing) | 87 | 100 | 187 |
| Joint pain or swelling | Yes | 79 (35·3) | 236 (54·3) | 315 (47·8) |
|  | (Missing) | 91 | 105 | 196 |
| Limb weakness | Yes | 79 (34·3) | 231 (52·6) | 310 (46·3) |
|  | (Missing) | 85 | 101 | 186 |
| Difficulty with concentration | Yes | 77 (33·9) | 191 (43·5) | 268 (40·2) |
|  | (Missing) | 88 | 101 | 189 |
| Short term memory loss | Yes | 77 (33·8) | 202 (46·2) | 279 (42·0) |
|  | (Missing) | 87 | 103 | 190 |
| Headache | Yes | 73 (31·7) | 151 (34·2) | 224 (33·4) |
|  | (Missing) | 85 | 99 | 184 |
| Tingling feeling/pins and needles | Yes | 60 (26·9) | 186 (42·5) | 246 (37·2) |
|  | (Missing) | 92 | 102 | 194 |
| Confusion/fuzzy head | Yes | 57 (24·9) | 146 (33·0) | 203 (30·2) |
|  | (Missing) | 86 | 97 | 183 |
| Dizziness or lightheaded | Yes | 55 (24·7) | 163 (37·6) | 218 (33·2) |
|  | (Missing) | 92 | 106 | 198 |
| Chest tightness | Yes | 55 (23·7) | 126 (28·7) | 181 (27·0) |
|  | (Missing) | 83 | 101 | 184 |
| Problems with balance | Yes | 52 (23·3) | 183 (41·8) | 235 (35·6) |
|  | (Missing) | 92 | 102 | 194 |
| Altered personality/ behaviour § | Yes | 47 (20·3) | 93 (21·0) | 140 (20·8) |
|  | (Missing) | 84 | 97 | 181 |
| Chest pain | Yes | 42 (18·2) | 105 (23·8) | 147 (21·9) |
|  | (Missing) | 84 | 99 | 183 |
| Palpitations | Yes | 37 (17·0) | 95 (21·8) | 132 (20·2) |
|  | (Missing) | 97 | 105 | 202 |
| Leg/ankle swelling | Yes | 39 (16·9) | 151 (34·2) | 190 (28·3) |
|  | (Missing) | 84 | 99 | 183 |
| Difficulty with communication | Yes | 37 (16·1) | 75 (17·0) | 112 (16·7) |
|  | (Missing) | 85 | 98 | 183 |
| Skin rash | Yes | 31 (14·0) | 73 (17·1) | 104 (16·0) |
|  | (Missing) | 93 | 112 | 205 |
| Diarrhoea | Yes | 31 (13·6) | 83 (18·7) | 114 (17·0) |
|  | (Missing) | 87 | 97 | 184 |
| Problems seeing | Yes | 28 (12·6) | 73 (16·7) | 101 (15·3) |
|  | (Missing) | 93 | 104 | 197 |
| Pain on breathing | Yes | 26 (11·7) | 68 (15·6) | 94 (14·3) |
|  | (Missing) | 92 | 105 | 197 |
| Weight loss | Yes | 24 (10·8) | 46 (10·6) | 70 (10·7) |
|  | (Missing) | 92 | 106 | 198 |
| Tremor/shakiness | Yes | 23 (10·3) | 69 (15·8) | 92 (13·9) |
|  | (Missing) | 92 | 103 | 195 |
| Constipation | Yes | 21 (9·3) | 109 (24·8) | 130 (19·5) |
|  | (Missing) | 88 | 101 | 189 |
| Erectile Dysfunction | Yes | 20 (9·2) | 75 (17·9) | 95 (14·9) |
|  | N/A | 72 (33·2) | 170 (40·6) | 242 (38·1) |
|  | (Missing) | 98 | 121 | 219 |
| Loss of sense of smell | Yes | 21 (9·2) | 46 (10·4) | 67 (10·0) |
|  | (Missing) | 86 | 97 | 183 |
| Can’t fully move or control movement | Yes | 20 (8·9) | 54 (12·4) | 74 (11·2) |
|  | (Missing) | 91 | 106 | 197 |
| Abdominal pain | Yes | 20 (8·8) | 95 (21·4) | 115 (17·1) |
|  | (Missing) | 87 | 97 | 184 |
| Stomach pain | Yes | 19 (8·6) | 82 (19·0) | 101 (15·5) |
|  | (Missing) | 94 | 108 | 202 |
| Loss of control of passing urine | Yes | 19 (8·3) | 52 (11·8) | 71 (10·6) |
|  | (Missing) | 85 | 98 | 183 |
| Loss of appetite | Yes | 19 (8·2) | 68 (15·3) | 87 (12·9) |
|  | (Missing) | 84 | 97 | 181 |
| Loss of taste | Yes | 18 (7·8) | 52 (11·7) | 70 (10·4) |
|  | (Missing) | 84 | 97 | 181 |
| Nausea/vomiting | Yes | 17 (7·5) | 51 (11·7) | 68 (10·2) |
|  | (Missing) | 87 | 103 | 190 |
| Bleeding | Yes | 13 (6·0) | 22 (5·4) | 35 (5·6) |
|  | (Missing) | 98 | 129 | 227 |
| Can’t move and/or feel one side of your body or face | Yes | 9 (4·0) | 30 (6·9) | 39 (5·9) |
|  | (Missing) | 90 | 104 | 194 |
| Loss of control of opening bowels | Yes | 7 (3·1) | 30 (6·8) | 37 (5·5) |
|  | (Missing) | 86 | 100 | 186 |
| Lumpy lesions on toes | Yes | 5 (2·3) | 13 (3·2) | 18 (2·9) |
|  | (Missing) | 100 | 134 | 234 |
| Fainting / blackouts | Yes | 4 (1·8) | 11 (2·5) | 15 (2·3) |
|  | (Missing) | 93 | 108 | 201 |
| Seizures | Yes | 2 (0·9) | 5 (1·2) | 7 (1·1) |
|  | (Missing) | 93 | 106 | 199 |

Data are n (%).

**Table SR6. Proportion unchanged, worse or better in terms of a) Health-related quality of life (EQ5D-5L) b) Disability (WG-SS) and c) Symptoms at follow-up compared to prior to hospitalisation stratified by severity of acute illness.**

a) **EQ5D-5L**

|  | **WHO – class 3-4** | **WHO – class 5** | **WHO – class 6** | **WHO – class 7-9** | **Total** |
| --- | --- | --- | --- | --- | --- |
| **Do you feel fully recovered from COVID-19? (n - %)***** | ·· | ·· | ·· | ·· | ·· |
| Yes | 51 (30·9) | 102 (36·3) | 41 (28·5) | 45 (18·8) | 239 (28·8) |
| No | 75 (45·5) | 126 (44·8) | 65 (45·1) | 163 (67·9) | 429 (51·7) |
| Not sure | 39 (23·6) | 53 (18·9) | 38 (26·4) | 32 (13·3) | 162 (19·5) |
| Missing | 61 | 97 | 41 | 48 | 247 |
| **How good or bad is your health overall (EQ5D-5L VAS 0-100)? (Mean -SD)** | ·· | ·· | ·· | ·· | ·· |
| Pre-COVID | 78.7 (18.9) | 80.0 (17.4) | 81.6 (15.7) | 84.1 (13.7) | 81.1 (16.6) |
| Post-COVID | 70.5 (20.7) | 73.8 (18.7) | 70.9 (21.4) | 69.6 (18.9) | 71.5 (19.7) |
| Change since hospitalisation*** | -7.5 (21.6) | -7.9 (17.4) | -9.6 (17.6) | -14.7 (18.9) | -9.9 (19.0) |
| **EQ5D-5L Utility index (Mean -SD)***** | ·· | ·· | ·· | ·· | ·· |
| Pre-COVID | 0·82 (0·24) | 0·84 (0·22) | 0·82 (0·23) | 0·87 (0·21) | 0·84 (0·23) |
| Post-COVID | 0·72 (0·27) | 0·76 (0·24) | 0·69 (0·29) | 0·67 (0·25) | 0·71 (0·26) |
| Change | -0·09 (0·26) | -0·09 (0·20) | -0·11 (0·23) | -0·21 (0·24) | -0·13 (0·24) |
| **Mobility (n - %)***** | ·· | ·· | ·· | ·· | ·· |
| No change | 103 (67·8) | 163 (66·3) | 64 (58·2) | 95 (49·7) | 425 (60·8) |
| Improvement | 14 (9·2) | 20 (8·1) | 11 (10·0) | 6 (3·1) | 51 (7·3) |
| Worse | 35 (23·0) | 63 (25·6) | 35 (31·8) | 90 (47·1) | 223 (31·9) |
| (Missing) | 74 | 132 | 75 | 97 | 378 |
| **Self-Care (n - %)**** | ·· | ·· | ·· | ·· | ·· |
| No change | 94 (62·7) | 156 (63·2) | 57 (51·8) | 85 (44·5) | 392 (56·2) |
| Improvement | 4 (2·7) | 3 (1·2) | 2 (1·8) | 3 (1·6) | 12 (1·7) |
| Worse | 52 (34·7) | 88 (35·6) | 51 (46·4) | 103 (53·9) | 294 (42·1) |
| (Missing) | 76 | 131 | 75 | 97 | 379 |
| **Usual Activities (n - %)***** | ·· | ·· | ·· | ·· | ·· |
| No change | 89 (59·3) | 156 (63·2) | 61 (55·5) | 86 (45·0) | 392 (56·2) |
| Improvement | 16 (10·7) | 14 (5·7) | 9 (8·2) | 7 (3·7) | 46 (6·6) |
| Worse | 45 (30·0) | 77 (31·2) | 40 (36·4) | 98 (51·3) | 260 (37·2) |
| (Missing) | 76 | 131 | 75 | 97 | 379 |
| **Pain/Discomfort (n - %)***** | ·· | ·· | ·· | ·· | ·· |
| No change | 83 (55·3) | 143 (58·1) | 56 (50·9) | 79 (41·4) | 361 (51·8) |
| Improvement | 36 (24·0) | 53 (21·5) | 29 (26·4) | 33 (17·3) | 151 (21·7) |
| Worse | 31 (20·7) | 50 (20·3) | 25 (22·7) | 79 (41·4) | 185 (26·5) |
| (Missing) | 76 | 132 | 75 | 97 | 380 |
| **Anxiety/Depression (n - %)**** | ·· | ·· | ·· | ·· | ·· |
| No change | 74 (49·7) | 142 (57·3) | 45 (40·9) | 81 (42·4) | 342 (49·0) |
| Improvement | 27 (18·1) | 23 (9·3) | 18 (16·4) | 18 (9·4) | 86 (12·3) |
| Worse | 48 (32·2) | 83 (33·5) | 47 (42·7) | 92 (48·2) | 270 (38·7) |
| (Missing) | 77 | 130 | 75 | 97 | 379 |

Missing not included in %, *p<0·05, **p<0·01, ***p<0·0001, column proportions, EQ5D-5L VAS = Euroqol five level visual analogue scale 0-100, WHO = World Health Organisation. Category 3-4 = no continuous supplemental oxygen needed, 5= continuous supplemental oxygen only, 6= Continuous or Bi-level Positive Airway Pressure ventilation or High Flow Nasal O_2_, 7-9 = Invasive Mechanical Ventilation or other organ support, WGSS- Washington Group Short Set on Functioning

b) **Washington Short Set Function score**

|  | **WHO –class 3-4** | **WHO –class 5** | **WHO –class 6** | **WHO –class 7-9** | **Total** |
| --- | --- | --- | --- | --- | --- |
| **Q1 seeing N (%)** | ·· | ·· | ·· | ·· | ·· |
| No change | 130 (81·8) | 236 (87·1) | 114 (83·2) | 194 (83·3) | 674 (84·2) |
| Improvement | 2 (1·3) | 5 (1·8) | 4 (2·9) | 3 (1·3) | 14 (1·8) |
| Worse | 27 (17·0) | 30 (11·1) | 19 (13·9) | 36 (15·5) | 112 (14·0) |
| Missing N | 67 | 107 | 48 | 55 | 277 |
| **Q2 hearing N (%)** | ·· | ·· | ·· | ·· | ·· |
| No change | 143 (90·5) | 251 (94·4) | 131 (93·6) | 198 (86·8) | 723 (91·3) |
| Improvement | 3 (1·9) | 3 (1·1) | 1 (0·7) | 4 (1·8) | 11 (1·4) |
| Worse | 12 (7·6) | 12 (4·5) | 8 (5·7) | 26 (11·4) | 58 (7·3) |
| Missing N | 68 | 112 | 45 | 60 | 285 |
| **Q 3 walking N (%)***** | ·· | ·· | ·· | ·· | ·· |
| No change | 96 (61·1) | 182 (67·7) | 88 (64·2) | 104 (44·8) | 470 (59·1) |
| Improvement | 8 (5·1) | 16 (5·9) | 7 (5·1) | 20 (8·6) | 51 (6·4) |
| Worse | 53 (33·8) | 71 (26·4) | 42 (30·7) | 108 (46·6) | 274 (34·5) |
| Missing N | 69 | 109 | 48 | 56 | 282 |
| **Q4 remembering N (%)**** | ·· | ·· | ·· | ·· | ·· |
| No change | 96 (60·4) | 188 (69·9) | 87 (63·0) | 119 (51·3) | 490 (61·4) |
| Improvement | 6 (3·8) | 17 (6·3) | 7 (5·1) | 11 (4·7) | 41 (5·1) |
| Worse | 57 (35·8) | 64 (23·8) | 44 (31·9) | 102 (44·0) | 267 (33·5) |
| Missing N | 67 | 109 | 47 | 56 | 279 |
| **Q5 self-care N (%)***** | ·· | ·· | ·· | ·· | ·· |
| No change | 142 (90·4) | 254 (94·4) | 125 (89·9) | 172 (73·8) | 693 (86·8) |
| Improvement | 1 (0·6) | 1 (0·4) | 1 (0·7) | 10 (4·3) | 13 (1·6) |
| Worse | 14 (8·9) | 14 (5·2) | 13 (9·4) | 51 (21·9) | 92 (11·5) |
| Missing N | 69 | 109 | 46 | 55 | 279 |
| **Q6 communication N (%)** | ·· | ·· | ·· | ·· | ·· |
| No change | 143 (91·1) | 249 (93·3) | 124 (90·5) | 195 (85·5) | 711 (90·1) |
| Improvement | 1 (0·6) | 4 (1·5) | 2 (1·5) | 3 (1·3) | 10 (1·3) |
| Worse | 13 (8·3) | 14 (5·2) | 11 (8·0) | 30 (13·2) | 68 (8·6) |
| Missing N | 69 | 111 | 48 | 60 | 288 |
| **Any new Disability?**** | 41 (25·5) | 35 (12·9) | 25 (17·9) | 57 (24·4) | 158 (19·6) |

Missing not included in %, column proportions,*p<0·05, **p<0·01, ***p<0·0001, WHO = World Health Organisation. Category 3-4 = no continuous supplemental oxygen needed, 5= continuous supplemental oxygen only, 6= Continuous or Bi-level Positive Airway Pressure ventilation or High Flow Nasal O_2_, 7-9 = Invasive Mechanical Ventilation or other organ support

c) **Symptoms**

|  | **WHO –**  **class 3-4** | **WHO –**  **class 5** | **WHO –**  **class 6** | **WHO –**  **class 7-9** | **Total** |
| --- | --- | --- | --- | --- | --- |
| **Breathlessness N (%)***** | ·· | ·· | ·· | ·· | ·· |
| No change | 62 (39·7) | 115 (44·6) | 51 (38·3) | 71 (32·3) | 299 (39·0) |
| Improvement | 17 (10·9) | 37 (14·3) | 26 (19·5) | 19 (8·6) | 99 (12·9) |
| Worse | 77 (49·4) | 106 (41·1) | 56 (42·1) | 130 (59·1) | 369 (48·1) |
| *Missing N* | *70* | *120* | *52* | *68* | *310* |
| **Fatigue N (%)**** | ·· | ·· | ·· | ·· | ·· |
| No change | 46 (29·7) | 97 (37·6) | 43 (32·3) | 50 (22·9) | 236 (30·9) |
| Improvement | 19 (12·3) | 40 (15·5) | 17 (12·8) | 23 (10·6) | 99 (13·0) |
| Worse | 90 (58·1) | 121 (46·9) | 73 (54·9) | 145 (66·5) | 429 (56·2) |
| *Missing N* | *71* | *120* | *52* | *70* | *313* |
| **Cough N (%)** | ·· | ·· | ·· | ·· | ·· |
| No change | 94 (61·0) | 170 (66·1) | 76 (57·1) | 128 (59·0) | 468 (61·5) |
| Improvement | 14 (9·1) | 35 (13·6) | 18 (13·5) | 25 (11·5) | 92 (12·1) |
| Worse | 46 (29·9) | 52 (20·2) | 39 (29·3) | 64 (29·5) | 201 (26·4) |
| *Missing N* | *72* | *121* | *52* | *71* | *316* |
| **Pain N (%)***** | ·· | ·· | ·· | ·· | ·· |
| No change | 76 (50·7) | 152 (60·1) | 76 (56·7) | 81 (37·9) | 385 (51·3) |
| Improvement | 16 (10·7) | 29 (11·5) | 12 (9·0) | 18 (8·4) | 75 (10·0) |
| Worse | 58 (38·7) | 72 (28·5) | 46 (34·3) | 115 (53·7) | 291 (38·7) |
| *Missing N* | *76* | *125* | *51* | *74* | *326* |
| **Sleep N (%)**** | ·· | ·· | ·· | ·· | ·· |
| No change | 61 (40·1) | 116 (45·3) | 60 (44·8) | 70 (32·0) | 307 (40·3) |
| Improvement | 25 (16·4) | 52 (20·3) | 22 (16·4) | 37 (16·9) | 136 (17·9) |
| Worse | 66 (43·4) | 88 (34·4) | 52 (38·8) | 112 (51·1) | 318 (41·8) |
| *Missing N* | *74* | *122* | *51* | *69* | *316* |

Missing not included in %, column proportions. *p<0·05, **p<0·01, ***p<0·0001, WHO = World Health Organisation. Category 3-4 = no continuous supplemental oxygen needed, 5= continuous supplemental oxygen only, 6= Continuous or Bi-level Positive Airway Pressure ventilation or High Flow Nasal O_2_, 7-9 = Invasive Mechanical Ventilation or other organ support

**Table SR7. Cluster medoids and characteristics**

|  | **Cluster 1** | **Cluster 2** | **Cluster 3** | **Cluster 4** |
| --- | --- | --- | --- | --- |
| Size | 131 | 159 | 127 | 350 |
| **Cluster medoids (z-scores)** | ·· | ·· | ·· | ·· |
| Anxiety (GAD7) | 1·5921223 | 0·5270043 | -0·3605941 | -0·7156334 |
| Depression (PHQ7) | 1·6495548 | -0·066511 | -0·222517 | -0·846541 |
| PTSD (PCL5) | 1·1880387 | 0·5344896 | -0·3567137 | -0·713195 |
| Breathlessness (Dyspnoea 12) | 1·2262824 | 0·3746578 | -0·1119849 | -0·7202882 |
| Function (FACIT) | 1·3974373 | 0·7784612 | -0·4594909 | -0·8463509 |
| Physical performance (SPPB) | 0·4405809 | 0·0101025 | 0·0101025 | -0·4203759 |
| Cognition (MoCA) | -0·0714386 | -0·6086291 | 1·2715376 | -0·3400339 |
| **Cluster characteristics** | ·· | ·· | ·· | ·· |
| Maximal dissimilarity | 5·608549 | 4·146695 | 6·032896 | 3·159184 |
| Average dissimilarity | 2·539763 | 1·831245 | 1·864718 | 1·10634 |
| Isolation | 2·272929 | 1·680496 | 3·037516 | 1·590624 |

GAD7 = General Anxiety Disorder 7 Questionnaire, PHQ-9 = Patient Health Questionnaire-9, PCL-5 = Post Traumatic Stress Disorder Checklist, Dyspnoea-12 Questionnaire, FACIT Fatigue Scale (Facit), BPI =Brief Pain Inventory, SPPB = Short Physical Performance Battery, MoCA = Montreal Cognitive Assessment

**Figure SR1. Consort flow diagram for participants**

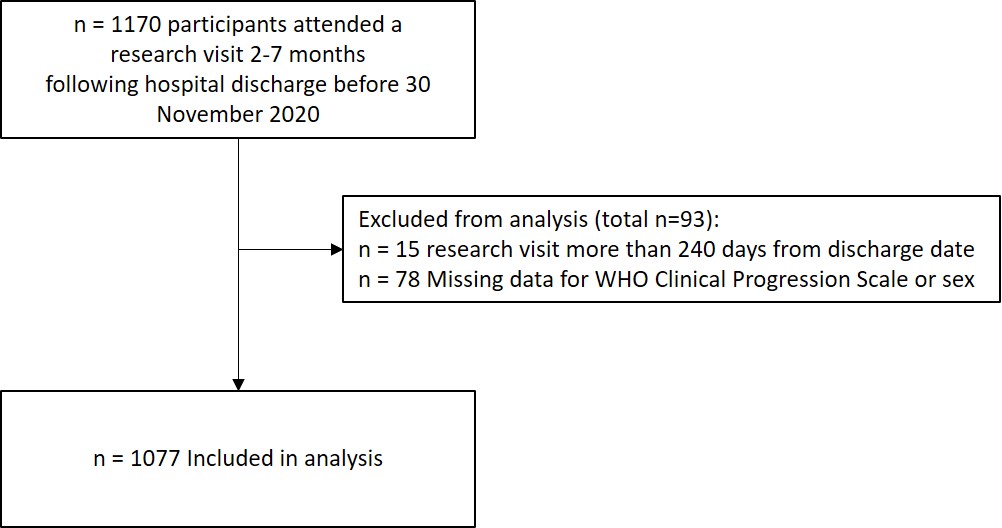

**Figure SR2. Histogram of number of symptoms reported at five months after discharge in survivors of a hospital admission due to COVID-19**

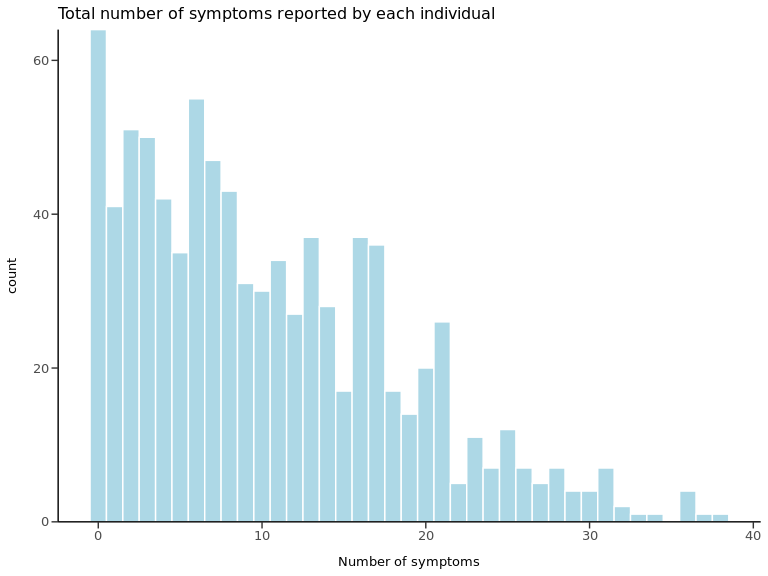

No. of symptoms reported

No. of participants

**Supplementary Figure SR3. Clusters of mental, cognitive and physical health impairments**

a) Scatter plots for anxiety versus other symptoms, cognition and physical function

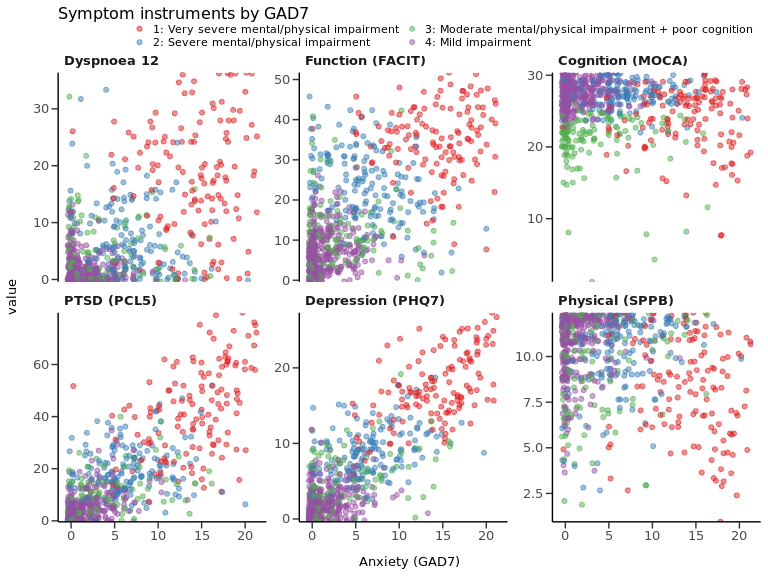

b) Scatter plots for breathlessness versus other symptoms, cognition and physical function

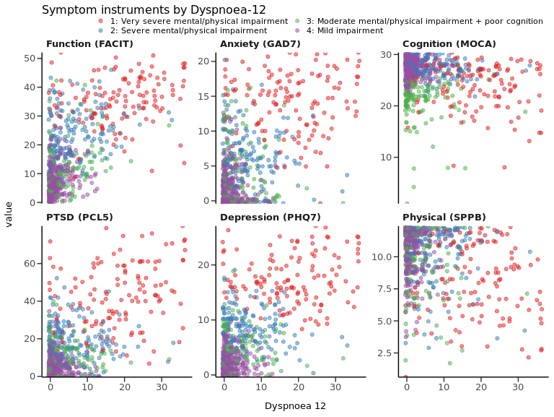

c) Scatter plots for physical function versus other symptoms and cognition

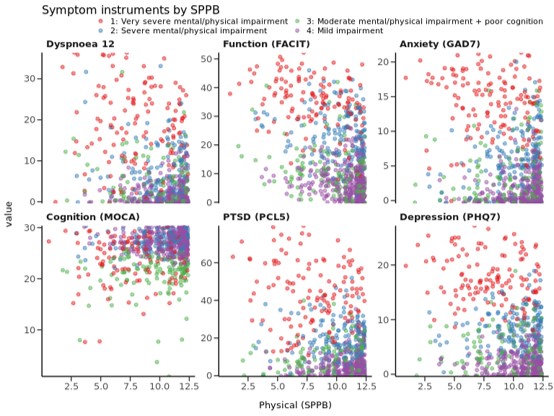

**Figure SR4 Correlation of C-Reactive Protein Level and Body Mass Index by clusters**

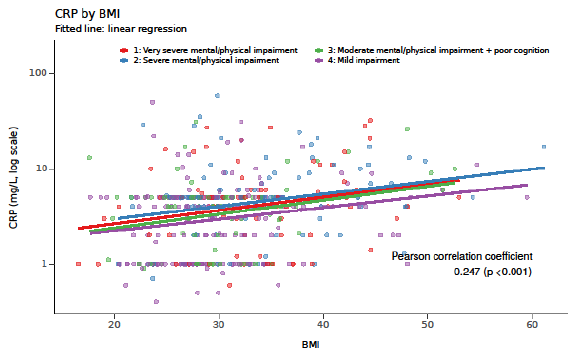

BMI = Body Mass Index kg/m^2^

21.	Acute heart failure: diagnosis and management. National Institute for Health and Care Excellence; 2014.

22.	Chronic heart failure in adults: diagnosis and management. National Institute for Health and Care Excellence; 2018.
